# Efficacy of a Spatial Repellent for Control of *Aedes*-Borne Virus Transmission: A Cluster Randomized Trial in Iquitos, Peru

**DOI:** 10.1101/2021.03.03.21252148

**Authors:** Amy C. Morrison, Robert C. Reiner, William H. Elson, Helvio Astete, Carolina Guevara, Clara del Aguila, Isabel Bazan, Crystyan Siles, Patricia Barrera, Anna B. Kawiecki, Christopher M. Barker, Gissella M. Vasquez, Karin Escobedo-Vargas, Carmen Flores-Mendoza, Alfredo A. Huaman, Mariana Leguia, Maria E. Silva, Sarah A. Jenkins, Wesley R. Campbell, Eugenio J. Abente, Robert D. Hontz, Valerie A. Paz-Soldan, John P. Grieco, Neil F. Lobo, Thomas W. Scott, Nicole L. Achee

## Abstract

Over half the world’s population is at risk for viruses transmitted by *Aedes* mosquitoes, such as, dengue and Zika. The primary vector, *Aedes aegypti*, thrives in urban environments. Despite decades of effort, cases and geographic range of *Aedes*-borne viruses (ABV) continue to expand. Rigorously proven vector control interventions that measure protective efficacy against ABV diseases is limited to *Wolbachia* in a single trial in Indonesia, and do not include any chemical intervention. Spatial repellents, a new option for efficient deployment, are designed to decrease human exposure to ABV by releasing active ingredients into the air that disrupt mosquito-human contact. A parallel, cluster-randomized controlled trial was conducted in Iquitos, Peru to quantify the impact of a transfluthrin-based spatial repellent on human ABV infection. From 2,907 households across 26 clusters (13 per arm), 1,578 participants were assessed for seroconversion (primary endpoint) by survival analysis. Incidence of acute disease was calculated among 16,683 participants (secondary endpoint). Adult mosquito collections were conducted to compare *Ae. aegypti* abundance, blood-fed rate and parity status through mixed effect difference-in-difference analyses. The spatial repellent significantly reduced ABV infection by 34·1% (1-sided 95% CI lower limit, 6·9%; 1-sided p-value=0·0236, z=1·98). *Aedes aegypti* abundance and blood-fed rates were significantly reduced by 28·6% (95% CI 24·1%, ∞); z=-9·11) and 12·4% (95% CI 4·2%, ∞); z=-2·43), respectively. Our trial provides the first conclusive statistical evidence from a pre-planned cluster randomized controlled clinical trial with a pre-defined effect size on the primary endpoint that was appropriate powered to prospectively quantify and statistically test for a difference in the impact of a chemical intervention, in this case a spatial repellent, to reduce the risk of ABV transmission compared to a placebo.

**Significance Statement:** Vector interventions are needed for *Aedes*-borne viral diseases (dengue, Zika, chikungunya, and yellow fever) prevention, but their application is hindered by the lack of evidence proving they prevent infection or disease. Our research reports the first conclusive statistical evidence from a pre-planned, prospective cluster-randomized, controlled clinical trial (cRCT) of significant protective efficacy (34.1% hazard estimate) against human *Aedes*-borne virus (ABV) infection by a chemical-based vector control intervention, the most commonly used intervention category among all ABV World Health Organization recommendations. A previous trial against malaria in Indonesia indicated a positive trend but did not detect a significant effect. Results from our ABV study will help guide public health authorities responsible for operational management and world-wide prevention of ABV, and incentivize new strategies for disease prevention.

## Introduction

*Aedes*-borne viral diseases (ABVD) [e.g., dengue (DENV), chikungunya (CHIKV), Zika (ZIKV), and yellow fever (YFV)] are devastating, expanding global public health threats that disproportionally affect low- and middle-income countries. Dengue, one of the most rapidly increasing vector-borne infectious diseases, results in ∼400 million infections each year [1,2], with four billion people at risk of infection annually [3]. Currently, the primary means for ABVD prevention is controlling the primary mosquito vector, *Aedes aegypti*. Existing vector control interventions, however, have failed to prevent ABV transmission and epidemics[4–6].

There is an urgent need to develop evidence-based guidance for the use of new and existing ABV vector control tools. The evidence-base for vector control against ABVs is weak, despite considerable government investments in WHO-recommended control of larval habitats (larviciding, container removal) and ultra-low volume (ULV) insecticide spraying [4,5,7–9]. These strategies continue to be implemented despite the lack of rigorously generated data from controlled clinical trials demonstrating they reduce ABV infection or disease [6]. The only ABV intervention with a proven epidemiological impact in a cluster randomized control trial (cRCT) assessed community mobilization to reduce mosquito larval habitats [10]. A recent test-negative trial with *Wolbachia*-infected mosquitoes reported a significant reduction of dengue illness in Indonesia[11].

Spatial repellents (SR) are devices that contain volatile active ingredients that disperse in air. The active ingredients can repel mosquitoes from entering a treated space, inhibit attraction to human host cues or disrupt mosquito biting and blood feeding behavior, and, thus, interfere with mosquito-human contact [12–14]. Any of these outcomes reduce the probability of pathogen transmission. Pyrethroid-based spatial repellents have shown efficacy in reducing malaria infections in China [15] and Indonesia [16]. There have, however, been no clinical trials evaluating the protective efficacy of spatial repellents against ABV infection or disease.

To generate evidence for public health consideration we conducted a double-blinded, parallel, cluster-randomized controlled trial (cRCT) to demonstrate and quantify the protective efficacy (PE) of a transfluthrin-based spatial repellent (SR) to reduce ABV infection incidence over two years in a human cohort in Iquitos, Peru.

## Results

We report results from the intervention phase of a cRCT, conducted in 26 clusters (13 per arm, see Methods and SI Section 1.2.1 for randomization scheme) each with approximately 140 households (60 qualifying participants) between August 2016 and March 2019 (**Figure 1-2, SI Section 1.1**). The primary endpoint was ABV seroconversion, as measured by DENV-or ZIKV-specific neutralizing antibodies, in blood from children ≥ 2 years to ≤ 18 years collected just prior to the deployment of the SR intervention, and approximately 1 and 2 years later. The SR intervention was a transfluthrin passive emanator placed in participating households according to manufacturer’s instructions, 1 product per 9 m^2^ and replaced at 15 d intervals (**Figure S1, SI Section 1.3.2**) during the two-year (two-transmission season) study period. Secondary endpoints were clinically apparent laboratory confirmed ABV disease and indoor female *Ae. aegypti*: 1) abundance, 2) blood-fed status (proxy for human-biting rates), and 3) parity status (proxy for age-structure). Participants followed for seroconversion were the ‘longitudinal cohort’ and those followed for disease were the ‘febrile surveillance cohort’.

**Figure 1.**
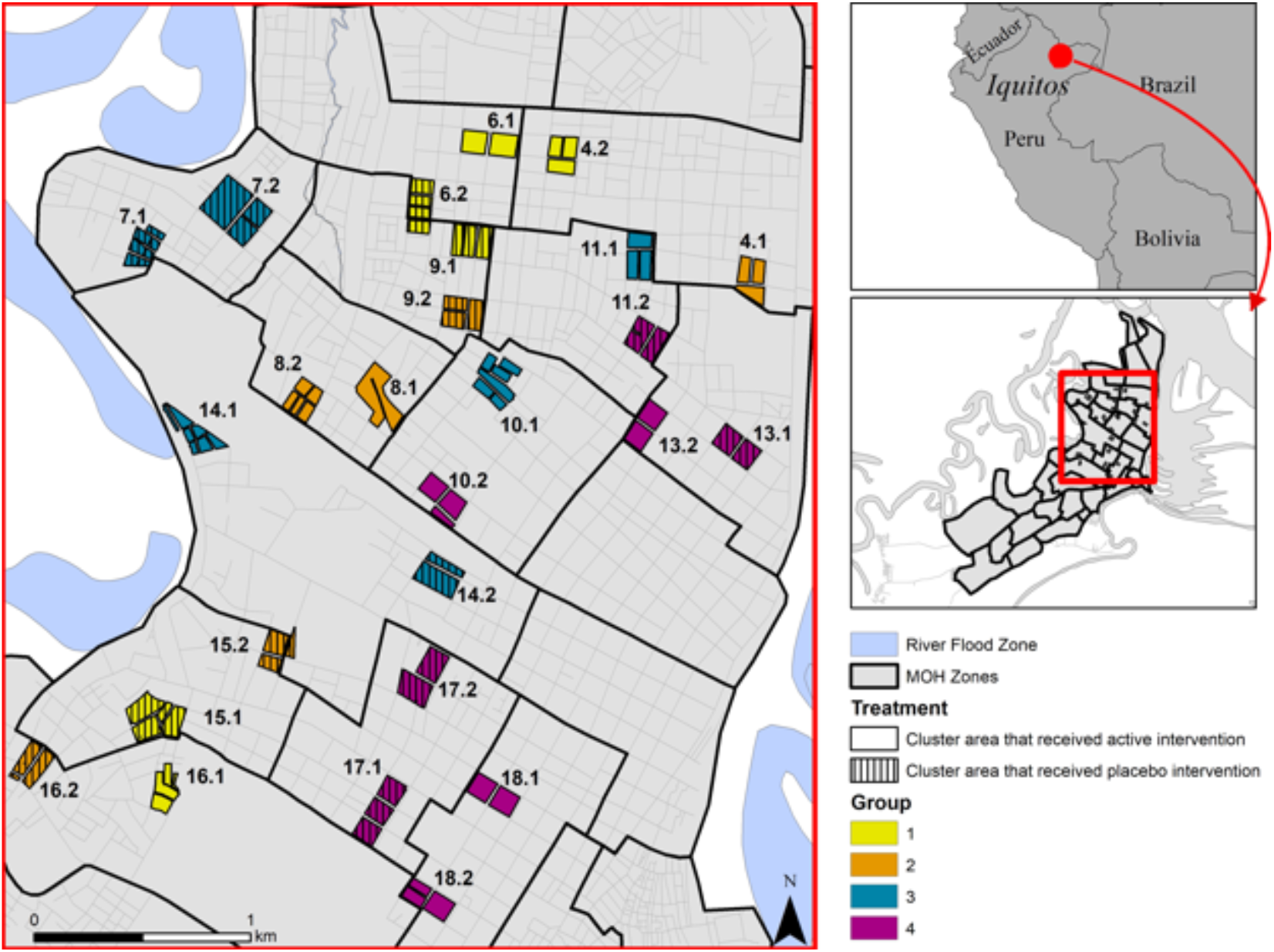
Location of 26 study clusters in Iquitos and Punchana Districts, Loreto Department, Iquitos, Peru. Each cluster consisted of ca. 140 households with an average distance of 300m between clusters.

**Figure 2.**
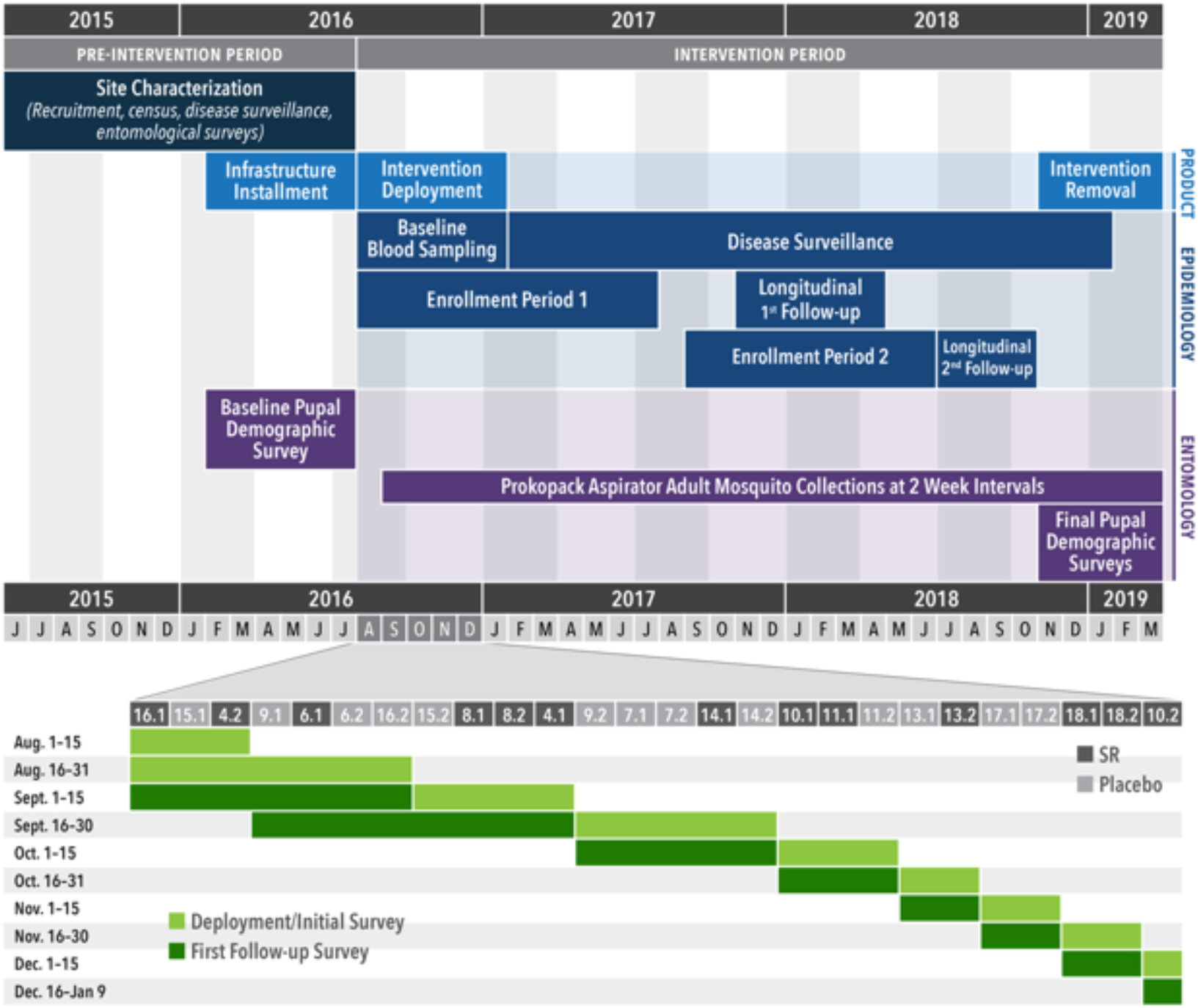
Study timeline. Top panel: Human blood-sampling, disease surveillance, and entomological monitoring in relation to deployment of the spatial repellent intervention. Bottom panel: Intervention roll-out between August-December 2016 by cluster. Horizontal numbers correspond to cluster numbers shown in Figure 1.

### STUDY POPULATION

A total of 2,215 persons were enrolled in the longitudinal cohort. Of these, 1,578 qualifying participants (individuals who were seronegative or had a monotypic DENV antibody response when they entered the trial) were included in the ITT analysis for seroconversion (**Figure 3**). Samples were tested by microneutralization enzyme immunoassay (MNT) for seroconversion to each DENV serotype and ZIKV (**SI Sections 1.3.3.1 and 1.3.4.1**). Only participants that provided at least 2 blood samples were included in final analyses. We observed a total of 196 ABV infections from 754 (1,090 paired samples) qualifying participants in the SR arm and 294 ABV infections from 824 (1,237 paired samples) qualifying participants in the placebo arm. Baseline covariates were balanced at both the cluster and individual level (**Table 1**).

**Figure 3.**
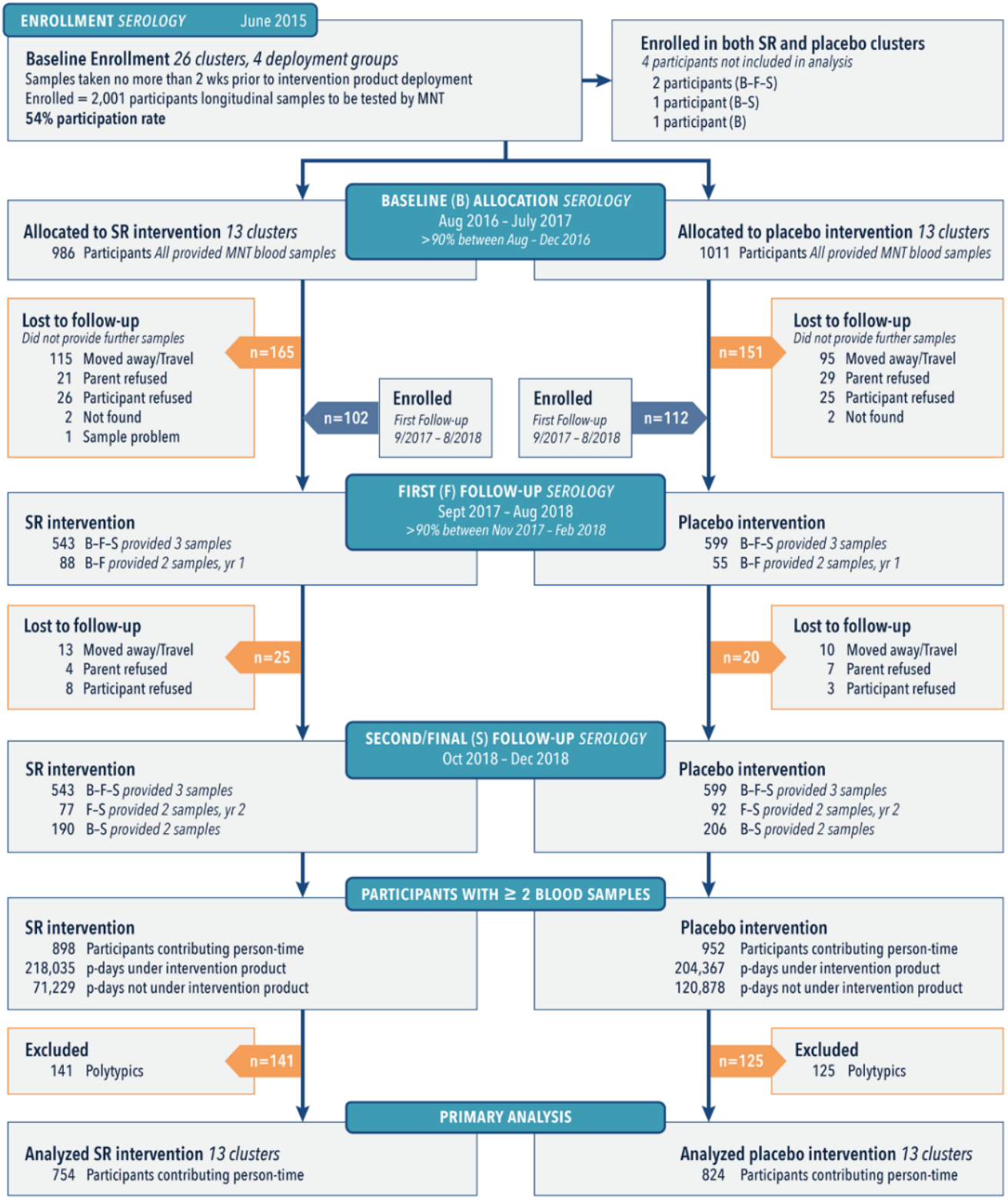
Allocation and follow-up of the longitudinal cohort population during three blood collection periods (Baseline [B], First [F] and Second/Final [S]). The majority (62%) of participants provided samples at each collection period, whereas some only participated during year 1 (B-F) or year 2 (F-S). Participants with a single sample were lost to follow-up and four individuals moved or had two houses located in the spatial repellent and placebo clusters are shown as removed at the baseline period for clarity.

**Table 1.**
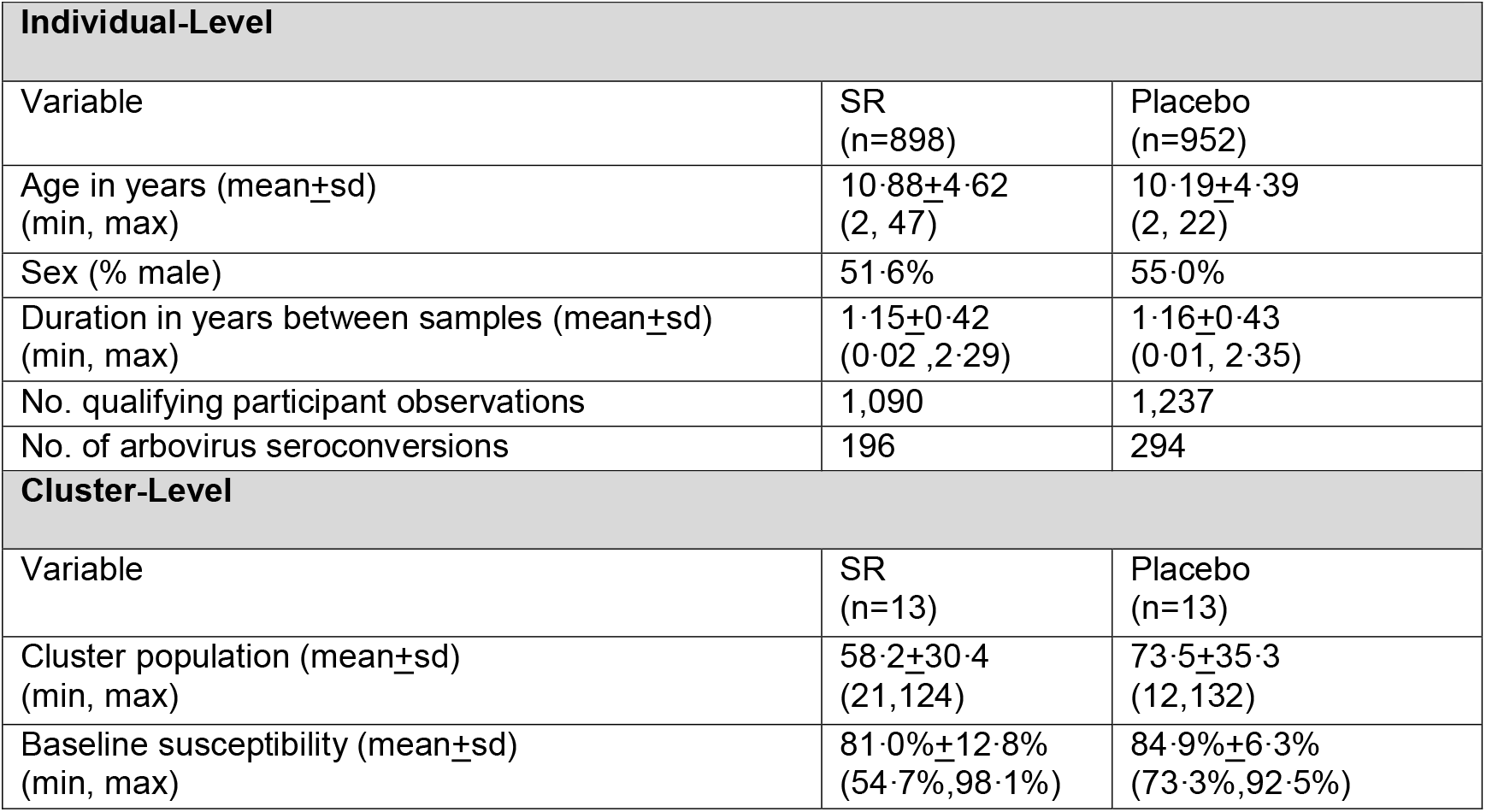
Summary of baseline characteristics, for qualifying participants of the intent-to-treat (ITT) longitudinal cohort population in spatial repellent (SR) and placebo arms. Qualifying participants were defined as individuals in a participating house that were seronegative or had a monotypic DENV antibody response when they entered the trial.

| **Individual-Level** |  |  |
| --- | --- | --- |
| Variable | SR<br>(n=898) | Placebo<br>(n=952) |
| Age in years (mean±sd)<br>(min, max) | 10.88±4.62<br>(2, 47) | 10.19±4.39<br>(2, 22) |
| Sex (% male) | 51.6% | 55.0% |
| Duration in years between samples (mean±sd)<br>(min, max) | 1.15±0.42<br>(0.02, 2.29) | 1.16±0.43<br>(0.01, 2.35) |
| No. qualifying participant observations | 1,090 | 1,237 |
| No. of arbovirus seroconversions | 196 | 294 |
| **Cluster-Level** |  |  |
| Variable | SR<br>(n=13) | Placebo<br>(n=13) |
| Cluster population (mean±sd)<br>(min, max) | 58.2±30.4<br>(21, 124) | 73.5±35.3<br>(12, 132) |
| Baseline susceptibility (mean±sd)<br>(min, max) | 81.0%±12.8%<br>(54.7%, 98.1%) | 84.9%±6.3%<br>(73.3%, 92.5%) |

A total of 16,707 participants were followed for clinical disease in the ‘febrile surveillance cohort’ through approximately 3 wellness visits per week, of which 16,683 were included in the ITT analysis (**Figure S2**). Suspected acute ABV cases that provided consent provided acute and convalescent blood samples and were monitored clinically daily. Acute serum samples were tested for viral RNA by PCR (DENV and ZIKV; **SI Sections 1.3.3.2-1.3.3.4**) and by ELISA for DENV IgM (**SI Sections 1.3.3.5**,**1.3.4.2**) to quantify the secondary endpoint of clinically apparent laboratory confirmation cases of ABV disease. Baseline covariates from the febrile surveillance cohort were balanced at both the cluster and individual level (**Table S4**).

### INTERVENTION COVERAGE

The household participation rate (intervention deployed at some point during the study period) per cluster (SR and placebo) was of 56·6 % (SD = 10·5%), with slightly more participation in SR versus placebo clusters (58·8% v 54·5%, p-value = 0·336). In households consenting to receive intervention (SR or placebo), the mean percentage of days covered by an intervention at the cluster-level was 81·6% (SD = 3·9), with slightly higher coverage in households assigned to SR intervention (82·9%) compared to households in the placebo arm (80·3%), albeit insignificant (p-value = 0·153, **Table S2, Figure S3**). For all enrolled households, the mean percentage of days with an adequate intervention application rate (1 product per 9 m^2^) was 73·6% (SD = 9·1), with similar rates between SR and placebo clusters (72·7% versus 74·5%, p-value > 0·999, **Table S2**).

### SPATIAL REPELLENT EFFICACY

We used survival analysis with proportional hazards model with an exponential distribution assumption for baseline hazard to estimate a PE (**SI Section 1.5.2.1**). The estimated PE of the SR intervention was 34·1% (1-sided 95% CI lower limit, 6·9%) (**Table 2**). Reduction in the arbovirus infection hazard rate was significant at the 5% significance level (Test statistic: *z*=1·98, 1-sided p-value = 0·02). Baseline covariates included in the statistical model on the hazard of arbovirus infection in qualifying participants were age, which had statistically significant effects on the hazard of arbovirus infection in qualifying participants, and sex, which did not. Reported age- and sex-specific hazard rate changes are conditional. Hazard rate increases by 4·6% for every one-year increase in age. Hazard rate decreases by 4·4% in males relative to females (**Table 2**). The originally proposed ITT mixed effects logistic regression analysis (**SI Section 1.5.2.2**), which ignores differential participation duration across qualifying participants, produced a result consistent with those presented here; i.e., PE > 30% and statistical significance at the 5% level (**Table 2**).

**Table 2.**
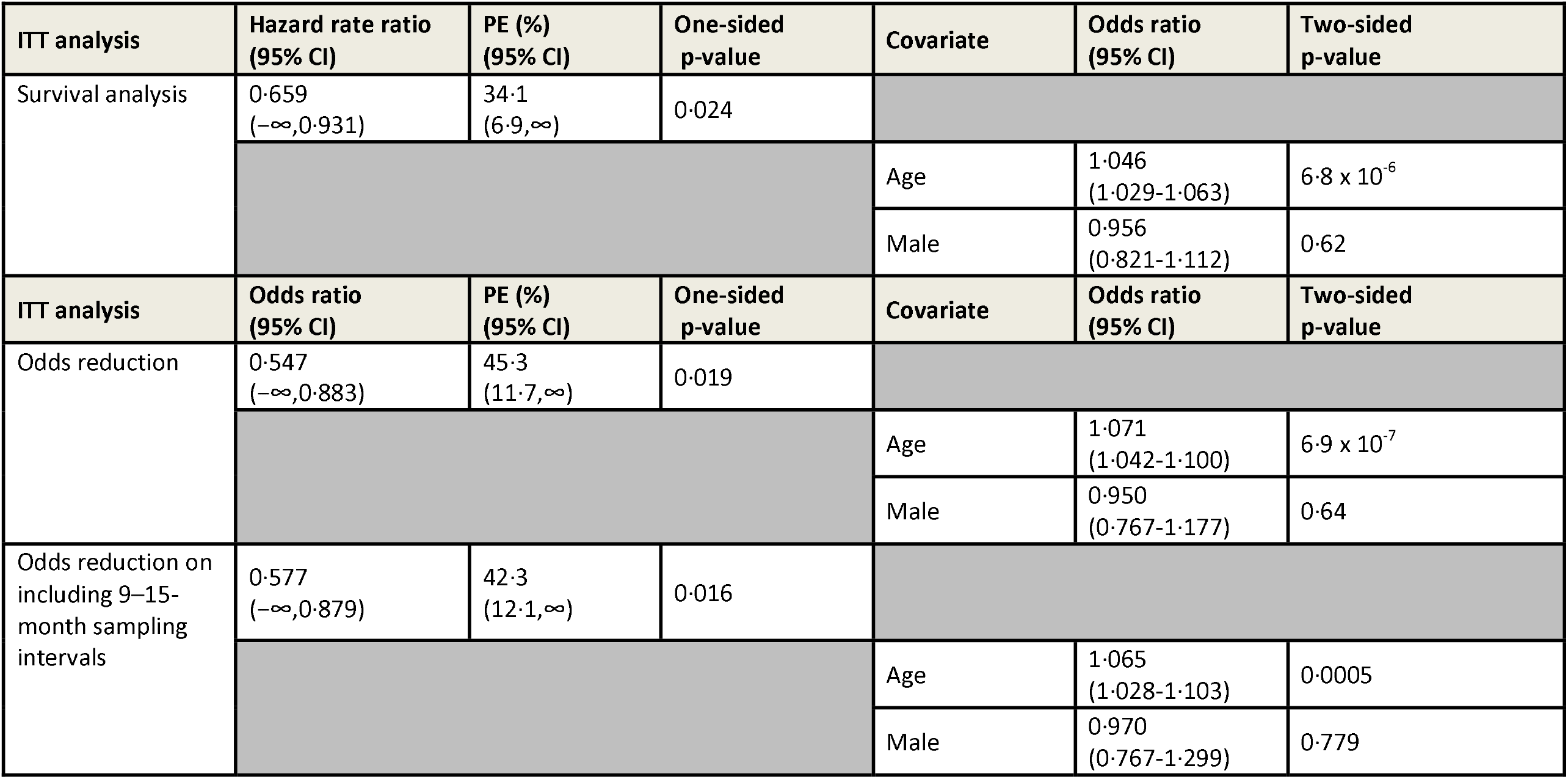
Protective efficacy (PE) estimates from intent to treat (ITT) analyses for the spatial repellent intervention against *Aedes*-borne virus infection in qualifying participants measured by seroconversion (primary endpoint), including covariate effects.

The Kaplan-Meier curves of arbovirus infection for qualifying participants by cluster show considerable between-cluster variation (SR and placebo clusters) as evidenced by the wide spread of survival curves (Figure 4). For example, there were no arbovirus infections in qualifying participants in placebo Cluster 7.2, which had only 18 qualifying participants. Conversely, of the 11 qualifying participants who had a duration of at least 15 months between tests in Cluster 8.2, a total of 5 subjects became infected since their last test. The duration between tests varied by participant and across clusters (**Figure S4**), resulting in some Kaplan-Meier curves being estimated beyond two years. In many of those clusters, the only participants that went over two years between blood sampling were universally found to have had an arboviral infection.

**Figure 4.**
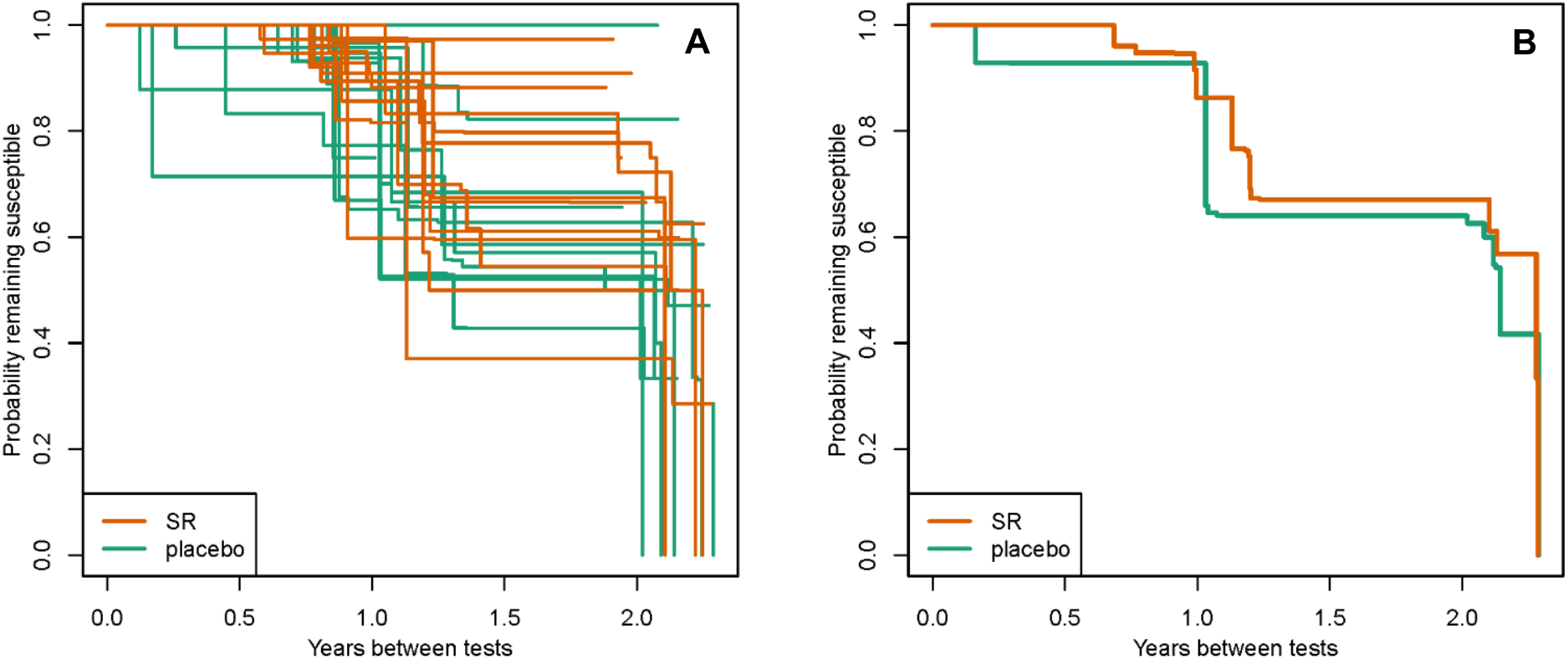
Kaplan-Meier curves of arbovirus infection for 13 spatial repellent (SR) and 13 placebo clusters in qualifying participants measured by seroconversion (primary endpoint) by cluster. Panel A, Hazard rates by individual cluster. Panel B, Aggregated hazard rate.

A Poisson generalized linear regression was used to assess intervention impact on clinical disease, with an offset for the number of participant-days each participant spent in each cluster. No covariates were used and due to the small sample size, no random effects were incorporated (**SI section 1.5.3**). No statistical difference between incidence in the SR and Placebo arms was detected. Baseline characteristics of covariates included in the analysis of PCR/ELISA confirmed DENV and ZIKV cases were balanced between SR and placebo arms **(Table S5)**. Results from an ITT fixed effect Poisson generalized linear model indicate the rate ratio is 1.144 with an upper bound on the 1-sided 95% CI of 1.601. This translates into a 14.4% increase in the rate of PCR/ELISA confirmed arbovirus infections by SR intervention compared with placebo with the lower bound of the 1-sided 95% CI of –60.1%. This apparent increase in the intervention area was not statistically significant at the 5% level (Test statistic: *z* = -0.975), in part because only 96 disease cases were detected, 51 in the SR arm and 45 in the placebo arm during 10,793,792 participant days that appeared balanced between SR and placebo clusters **(Table S4, SI Section 2.5.2.1**).

The estimated reduction in adult female *Ae. aegypti* abundance in clusters receiving SR intervention was 28·6% (1-sided 95% CI lower limit: 24·1%, test statistic: z= -9·11) using mixed effects difference-in-difference (DID) Poisson regression, with factor-level covariates (**Table 3, SI Section 1.5.4**). Baseline mosquito abundance was balanced between treatment arms (**Table S5**) with post-baseline quantities estimated based on 47,518 and 43,417 household collections in SR and placebo arms, respectively. Baseline abundance averaged 0·277 (standard deviation [SD] 0·153) and 0·279 (SD 0·122) per house survey in SR and placebo arms, respectively, whereas post-baseline abundance averaged 0·276 (SD 0·091) and 0·391 (SD 0·142) in the SR and placebo arms, respectively (**Table 3**). There was strong indication of seasonality, with estimated z-scores of 6 or greater when comparing each month to the reference month of January (**Table S7**). Overall, abundance trended lower in the SR clusters compared to the placebo clusters after intervention deployment for the duration of the trial, from 2017-2019 (**Figure 5**). Post-intervention entomological surveys indicate that this difference disappeared after removal of the intervention (**Figure S8B**).

**Table 3.**
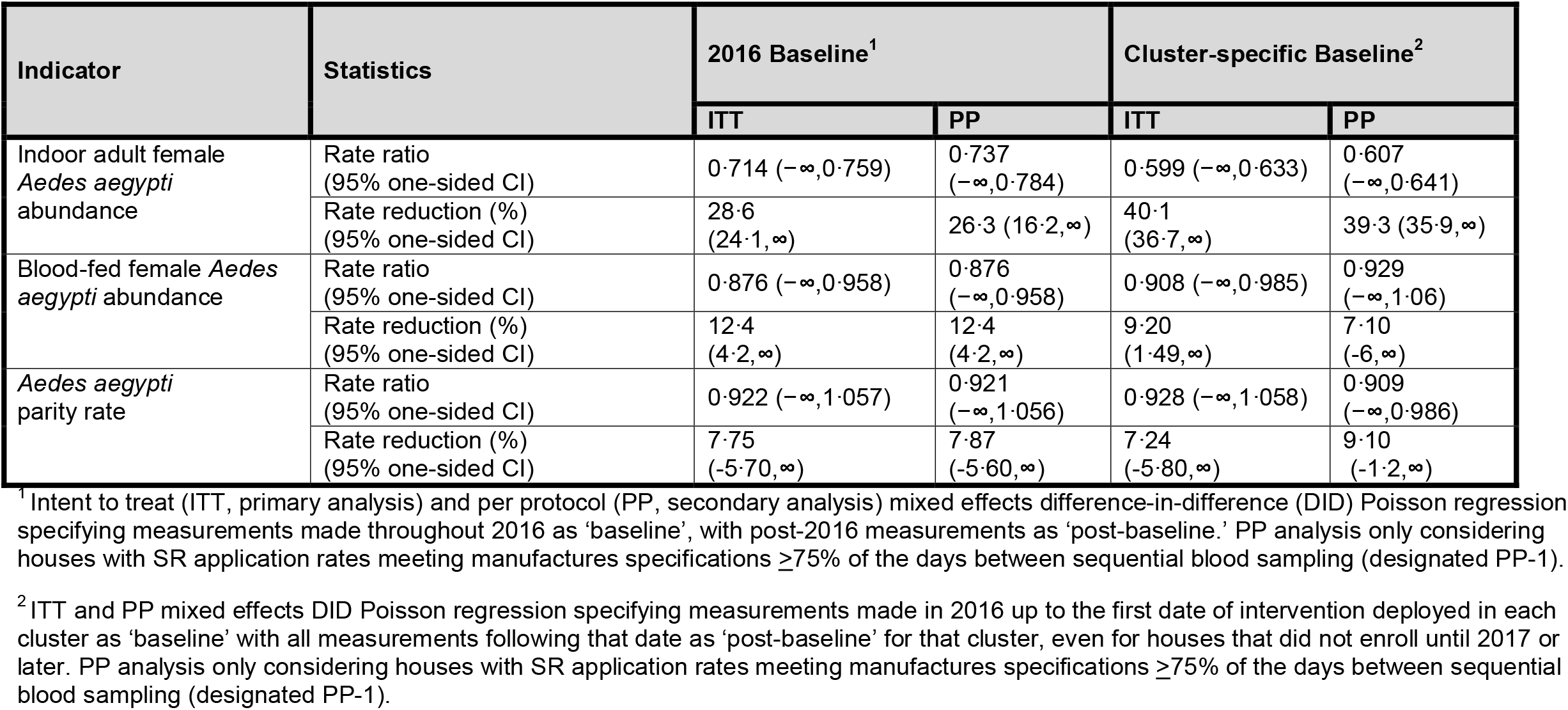
Rate reduction summary of indoor adult female *Aedes aegypti* abundance, blood-fed female abundance, and parity rates (secondary endpoints) for primary (ITT) and secondary analyses (PP). No correction for multiple testing was performed for secondary endpoint analyses and, as such, in accordance with CONSORT guidelines p-values are not presented.

| Indicator | Statistics | 2016 Baseline<sup>1</sup> |  | Cluster-specific Baseline<sup>2</sup> |  |
| --- | --- | --- | --- | --- | --- |
|  |  | ITT | PP | ITT | PP |
| Indoor adult female *Aedes aegypti* abundance | Rate ratio (95% one-sided CI) | 0.714 ($-\infty, 0.759$) | 0.737 ($-\infty, 0.784$) | 0.599 ($-\infty, 0.633$) | 0.607 ($-\infty, 0.641$) |
| | Rate reduction (%) (95% one-sided CI) | 28.6 (24.1, ∞) | 26.3 (16.2, ∞) | 40.1 (36.7, ∞) | 39.3 (35.9, ∞) |
| Blood-fed female *Aedes aegypti* abundance | Rate ratio (95% one-sided CI) | 0.876 ($-\infty, 0.958$) | 0.876 ($-\infty, 0.958$) | 0.908 ($-\infty, 0.985$) | 0.929 ($-\infty, 1.06$) |
| | Rate reduction (%) (95% one-sided CI) | 12.4 (4.2, ∞) | 12.4 (4.2, ∞) | 9.20 (1.49, ∞) | 7.10 (-6, ∞) |
| *Aedes aegypti* parity rate | Rate ratio (95% one-sided CI) | 0.922 ($-\infty, 1.057$) | 0.921 ($-\infty, 1.056$) | 0.928 ($-\infty, 1.058$) | 0.909 ($-\infty, 0.986$) |
| | Rate reduction (%) (95% one-sided CI) | 7.75 (-5.70, ∞) | 7.87 (-5.60, ∞) | 7.24 (-5.80, ∞) | 9.10 (-1.2, ∞) |

<sup>1</sup> Intent to treat (ITT, primary analysis) and per protocol (PP, secondary analysis) mixed effects difference-in-difference (DID) Poisson regression specifying measurements made throughout 2016 as 'baseline', with post-2016 measurements as 'post-baseline.' PP analysis only considering houses with SR application rates meeting manufactures specifications $\geq 75\%$ of the days between sequential blood sampling (designated PP-1).
<sup>2</sup> ITT and PP mixed effects DID Poisson regression specifying measurements made in 2016 up to the first date of intervention deployed in each cluster as 'baseline' with all measurements following that date as 'post-baseline' for that cluster, even for houses that did not enroll until 2017 or later. PP analysis only considering houses with SR application rates meeting manufactures specifications $\geq 75\%$ of the days between sequential blood sampling (designated PP-1).

**Figure 5.**
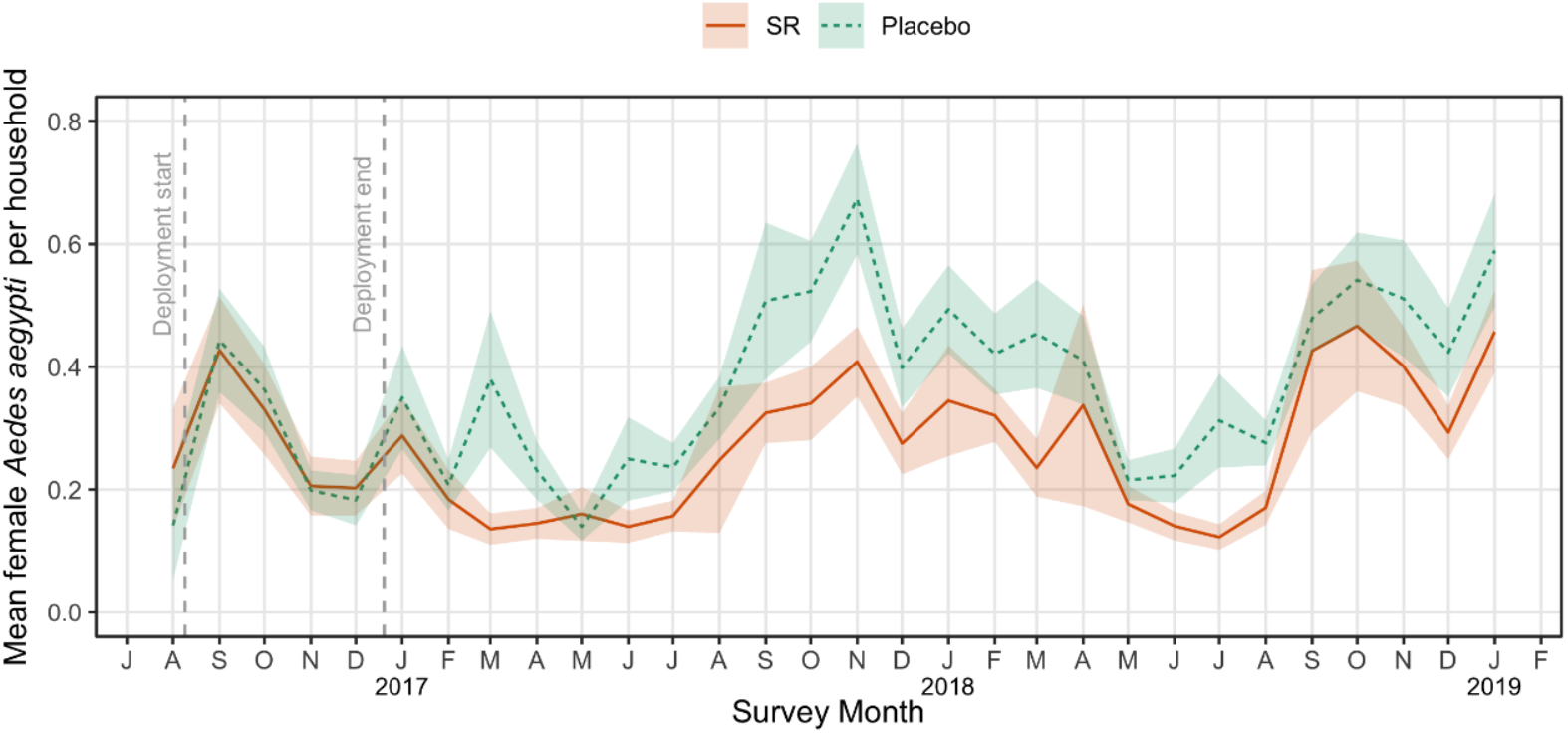
Mean densities of adult female *Aedes aegypti* collected per household survey in 13 spatial repellent (SR) and 13 placebo clusters by study month. Shaded areas represent the 95% Confidence Interval around the mean.

The estimated reduction in the rate of blood-fed *Ae. aegypti* collected inside houses was 12·4% (1-sided 95% CI lower limit: 4·2%, test statistic: z= -2·430) also using mixed effects DID Poisson regression (**Table 3**). Baseline abundance of engorged *Ae. aegypti* was balanced across treatment arms (**Table S5**) with post-baseline quantities estimated based on 9,257 and 11,496 mosquitoes assessed for blood-fed status in SR and placebo arms, respectively. Baseline abundance of blood-fed mosquitoes averaged 0·593 (standard deviation 0·440) and 0·536 (standard deviation 0·451) per collection in SR and placebo arms, respectively. Post-baseline rates averaged 0·606 (SD 0·460) and 0·634 (SD 0·447) in SR and placebo arms, respectively. There was no strong indication of seasonality (**Table S8**).

There was no observed intervention effect (**Table 3**) or indication of seasonality (**Table S9**) based on the parity rate of *Ae. aegypti* females.

### ADVERSE EVENTS (AE) AND SERIOUS ADVERSE EVENTS (SAE)

Twenty-nine AEs were reported during the trial (**SI Section 2.8**). Of these, three were associated with blood draw (vasovagal response, two from SR and one from placebo clusters). The remaining 26 AEs reflected symptoms consistent with pyrethroid/ transfluthrin exposure. Reporting of these AEs was higher in the SR clusters (22 of 8,235 subjects: 0.267%) than the placebo clusters (4 of 8,448 subjects: 0.047%). The relative risk of experiencing a mild adverse event due to the SR product was 5.6 (95% CI: 1.9-16, p=0.0015). The 26 affected individuals (26 of 16,707 subjects; 0.155%) came from 18 households, often reporting different combinations of the following symptoms: allergic response with itching and skin irritations (n=19; 15 from SR, four from placebo clusters), dry mouth/bad taste (n=5, from SR clusters only), breathing issues (n=7, from SR clusters only) including exacerbation of chronic bronchitis or asthma (n=2), and headaches (n=4, from SR clusters only). The total number of AEs reported from longitudinal subject cohort households during the trial was similarly low (18 of 2,907 households; 0.619%). No reported SAEs (deaths) were deemed associated with the spatial repellent intervention.

## Discussion

*Aedes*-borne viruses have expanding regions of transmission, cause increasingly frequent epidemics, and are transmitted by one of the most anthropophilic mosquitoes. There is, therefore, a growing unmet need for effective ABV disease prevention [9]. Our cRCT provides the first conclusive statistical evidence from a pre-planned, prospective cluster-randomized, controlled clinical trial (cRCT) of significant protective efficacy (34.1% hazard estimate) against human *Aedes*-borne virus (ABV) infection by a chemical-based vector control intervention, in this case a spatial repellent. The uniqueness of our trial in the realm of *Aedes* vector control is that it is 1) the first pre-planned, cluster randomized controlled clinical trial 2) with a pre-defined effect size on the primary endpoint of human infection 3) that was appropriately powered to prospectively quantify and statistically test for a difference in the impact of a chemical intervention against *Aedes-*borne virus human infection incidence compared to a placebo. Currently used adult *Aedes* chemical control strategies are not supported by such evidence. Reduced human infection caused by a reduction in mosquito biting supports development of (1) improved repellent formulations and (2) enhanced methodologies for broad scale application.

PE was detected despite assumed dilution and contamination effects due to participant movement in and out of study clusters. Unlike a vaccine, SR protected study participants in their own homes or another protected home within their neighborhood (i.e., Study Cluster), but did not provide continuous protection after they left treated houses. Our cohort, tested for ABV seroconversion, was comprised principally of children ≤ 17 years of age, most whom attended schools that have been shown in Iquitos to be of lower risk of *Ae. aegypti* infestation than residential sites [17]. Our outcome was demonstrated in an operational context, reflecting complex interactions among ongoing Ministry of Health interventions across the study area, imperfect coverage at the household-level (rooms closed to intervention, homeowner removal and/or loss of intervention), <100% household participation within clusters, and the suggestion of pyrethroid resistance in the local *Ae. aegypti* population [18] (**SI section 2.7**). Reduced risk of ABV infection was associated with a significant reduction in indoor female *Ae. aegypti* abundance and blood feeding. Although entomological outcomes were modest, detected effects are consistent with the expected mode of SR action (i.e., deterrence from house entry and/or interfering with human-biting) [12,19] and the impact waned after the intervention was removed at the end of the trial.

Our results support SRs as a flexible class of vector control products with positive public health impact not limited to ABV diseases. Transfluthrin [15] and metofluthrin-based[20] mosquito coils have been shown to reduce malaria and the same spatial repellent device used in our Iquitos trial reduced malaria infections in an Indonesia cRCT[16]. The SR product we tested was generally well tolerated even though it produced mild skin and respiratory irritation, a well know side effect of pyrethroids. Our trial was the first to quantify these types of adverse events for a chemical intervention in a double-blinded trial. Our results, therefore, support the potential for SRs to reduce a variety of vector-borne diseases, augment existing public health efforts and support SRs as an effective component in vector control intervention strategies. To facilitate implementation and programmatic scale-up, additional assessments, which have already begun [21–24], are needed.

Our Peru cRCT is one of two trials recommended by the WHO for assessing public health value and developing global health policy for the SR intervention class [25]. Our study was powered to detect a 30% reduction in ABV infection risk, not acute ABV disease nor virus infection rates in mosquitos. During the trial period, dengue prevalence was lower than previous years and a Zika epidemic occurred in 2016 [26]. This epidemiological uncertainty is typical of ABV transmission, making powering ABV cRCTs challenging and helps explain why cRCTs with epidemiological outcomes for ABVs are rare [27]. We used seroconversion as our primary endpoint of PE to address this challenge. At our Iquitos study site, powering a trial based on clinically apparent ABV disease would not be logistically feasible.

Fully integrating vector control into ABV disease prevention programs will require quantitative guidance based on quantitative measures of the impact from each intervention component. Ministries of Health, local to national governments and non-governmental organizations can use our trial results as an evidence-base for informed application of SRs. Considering the growing ABV public health threat, difficulties of developing vaccines against multiple viruses and past, poorly informed vector control failures [6], enhanced ABV disease prevention will benefit greatly from interventions with proven public health value.

## Materials and Methods

This trial is registered with ClinicalTrials.gov, number NCT03553277.

### ETHICAL STATEMENT

Our study protocol (#NAMRU6.2014.0021, Supplementary Information (SI)) was approved by the U.S. Naval Medical Research Unit No. 6 (NAMRU-6) Institutional Review Board (IRB), which includes Peruvian representation and complies with US Federal and Peruvian regulations governing the protections of human subjects, and the Regional Health Authority (DIRESA), the local branch of the Peruvian Ministry of Health. IRB authorization agreements were established between the NAMRU-6, the University of Notre Dame (Sponsor), the University of California at Davis, and the University of Washington.

### TRIAL DESIGN

Detailed study methods are provided in the SI.

Our trial was conducted from June 2015 through March 2019 in the Iquitos and Punchana Districts of Iquitos, Peru (**Figure 1, SI Section 1.1**). Clusters were selected in January 2015. Enrollment began June 2015. Participation included: 1) a house census, 2) disease surveillance, 3) annual blood draws, 4) bi-monthly entomological surveys, and 5) intervention application in the house. Epidemiological monitoring and entomological surveillance lasted from February 2016 through March 2019 (**Figure 2**).

Our main objective was to demonstrate and quantify the protective efficacy (PE) of a SR in reducing ABV infection incidence in a human cohort. Qualifying participants were individuals in a participating house who were seronegative or had a monotypic DENV antibody response when they entered the trial. Assuming the probability of seroconversion for seronegative or monotypic individuals was 10% with a coefficient of variation of 0·25, and an alpha of 5%, we estimated we would need 26 clusters (13 per arm) with approximately 60 qualifying individuals to achieve a power of 80% to detect a reduction in the odds of 30%.

The primary endpoint was ABV seroconversion, as measured by DENV-or ZIKV-specific neutralizing antibodies, in blood from children ≥ 2 years to ≤ 18 years. To increase the pool of baseline seronegative participants, we expanded screening to ≥18 years. Secondary endpoints were clinically apparent laboratory confirmed ABV disease and indoor female *Ae*. aegypti: 1) abundance, 2) blood-fed status (proxy for human-biting rates), and 3) parity status (proxy for age-structure). Participants followed for seroconversion were the ‘longitudinal cohort’ and those followed for disease were the ‘febrile surveillance cohort’.

### RANDOMIZATION AND INTERVENTION

A total of 26 clusters (13 per arm) each with approximately 140 households (60 qualifying participants) were randomly allocated in August 2016 to receive SR or placebo intervention by the external statistician serving on the Data Safety Monitoring Board (DSMB) using a random number generator (https://www.random.org) (**SI Section 1.2.1**). Investigators, research staff, and study participants were blinded to cluster allocation. Our intervention was a transfluthrin passive emanator designed and produced by SC Johnson (Racine, WI), replaced at 2-week intervals, as described previously (**Figure S1, SI Section 1.3.2**)[16]. Our trial was specifically designed to evaluate the protective efficacy of the first in class spatial repellent product prototype to enable WHO assessment for public health value of the spatial repellent product class. Spatial repellent products are not yet recommended by WHO for inclusion into programmatic vector control strategies. We selected the SCJ manufactured, transfluthrin-based passive emanator because it represents the first in class prototype of a spatial repellent under WHO public health value assessment[28]. The spatial repellent intervention used in our study is the same intervention evaluated against malaria infections in Sumba Indonesia[16]. Spatial repellent and placebo intervention had identical packaging and were deployed in houses by study personnel using a blinded coding scheme. The placement of the intervention followed manufacturer specifications for indoor use conditions.

### SEROCONVERSION AND DISEASE SURVEILLANCE

Recruitment for the longitudinal cohort focused on children because they were likely to be antibody test-negative or monotypic at baseline than adults, which would facilitate interpretation of laboratory assays, and less mobile than adults[29], thus spending more time in their houses or their assigned cluster. Baseline blood samples were obtained within 2 weeks before or after initial intervention deployment. As new families moved into the study area, they were recruited to participate, resulting in longitudinal participant enrollment throughout the interval between Baseline (B), First (F), and Second/Final (S) longitudinal blood draws (**Figure 3**). Samples were tested by microneutralization enzyme immunoassay (MNT) for seroconversion to each DENV serotype and ZIKV (**SI Sections 1.3.3.1 and 1.3.4.1**). Only participants that provided at least 2 blood samples were included in final analyses.

The febrile surveillance cohort was recruited by nurse technicians during door-to-door wellness checks starting with the first week of intervention deployment. Suspected cases exhibited axillary temperature of ≥ 37·5°C or, for suspected ZIKV infection, absence of fever but presence of rash, arthralgia, arthritis, or non-purulent conjunctivitis for ≤ five days. Participants meeting these criteria provided acute and convalescent (14–21 days later) serum samples and were monitored clinically daily. Acute serum samples were tested for viral RNA by PCR (DENV and ZIKV; **SI Sections 1.3.3.2-1.3.3.4**) and by ELISA for DENV IgM (**SI Sections 1.3.3.5**,**1.3.4.2**).

### ENTOMOLOGICAL ENDPOINTS

Indoor Prokopack aspirations [30] were conducted in all consented homes at time of first intervention deployment and subsequent intervention replacement; i.e., two-week intervals. Adult mosquitoes were transported to the NAMRU-6 Iquitos laboratory, sedated at 4°C, identified to species and sex, counted by date and house. Up to 30 female *Ae. aegypti* per household per collection were examined for blood meal status and scored as unfed, blood-fed (fully engorged, half-engorged, or trace amounts), or gravid [31]. These female mosquitoes were then dissected to determine their parity status (parous, nulliparous, or gravid) (**SI Section 1.3.5.1**). Standard insecticide resistance assays were used to assess vector susceptibility to transfluthrin one-year into the trial (**SI Section 1.3.5.2**).

### SAFETY MONITORING

Adverse events (AEs) and serious adverse events (SAEs) were actively collected throughout the trial during surveillance follow-up and entomological surveys (**SI Section 1.3.6**). Reported AEs were investigated by study staff and appropriate care was recommended by a study physician within 24 hours. Safety reporting to the NAMRU-6 IRB was managed by UC Davis in accordance with the approved protocol. Quarterly reports summarizing reported AEs and SAEs were reviewed by the DSMB for trial safety assessment.

### STATISTICAL ANALYSIS

Details of our analytical approach are provided in the statistical analysis plan (**SAP**) and **SI Section 1.5**. All analyses were conducted by RCR using R 3.6.1 (R Core Team)[32] and the Ime4[33] and survival[34] packages. No correction for multiple testing was performed for secondary endpoint analyses and, as such, in accordance with CONSORT guidelines, in those cases we do not present p-values.

The choice of the investigators to select and report a one-sided p-value and corresponding one-sided confidence interval is based on the underlying assumption associated with the intervention; i.e., the spatial repellent will not increase risk/harm (disease) over the standard of care. The statistical translation is that we are testing to see if a) the intervention is no better than the standard of care or b) the intervention is better than the standard of care (superior). As this is a one-sided test of superiority, we present one-sided test results. We also provide the null and alternative hypotheses, test statistic, and its assumed distribution to provide readers information to interpret the p-value and confidence interval.

The primary analysis was an intent-to-treat (ITT) assessment of ABV seroconversion for all qualifying participants per treatment assignment who were >2 years to <18 years of age. Due to the rolling nature of enrollment, we used a survival analysis with a proportional hazards model and exponential distribution assumption for the baseline hazard (i.e., constant baseline hazard through time) and a frailty component to account for correlation within clusters. If *h*(*t*_*ij*_ | *x*_*ij*_) is the hazard rate of the *j*^*th*^ individual in the *i*^*th*^ cluster with covariate values *x*_ij_ then this individual’s hazard rate of an arbovirus infection can be written as:

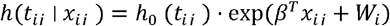

where 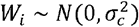 is the random effect of the *i*^*th*^ cluster. Covariates included age, sex, and treatment status (SR or placebo). PE was estimated as 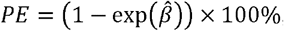, where 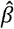 is the estimated regression coefficient for the intervention group and 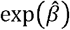 is the estimated hazard ratio (HR) between SR and placebo. The null hypothesis of PE = 0% is equivalent to *β* = 0, which is tested by Wald’s test *z* = *β*/*s*, where *s* is the estimated standard error of 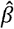, at the 1-sided significance level of 5%.

A Poisson generalized linear regression was used to assess intervention impact on clinical disease, with an offset for the number of participant-days each participant spent in each cluster. No covariates were used and due to the small sample size, no random effects were incorporated.

Indoor adult female *Ae. aegypti* abundance, blood-fed rates, and parity were tested through difference-in-difference Poisson generalized linear mixed models. Collections conducted in 2016 were defined as ‘baseline’ and collections conducted in 2017 and 2018 were estimated as ‘post-baseline’. Each analysis accounted for month of year and year as fixed effects and contained a random effect by cluster. Model formulation details are presented in **SI Sections 1.5.4-1.5.6**.

The primary analysis conducted differs from that in the original SAP (**SI SAP**). Procedures called for yearly blood draws from children. To increase the number of qualifying participants, we expanded the age range of the longitudinal cohort to all ages ≥ 2yrs. The rolling enrollment resulted in substantial variation in time intervals between blood draws. Simple logistic regression, therefore, was inadequate. The decision to alter the primary analysis was discussed and agreed upon by the study statistician and the external DSMB statistician before outputs were unblinded to the DSMB statistician.

## Supporting information

S2. Initial IRB Approved Study Protocol

S3. Final IRB Approved Study Protocol

S4. Statistical Analyses Plan

S5. CONSORT Checklist

## Data Availability

De-identified analytical datasets and statistical codes will be made available upon pre-print posting. The data will be made available without investigator support to anyone requesting the data and distributed under the terms of the Creative Commons Attribution (CC-BY) License which permits unrestricted use, distribution, and reproduction in any medium, provided the original author and source are credited. Requests for analytical datasets and statistical codes should be sent to.

## Acknowledgments and Funding Sources

We extend sincere appreciation to DSMB members: Dennis Shanks, Chris Drakeley, and Neal Alexander for assurances on data integrity and safety assessments. We thank the World Health Organization Vector Control Advisory Group (WHO VCAG) for their comments during project assessments. We are grateful for the efforts of Marianne Kent, Program Coordinator at the University of Notre Dame, for her administrative, and logistical support of the program, as well as Brett Fox of the University of Notre Dame Center for Research Computing for assisting with central database development for data verification. We thank Kristina Davis and Matthew Sisk of the University of Notre Dame Center for Research Computing and Hesburgh Libraries, respectively, for their assistance in graphic development. We acknowledge Audrey Lenhart of the Centers for Disease Control and Prevention (CDC) in Atlanta, GA USA and J. Kevin Baird of the Eijkman-Oxford Clinical Research Unit, Jakarta, Indonesia and Nuffield Department of Medicine, Centre for Tropical Medicine, University of Oxford, Oxford, United Kingdom for their role on the Scientific Advisory Committee.

We are grateful to SC Johnson for providing integral industry and product expertise including the independent funding of the development, manufacturing, delivery, and shipment of the spatial repellent and placebo products used in the study. SC Johnson provided expertise in ensuring intervention quality, storage, application and disposal assurances throughout the trial.

We thank the residents of Iquitos for their support and participation in this study and willingness to allow this study to be conducted in their homes and community. We greatly appreciate support of the Loreto Regional Health Department, including Drs. Hugo Rodriguez-Ferruci, Christian Carey, Carlos Alvarez, and Lic. Wilma Casanova Rojas, who facilitated our work in Iquitos. We thank the Naval Medical Research Unit-6 (NAMRU-6) Virology and Emerging Infections Department (VEID) leadership who provided institutional support, IRB guidance, and support supervising field staff. We appreciate the commentary and advice provided by the NAMRU-6 IRB and Research Administration Program for the duration of this study. We thank the NAMRU-6 VEID field team which provided daily support through the duration of the project and without whom the recruitment of acute dengue cases would not have been possible. We thank Cecilia Gonzales, Gloria Talledo, and Gabriela Vasquez de la Torre for their administrative support of the project.

