## Supplementary material for "Efficacy of a Spatial Repellent for Control of *Aedes*-Borne Virus Transmission: A Cluster Randomized Trial in Iquitos, Peru": S1. Supplementary Appendix

This appendix has been provided by the authors to give readers additional information about their work.

1.5.2.3 Secondary analysis – ITT mixed effects logistic regression (adjusted from original SAP)... 17

Supplemental Figure S6. Hazard rate reduction summary of PP-2 secondary analysis. Regression plot of hazard risk reduction based on the fraction of time a qualifying participant house had at least as many SR products as recommended between sequential blood sampling. This analysis reconsidered the threshold for inclusion in per protocol analysis and assessed the hazard risk reduction (fixed effect) as a function of inclusion thresholds (fraction covered), which ranged from 5% to 100%. As the coverage inclusion criteria became more strict (100%), fewer individuals were included in the analysis, resulting in fewer total seroconversions (primary endpoint) among all qualifying participants in SR and placebo clusters. Mean results from the primary analysis are included as a grey dashed horizontal line for reference. ..... 34

Supplemental Figure S7. Hazard rate reduction summary of PP-4 secondary analysis. Regression plot of hazard risk reduction based on the fraction of time a qualifying participant house had any SR products between sequential blood sampling. This analysis reconsidered the threshold for inclusion in per protocol analysis and assessed the hazard risk reduction (fixed effect) as a function of inclusion thresholds (fraction covered), which ranged from 5% to 100%. As the coverage inclusion criteria became more strict (100%), fewer individuals were included in the analysis, resulting in fewer total seroconversions (primary endpoint) between all qualifying participants in SR and placebo clusters. Mean results from the primary analysis are included as a grey dashed horizontal line for reference .. 35

Supplemental Figure S8. Mean densities of adult female *Aedes aegypti* collected per household survey in 13 spatial repellent (SR) and 13 placebo clusters by study month during 2015 to July 2016 preintervention (cluster-based baseline) (panel A) and January-September 2019 post intervention surveys (panel B). Pre- and post-intervention full-pupal demographic surveys that included collection of adult mosquitos with Prokopack aspirator were carried out over 2–4-month intervals. Shaded areas represent the 95% Confidence Interval around the mean. ..... 36

Supplemental Table S5. Summary of baseline characteristics for spatial repellent (SR) and placebo treatment arms with incidence of Aedes-borne virus (DENV/ZIKV) laboratory-confirmed cases (secondary endpoint) and protective efficacy (PE) estimate from intent to treat (ITT) analyses using Poisson generalized linear regression. No correction for multiple testing was performed for secondary endpoint analyses and, as such, in accordance with CONSORT guidelines, we do not present p-values. ........ 42

### 1. Materials and Methods

#### 1.1 Study site

Our trial was conducted in the Iquitos and Punchana Districts of Iquitos, Peru (**Figure 1**). Detailed descriptions of Iquitos city have been previously published.(1–4)

Briefly, Iquitos is located in the Northern Peruvian Amazon, 120 m above sea level with an approximate population of 400,000 in 2017 and has experienced rapid urbanization over the past 3 decades.(1) The majority of Iquitos residents subsist through small commercial enterprises, transportation (bus and moto-taxis), construction, extractive industries (logging, mining, petroleum), agriculture, and ecotourism. Incomes are modest overall, with 18.2% and 3.0% of the population living in poverty and extreme poverty, respectively.(1) Literacy rates range from 88-92%, and 90-97% of the population has electricity.(5) Evidence of extreme wealth does not exist in Iquitos, and luxury items such as window screening and air conditioners are rarely part of domestic construction.

House construction in Iquitos is relatively homogenous, with a patchwork of homes ranging from wood structures with dirt floors to cement block or brick structures with concrete or ceramic floors (**Figure S1**). Homes tend to be long and narrow, commonly attached to adjacent houses, without enclosed ceilings, and with open eaves between the top of walls. Roofs are usually made of corrugated metal (82-83%).(5)

Dengue virus (DENV) transmission was detected again in Iquitos in 1990 after a 30-year absence, and successive epidemics have occurred with periodic subsequent DENV serotype invasions since then. (6–12) In May 2016, the first human Zika virus (ZIKV) infections were reported and transmission continued through April 2017. There was almost no detection of DENV during that time period of ZIKV transmission.(13)

Routine *Aedes aegypti* control in Iquitos consists of larviciding at ~ 3-month intervals and health education activities utilizing billboards, radio, and TV messages focusing on dengue and its prevention. During the trial period, in response to ZIKV transmission, indoor applications of Malathion using thermal foggers were conducted in all the study clusters during November 2016, February and November-December 2017, and April-May and November-December 2018.

Despite its isolation, Iquitos is a dynamic and economically unstable population center. Many households have multiple families sharing the same living space and a high turnover of residents. In many cases extended families live in the same neighborhood and members move freely between households. There is movement between Iquitos and the capital Lima, and between Iquitos and rural river communities beyond the city.

#### 1.2 Trial design

Our study was a parallel, cluster-randomized controlled clinical trial (cRCT) consisting of 26 clusters each with approximately 140 households conducted over three distinct study phases: Pre-intervention (December 14, 2014 -August 8, 2016), Intervention Deployment (August 9, 2016 -December 31, 2016), and Intervention (January 1, 2017-March 21, 2019) **(Figure 2)**.

The primary endpoint was DENV or ZIKV seroconversion. Secondary endpoints were clinically apparent laboratory confirmed cases of ABV disease and the following entomological parameters for adult female *Aedes aegypti*: 1) indoor abundance, 2) blood-fed rate (proxy for human biting rates) and 3) parity rate (proxy for mosquito age-structure) (**Table S1**).

<u>The primary hypothesis was that </u>SR does not reduce the arbovirus infection rate in qualifying participants compared to placebo. The trial was designed to detect a reduction in hazard rate of 70% or expected PE of 30%.

##### 1.2.1 Cluster delineation, allocation, and blinding

The Peruvian Ministry of Health (MOH) has divided Iquitos into 34 zones used for operational vector control programs. We selected thirteen of these zones to serve as the study area based on the following characteristics: 1) predominantly residential; 2) unaffected by seasonal flooding, which could cause higher rates of human migration and lower rates of DENV transmission; and 3) logistical considerations, proximity to our study field laboratory.

A total of 26 clusters were delineated across the 13 MOH zones. Using a geographic information system (GIS) containing Iquitos house locations, a single block was randomly selected from each of the 13-study area MOH zones using SAS software, Version 9.4 of the SAS System for Windows (SAS Institute Inc., Cary, NC, USA). Clusters were then formed by digitally adding blocks adjacent to the primary block selection until approximately 140 households (1-8 blocks) were captured. A second cluster was delineated in the same zone in a similar manner. The average distance from the nearest cluster was 523 m (range 280-879 m). Clusters were categorized into four strata (designated 1-4) based on anticipated logistical order of intervention deployment (**Figure 1**).

The external statistician serving on the Data Safety Monitoring Board (DSMB) allocated clusters to spatial repellent (SR) or placebo treatment arms, balanced within each stratum, using a random number generator (https://www.random.org). Investigators, research staff, and study participants were blinded as to which clusters received SR versus placebo intervention.

##### 1.2.2 Epidemiological surveys

Two human cohorts were established in the study: 1) all residents of any age group living in households invited to participate in active disease surveillance to identify acute *Aedes*-borne virus (ABV) cases, and 2) a subset of this cohort comprised principally of children $\geq$ 2 – 17 years of age invited to participate in longitudinal monitoring for neutralizing antibodies against DENV and ZIKV. Individuals who turned 18 years of age during baseline were retained in the study for future blood draws during the intervention phase. In addition, subjects with known naïve serostatus from previous studies were purposively recruited to increase the number of susceptible individuals in the cohort. The longitudinal cohort was recruited no earlier than 2 weeks prior to initial deployment of SR intervention within each cluster. Measurement of seroconversion rates was initiated during the Intervention deployment period and continued through November 2018 (**Figure 2)**. Active disease surveillance was initiated in June 2015, with pre-intervention activities conducted on a phased approach across all clusters.

###### *1.2.2.1 Longitudinal cohort for measurement of seroconversion to ABV (Primary Endpoint).*

Longitudinal samples were collected by venipuncture or fingerstick in participant's homes during three sampling periods: August 2016-February 2017 representing Baseline (B), November 2017-February 2018 representing First (F) follow-up survey, and October 2018 -November 2018 representing the Second/Final (S) follow-up survey. Only participants providing $\geq$2 blood samples were screened for all four DENV serotypes and for ZIKV antibodies (**Figure 3**). Samples were transferred to the Iquitos field laboratory in a cooler (4°C), where serum was separated and stored at -80°C until shipping on dry ice to the NAMRU-6 Lima laboratory for testing by a microneutralization enzyme immunoassay (MNT) for serologic reaction against each DENV serotype. Remaining serum was shipped to the University of California, Davis (UC Davis) for MNT testing for ZIKV antibody.

###### *1.2.2.2 Disease surveillance cohort for measurement of laboratory confirmed acute ABVD (Secondary Endpoint).*

Disease cases were identified through door-to-door surveillance with acute samples from those persons meeting inclusion criteria tested by PCR for DENV and ZIKV. Acute- and convalescent-phase blood samples were screened for anti-DENV IgM antibody by IgM-capture ELISA.

##### 1.2.3 Entomological surveys

Longitudinal entomological surveillance was implemented across study clusters starting in December 2014, with 2-4 pre-intervention surveys conducted per cluster up until December 2016 (**Figure 2**). At least one pre-intervention survey was a pupal demographic survey(14–16) where both immature and adult mosquitoes were surveyed. Adult mosquito collections, using Prokopack aspirators(17), commenced two weeks after intervention deployment, and during intervention starting between August 17, 2016 and December 28, 2016 (**Figure 2**). Following removal of the SR and placebo devices from each cluster, two full pupal demographic surveys (post-intervention) were conducted per cluster until September 2019; as with the pre-intervention surveys immature and adult mosquitoes were collected and quantified.

Our rationale for conducting adult mosquito aspirations over using fixed trap methods(18), was to ensure the majority of participating households could be sampled twice each month. Because our trial required sampling up to 2,400 households twice per month and we know that entomological risk needs to be accessed at the individual household level(4), using our experienced staff for aspiration collections was the only way this could be achieved. We used the same collectors, rotating households throughout the study. In previous studies using aspirations we explored inter-operator variation in our statistical models, which was never a significant parameter. Our ability to sample most of the participating households will be critical for accessing the impact in household characteristics, including the number of residents, larval habitat data and SR presence on adult mosquito densities, in our primary analysis described in this manuscript and in future more in-depth studies.

#### 1.3 Procedures

##### 1.3.1 Participants and method of consent

Participant recruitment began on June 30, 2015 (**Figure 2**). Participation in the study included: 1) a household census, 2) febrile surveillance and confirmatory testing with clinical follow-up for identification of ABV disease, 3) children ($\geq$ 2 years to < 18 years) providing three annual blood samples when healthy for detection of seroconversion, 4) routine entomological surveys, and 5) intervention placement and replacement (at 2-week intervals) in households.

Standardized study information sheets, with an overview of the experimental design and study procedures, were provided to all households starting 18 months prior to intervention deployment and throughout the study. Adult participants ($\geq$ 18 years of age), or a parent or guardian of child participants ($\geq$2 years to $\leq$17 years), provided written consent for blood draws. Written assent was also obtained from children $\geq$ 8 years of age. Although the ability for participants < 8 years to give verbal assent varied, staff did not sample children in this age category who said they did not want to participate or exhibited behavior that they did not want to participate, e.g., resisting or crying. Verbal consent was obtained by a head-of-household for intervention deployment and entomological monitoring activities.

Residents could participate in any of the study components described above, where they met inclusion and exclusion criteria, and were allowed to start and stop activities as they requested. Our study teams tracked resident movements closely. Status of all censused participants was assessed and updated at 6-week to quarterly intervals to estimate participant time at risk for each day of the study. During the intervention phase, it was not uncommon for families to vacate a home and another family to move in. As new families moved into the study area, they were recruited into the study.

##### 1.3.2 Intervention application and quality assurance

Our intervention was a passive emanator SR designed and produced by SC Johnson (Racine, WI) and described previously.(19) Briefly, the SR intervention was a clear polyethylene terephthalate (PET) plastic sheet (8.5 x 11 inch) coated on one side during manufacturing with transfluthrin (SR) or solvent (placebo), then folded in half and sealed to prevent active ingredient and solvent release until the product was opened. Dosing specification was 68.75 +/- 3.34375 mg of formula (80% transfluthrin and 20% solvent). Transfluthrin is a registered compound commonly found in commercially available mosquito coils globally based on WHO specifications.(20) The U.S. Environmental Protection Agency (USEPA) has approved transfluthrin products for indoor use within the USA.(21)

Spatial repellent and placebo intervention had identical packaging and were deployed in households by study personnel using a blinded cluster:intervention coding scheme. Removal and replacement of intervention was conducted by study personnel every two weeks throughout the intervention period according to manufacturer's specifications. Two field teams consisting of a coordinator, product placement technicians, and entomological technicians were responsible for managing the logistics of intervention deployment. A cell phone application was developed using CommCare© (Dimagi Inc, MA, USA), a mobile data collection platform, to facilitate deployment and enable monitoring of intervention coverage at the household-level.

The indoor placement of the intervention was designed with the objective to measure PE under indoor use conditions. To ensure a standardized intervention application rate in each household (1 unit per $9m^2$), the dimensions of all enclosed spaces within each house was measured following consent. When a room had an enclosed ceiling (e.g., sleeping area), the application rate was calculated for that room independently. Each intervention was opened immediately before placement, folded into a cylinder, ensuring active ingredient/solvent treated side faced outward towards the interior space of the home, and hung from elastic lines using metal clips (**Figure S1**).

We measured 3 principal metrics of intervention coverage: 1) percentage of houses in each cluster that agreed to intervention application (study participation), 2) percentage of days a house had intervention at any application rate, and 3) percentage of days enrolled houses met application specifications (1 unit/ $9m^2$).

Intervention quality assurance was addressed at the time of manufacturing, as well as, during the trial. As part of the manufacturing process control plan, incoming transfluthrin purity and the transfluthrin content in the formulation were analyzed for every batch by GC (gas chromatography). Spatial repellent and placebo intervention filling weights were measured every hour during production. To verify the amount of transfluthrin in SR products and the absence of transfluthrin in placebo products that had been received in Peru, an independent analysis of a subsample of unused interventions was performed by Ross Laboratories, India.

During the intervention phase and at the end of the trial (March 2019), used, unused and expired emanators were disposed of through a company specializing in chemical waste disposal (Brunner Consultores & Servicio) according to Peruvian disposal regulations.

#### 1.3.3 Laboratory assays

##### *1.3.3.1 Microneutralization Enzyme Immunoassay (MNT).*

We used a validated NAMRU-6 protocol adapted from Vorndam and Beltran,(22) in which 96-well plates (TC-treated) were inoculated with Vero cells at 2 x $10^5$ cells/mL and then incubated at 37ºC, with 5% $CO_2$ for two days or until the cell monolayer was confluent. Serum samples were inactivated at 56°C for 30 min, then serially diluted in triplicate in a two-fold series from 1:20 to 1:1280 on a 96-well plate, along with negative serum and positive hyperimmune mouse ascitic fluid (HMAF) controls. Diluted virus (dilution factor determined by NAMRU-6 validation assays) was mixed with inactivated sera and incubated at 4ºC overnight. A Vero cell suspension at 2x$10^5$cells/mL in 10% Fetal Bovine Serum (FBS) Eagle's Minimum Essential Medium (EMEM) was then added to each well with the serum-virus mixture and incubated at 37ºC with 5% $CO_2$ for five days. After the five days, the cell culture supernatant was discarded and the cells were fixed with ethanol/methanol, washed with phosphate-buffered saline (PBS), blocked with skim milk, then anti-DENV HMAF was added and incubated for 2 h at 37ºC, washed with PBS again and then incubated at room temperature for one hour with 2,2'-azinobis(3-ethylbenzthiazoline-6-sulfonic acid) (ABTS) substrate.

Plates were read using an enzyme-linked immunosorbent assay (ELISA) reader (Microplate Reader Biotek Instruments Inc.) at 405 nm with a 630 nm reference filter. A cut-off value was established for each plate as the numeric value of 50% of the mean optical density (OD) of virus controls and the endpoint titer was the highest serum dilution with mean OD below the cut off value. Endpoint titers were reported as <1/40, 1/80, 1/160, …, ≥1/2560. The MNT for ZIKV was carried out as described for DENV, except that following the five-day incubation period, all plates were gently rinsed with water and stained with 100 µL of crystal violet per well for 1 hour. Afterwards, plates were rinsed again, and each well was examined visually for cytopathic effect. Endpoint titers, which were the dilutions at which cytopathic effect was reduced by ≥ 50% were reported as <1/20, 1/40, 1/80, …, ≥1/1280.

##### *1.3.3.2 Taqman Real-Time PCR for DENV serotypes 1–4.*

Viral RNA was extracted from whole blood and/or serum samples using QIAamp Viral RNA Mini Kits following the manufacture's guidelines. Using a method modified from Johnson et al.(23), primer and probe sets validated on an ABI 7500 real-time PCR platform (Applied Biosystems) were used with TaqMan Fast Virus 1-step RT-PCR master mix (Life Technologies) to detect DENV RNA in serum samples. The protocol consisted of two assays. The first was a real-time multiplex assay that detects DENV-1, DENV-3, and DENV-4 and the second was a real-time singleplex assay that detects DENV-2.

##### *1.3.3.3 Quantitative Real-Time PCR for ZIKV.*

Viral RNA was extracted from serum samples using QIAamp Viral RNA Mini Kit (QIAGEN) according to the manufacturer's instructions. Assays were performed on the ABI 7500 real-time PCR platform (Applied Biosystems) using Superscript III Platinum One-Step qRT-PCR kits (ThermoFisher) in 25 ul final volume reactions comprised of 5 ul template, 12.5 ul of 2x reaction mix, 0.5 ul of 50 mM magnesium sulfate, 0.5 ul of SuperScript III RT/Platinum Taq mix, and 0.5 ul of 10 mM each primers and probe. (24) Reactions were cycled in duplicate under the following amplification conditions: 50°C for 15 min followed by 95°C for 2 min and 40 cycles of 95°C for 15 sec and 58°C for 30 sec.

###### *1.3.3.4 Nested reverse transcription polymerase chain reaction (RT-PCR).*

Human samples with borderline cycle threshold values were confirmed positive or negative for DENV and/or ZIKV using previously extracted RNA and a nested RT-PCR protocol described by Lanciotti et al.(25)

###### *1.3.3.5 IgM capture ELISA.*

Serum was tested for anti-DENV IgM antibody using a NAMRU-6 protocol.(26, 27) Antibody in sera that tested positive was tittered. A 4-fold rise (from acute to convalescent) in titer was considered evidence of seroconversion and recent DENV infection. During the period from May 2016-April 2017 a rise in DENV IgM antibody was presumed to be seroconversion to ZIKV due to a Zika outbreak and undetected DENV transmission throughout Iquitos, including all study clusters(13).

##### 1.3.4 Interpretation of laboratory assay outputs

###### *1.3.4.1 Identification of seroconversions in longitudinal cohort.*

To aid in the identification of seroconversions, a five-character code was used for each participant sample pair where character 1 represented the dengue serostatus for the first sample, character 2 represented the dengue serostatus for the second sample, character 4 represented the Zika serostatus for the first sample, character 5 represented the Zika serostatus for the second sample. Character 3 was always an underscore (_) to separate dengue from Zika serostatus.

For example, an individual participant who had DENV and ZIKV antibody titers of less than 1:40 throughout the study would be characterized as NN_NN for Baseline (B) and First (F) blood sample follow-up surveys and NN_NN for the F to Second/Final (S) follow-up survey interval. The first two letters represented results for DENV with a unique number or letter combination for all possible combinations of DENV results; e.g., 1 would represent a sample with a titer ≥ 1:40 for DENV-1 and < 1:40 for serotype DENV-2, DENV-3, or DENV-4; H would represent a sample with a titer of ≥ 1:40 for all four DENV serotypes. The two letters to the right of the underscore mark represented ZIKV results with N representing samples with ZIKV MNT titer < 1:40 and Z representing a titer ≥ 1:40. Thus the sequence NN_NZ would represent a seroconversion to ZIKV.

All initial coded sequences were verified and reviewed by the trial PI (ACM). The criteria for seroconversion for people with no detectable antibody at baseline was a rise from < 1:40 to > 1:40. For those with detectable baseline antibody a rise from 1:40 to a minimum of an 8-fold rise was required for seroconversion. As an additional quality control measure, R-code was developed by WHE using R 3.6.1 (R Core Team)(28) to score seroconversions using the rules described above. The seroconversion outputs were cross-referenced using both methods with a final agreement of 95%. The final 5% were scored as special cases based on investigator consensus.

###### *1.3.4.2 Criteria for scoring a laboratory confirmed DENV/ZIKV infection*.

Laboratory confirmed DENV or ZIKV infections were scored for individuals with viral RNA detected by PCR or a four-fold rise in IgM titer between acute and convalescent blood samples. Individuals with IgM titers ≥ 1:400 in acute samples without an accompanying convalescent sample, or in both acute and convalescent samples, were considered presumptive infections and considered evidence of a clinically apparent case of dengue or Zika.

##### 1.3.5 Entomological surveys

*1.3.5.1 Collections for measuring indoor adult female Ae. aegypti abundance, blood-fed, and parity rates.*
Pre-intervention entomological surveys started December 17, 2014. Indoor aspiration for adult mosquitoes was conducted at approximate two-week intervals at time of intervention replacement in all consented houses providing access. A final pupal demographic survey and adult collection was conducted in each cluster in association with completion of the intervention phase of the trial.

Up to 30 female *Ae. aegypti* per household per collection date were examined for a blood-meal and parity status, the latter involving examination of the dissected ovaries to determine if the female had or had not developed and/or laid eggs using standard ovarian dissection techniques.(29)

*1.3.5.2 Insecticide resistance assays.*
In September 2017, *Ae. aegypti* eggs were collected from each of the 26 clusters and pooled into four groups (A-D), two groups represented SR clusters and two groups represented placebo clusters. The group allocation was blinded to investigators.  A F3 generation of a genetically diverse *Ae. aegypti* laboratory strain (GDLS), established in 2016 prior to intervention deployment from 10 populations collected throughout Iquitos, including study clusters, served as a control population. We used the pyrethroid susceptible USDA Rockefeller laboratory strain as the reference comparator.

Standard procedures outlined for the Centers for Disease Control (CDC) bottle bioassay were followed.(30) The diagnostic dose of transfluthrin (7.5 ug/bottle) used in our bioassays was guided by CDC staff following independent testing of technical grade transfluthrin using the USDA Rockefeller strain. Three trials were performed for each A-D group and the GDLS population. Each trial consisted of four treatment and three matched reference control replicates (bottles), containing 25 female *Ae. aegypti.* Mortality observations were recorded at 15-min intervals within a 2-h exposure period with rates reported at 30-min used as the diagnostic time for interpreting susceptibility status per the CDC protocol.

##### 1.3.6 Safety monitoring

There are three primary factors that support the transfluthrin intervention used in our trial as safe for indoor use: (1) the product was evaluated for human safety and did not meet the criteria for classification in any hazard class (oral, dermal, inhalation) according to regulation OSHA 29 CFR 1910.1200; (2) products containing transfluthrin, or similar volatile pyrethroids, are already on the market and in use for household mosquito control (coils, electric oils etc.); and (3) there were no reported Significant Adverse Effects (deaths) associated with our spatial repellent intervention during our Data and Safety Monitoring Board safety assessments.  We do recognize that mild reactions to pyrethroid chemicals have been reported and thus our study design included a safety assessment.

To assess safety associated with intervention exposure, we reported both solicited (detected during weekly febrile surveillance and bi-monthly entomology surveys) and unsolicited (calls to study nurses and/or clinician) Adverse Event (AE) complaints from residents living in the study area. To better assess mild or unreported reactions to the intervention, we recorded the reason provided by families that withdrew from the intervention component of the study. A self-administered cell phone survey was implemented May-July 2017 and May-June 2018. As part of this survey, a single question was included asking residents to report any problems they had or were experiencing with the intervention. Data from the survey was used for safety reporting only and was not explored by study staff for trends to avoid unintentional unblinding.

All participants had telephone contact information for the site PI (ACM) and study nurses, which were included on the study consent forms and information sheets. Residents were instructed to call these numbers to report any problems or concerns, especially with the intervention and were asked to report illness. A cell phone application was developed for nurse technicians to report AEs in a standardized format. A project physician evaluated complaints within 24 h of notification and provided a detailed report to the site PI (ACM) who managed further reporting according to local ethical assurance approvals.

We recorded and reported detected deaths in the study clusters as Severe Adverse Events (SAEs), although the rate at which our study personnel received information reported to them was variable. In January 2018, a cell phone application was deployed to monitor self-reported hospital stays by cohort members to enhance SAE data detection.

Safety data reported from the site, which included AEs reported directly to field staff (visits/phone), deaths, hospitalizations, withdrawals (reasons reported, if provided), and results from the self-administered questionnaire was reviewed by the DSMB on a quarterly basis from June 2017 to February 2019. The DSMB conducted a safety assessment in February 2019, from a cumulative safety report, representing the final safety assessment for the trial.

#### 1.4 Data Management

Data collection was carried out using a combination of: 1) standardized paper forms adapted from a series of dengue cohort studies conducted between 1999-2015 in Iquitos, Peru by the UC Davis/ NAMRU-6 team(9, 31–33) and 2) digital forms using a mobile Android device-based survey platform built with CommCare© (Dimagi Inc, MA, USA). With Commcare©, form data were collected offline during follow-up visits to study households and uploaded to the secure CommCare© server daily upon return to our Iquitos research facility.

Both sources of data were managed in an integrated data management system (DMS) that was upgraded using Django(34), a Python web-framework that followed a model-view-template architecture. The DMS included a secure PostgreSQL database, linked to our Django web interface we developed and built with open-source software and three 64-bit database servers (1 TB storage 8 GB RAM). Servers were housed in our two laboratory facilities in Iquitos and in a secure server facility on the UC Davis campus. High speed communication between the servers allowed for a constant data flow, maintaining the same data on all three servers. This allowed for high-speed access to data at UC Davis for team members based in the U.S. and significantly increased security due to the redundancy of the offsite data backup. Data access and sharing was mediated through our secure website and limited to authorized users.

An updated geographic information system (GIS) was built from shapefiles previously managed in ArcGIS™ (ESRI, Redlands, CA, USA) using the PostGIS database extension(35) in the PostgreSQL database extension. These spatial data were visualized and updated with interactive tools in the open-source software QGIS.(36) Every household lot located in the 26 study clusters was assigned a unique code. All serological, virological, participant status and entomological survey data were linked to this unique code.

We used the mobile Android device-based survey platform to record surveillance visits, update participant status, and record deployment and replacement of the intervention. We calculated the person-days for each censused participant contributing to the surveillance denominator and intervention coverage. This was recorded in a status data table that provided start and stop dates for alternating periods of participant presence and absence under surveillance in the study area. The status of all participants was verified by field staff at 6-8-week intervals or updated if changes in status were reported by study participants. Participation in individual components of the trial were managed through a consent table facilitating linkage of multiple data sources.

Field staff had access to the DMS via the website for viewing only, to check and verify participant status. Other team members had access for data entry only, and/or full editing privileges depending on their role in the study. This DMS allowed the generation of real-time, customized paper census and longitudinal data collection forms based on existing data in the system. For example, the census list included the current participant status (active or lost to follow up) and verification of consent so staff could verify or update status or consent (e.g., add an assent form due to an age change) during longitudinal blood draw visits.

Quality assurance systems were built into the DMS, by response constraints, and a series of stored queries were established to identify missing and/or incomplete data (e.g., cluster code that did not match address). Data integrity was conducted in real-time by senior team members permanently stationed in Iquitos (ACM, WHE), who cross-checked DMS information with the source document (paper or CommCare© database). After cleaning, data were verified, and the dataset was locked for analyses. Locked data files were exported from our custom database in .csv format and analyzed using SAS statistical analysis software version 9.3 (SAS Institute Inc., Cary NC, USA) and R versions 3.4.3 and 3.6.1, as per the Statistical Analysis Plan (**provided as Supplemental document; Section 1.8.2**).

#### 1.5 Statistical Analyses

##### 1.5.1 Intent-to-treat and per protocol general considerations.

For all endpoint analyses, the primary analysis was an intent to treat (ITT). In this, every qualifying participant that lived in a SR or placebo cluster was considered for inclusion, independent of their participation in the intervention. An ITT analysis can more accurately capture the real-world effectiveness of an intervention given imperfect enrollment and imperfect application of the intervention.

A second analysis was conducted for each endpoint that focused only on those individuals who participated in the intervention as intended; i.e., per protocol (PP). Here, the per protocol analyses are based on adherence to the intervention, which can be measured in terms of the number of days when the individual's house had intervention and the number of intervention units. Each house was measured at time of enrollment to calculate the number of intervention units required per manufacturer specifications (1 unit/9m2) and, during the trial, at the time of intervention replacement, the number of actual intervention units deployed in households was counted.

The Statistical Analysis Plan (SAP) did not specify an exact definition of “per protocol” intervention coverage rate at the household-level to apply in analyses. Ideally, to test a vector control intervention the goal is to maximize intervention coverage and user compliance. Inherently, however, the research team has limited control over study participants and in practice can only monitor these parameters carefully.(37)

During our trial, we estimated the location days that participants were under adequate coverage (intervention units in a house meeting manufacturer specification of 1 unit/9$m^2$) to be 73-75% (**see Section 2.6 and Supplementary Table S2**). For this reason, and based on our prior experience with estimates of high coverage under assurances of quality control and user compliance,(37) we selected 75% as proportion of location days covered between longitudinal blood samples for the per-protocol threshold and conducted four unique analyses based on that rate:

- First, we only included individuals who had at least the adequate number of intervention units specified by manufacturer in their house for $\geq$75% of the days between sequential blood sampling (designated PP-1);
- Second, we evaluated the sensitivity of our conclusions to the “75%” threshold by varying it from 5 to 100% (designated PP-2);
- Third, we included individuals that had any number of interventions in their house for $\geq$75% of the days between sequential blood sampling (designated PP-3);
- Fourth, we evaluated robustness of results using the “75%” threshold in PP-3 by varying it from 0 to 100% (designated PP-4).

In addition to participants entering the trial throughout the trial’s duration, different clusters entered the trial in a staggered manner throughout the second half of 2016 (**Figure 2**). Two to 4 pre-intervention entomological surveys were conducted within each cluster between December 2014 and December 2016. These pre-intervention surveys were not carried out at the same consistent 15-day intervals as the post-intervention adult mosquito collections. They do, however, provide temporally stratified mosquito abundance, blood-fed and parity rate estimates before and after transfluthrin was first present in the cluster~~r.~~ Based on this, analyses of entomological parameters were conducted employing two approaches to specifying a ‘baseline’ period. First, the first specified measurements made throughout 2016 as baseline, with post-2016 measurements as ‘post-baseline’ (designated 2016 Baseline). Second, specifying measurements were made between December 2014 up to the first date of intervention deployment in 2016 in each cluster as baseline, with all measurements following that date as ‘post-baseline’ for that cluster, even for houses that did not enroll until 2017 or later (designated cluster-specific Baseline) **(Table S1)**.

Because we used a survival analysis with a proportional hazards model and exponential distribution assumption for the baseline hazard instead of a mixed effects logistic regression model that was originally proposed in the SAP, we also carried out the originally proposed statistical analysis. We also present a more appropriate logistic regression model including only individuals whose paired blood samples were separated by 9-15 months, representing closeness to proposed annual blood sampling in the study protocol.

No correction for multiple testing was performed for secondary endpoint analyses. As such, in accordance with CONSORT guidelines, we do not present p-values for these outcomes.

##### 1.5.2 Primary Endpoint – Seroconversion to ABV infection

###### *1.5.2.1 Primary analysis – ITT survival analysis.*

The primary hypothesis on PE against arbovirus infection in qualifying participants was tested by comparing the hazard rates of seroconversion to DENV and/or ZIKV (ABVs circulating in Iquitos during the study period) in qualifying participants between SR and placebo groups. Given the variation in the time interval between paired samples tested by MNT among the longitudinal cohort participants, a survival analysis using a proportional hazards model with an exponential distribution assumption for the baseline hazard (i.e., constant baseline hazard through time) was used to account for variation in the duration between blood draws by individual.

If $h(t_{ij}|\boldsymbol{x}_{ij})$ is the hazard rate of the $j^{th}$ individual in the $i^{th}$ cluster with covariate values $\boldsymbol{x}_{ij}$ then this individual's hazard rate of an arbovirus infection can be written as:

$$h(t_{ij}|x_{ij}) = h_0(t_{ij}) \cdot \exp(\boldsymbol{\beta}^T \boldsymbol{x}_{ij} + W_i)$$

Where $W_i \sim N(0, \sigma_c^2)$ is the random effect of the $i^{th}$ cluster. Covariates included are age, sex, and treatment status (SR or placebo). The protective efficacy (PE) was estimated as $PE = \left(1 - \exp(\hat{\beta})\right) \times 100\%$, where $\hat{\beta}$ is the estimated regression coefficient for the intervention group and $\exp(\hat{\beta})$ is the estimated hazard ratio (HR) between SR and placebo. The null hypothesis of PE = 0% is equivalent to $\beta = 0$, which is tested by Wald's test $z = \beta/s$, where $s$ is the estimated standard error of $\hat{\beta}$, at the 1-sided significance level of 5%.

The Kaplan-Meier curves for SR and placebo on ABV seroconversion of qualifying participants by cluster were plotted. The baseline characteristics of the enrolled subjects were summarized. The hazard ratio between SR and placebo was estimated from the model, along with the 1-sided 95% CI's upper bound.

###### *1.5.2.2 Secondary analysis – ITT mixed effects logistic regression (original SAP).*

The original analysis was a mixed effects regression with fixed effects for treatment arm, as well as, individual level characteristics (here, age and sex) and a random effect for cluster. The original intent was to only test children, so we subsetted "qualifying participants" down to those under the age of 18.

The model formulation is:

$$\log\left(\frac{\pi_{ijk}}{1 - \pi_{ijk}}\right) = \alpha + \beta_t * 1_{[i=1]} + \sum_l \gamma_l z_{ijkl} + u_{ij}$$

Where:

- $\pi_{ijk}$ is the true probability of seroconversion for the *k*th individual in the *j*th cluster in the *i*th treatment arm,
- $\beta_t$ is the intervention effect,
- $1_{[i=1]}$ is an indicator function equal to 1 if the individual is in the treatment arm,
- $z_{ijkl}$ is the value of the lth covariate for the *k*th individual,
- $\gamma_l$ is the effect of the *l*th covariate, and
- $u_{ij}$ is the random effect corresponding to the *j*th cluster in the *i*th treatment arm.

###### *1.5.2.3 Secondary analysis – ITT mixed effects logistic regression (adjusted from original SAP).*

In an approach to achieve an analytical dataset reflecting the intention of the original trial protocol, that was to only analyze individuals whose duration between blood sampling was close to 1 year, we arbitrarily chose durations between 9 and 15 months as "close". The model formulation was then identical to that of the original SAP's ITT analysis.

###### *1.5.2.4 Secondary analysis – Per protocol (PP) analyses*

###### 1.5.2.4.1 Secondary analysis – PP survival analysis (PP-1)

As specified in Section 1.5.1 (**see SI section 2.5.1.4 and Table S2**), we selected >75% as the threshold for adequate coverage rate of intervention (at manufacturer specifications for house-level application rate) for the duration between sequential blood sampling based on actual trial data and previous experience of coverage estimates with other interventions in Iquitos.(37) After subsetting the data, we retained 876 individuals and ran the same survival analysis as described above for the primary analysis of the primary endpoint.

###### 1.5.2.4.2 Secondary analysis – PP survival analysis (PP-2)

We assessed the robustness of the significance of our PP-1 analyses using the 75% coverage threshold by repeating the regression for the duration between blood sampling ranging between 5% and 100%.

###### 1.5.2.4.3 Secondary analysis – PP survival analysis (PP-3)

In the PP-1 analysis described above, we subsetted by the individuals that had at least as many intervention units as was estimated to be adequate to meet manufacturer's specifications when their house was enrolled. Here we reconsidered the PP analysis, with a more liberal inclusion criteria of participants that had any amount of intervention in their house for >75% of the duration between their sequential blood sampling.

###### 1.5.2.4.4 Secondary analysis – PP survival analysis (PP-4)

We repeated the analysis in PP-2 here for "any SR intervention coverage" in the household for any amount of time (0-100%) between sequential blood sampling for completeness.

##### 1.5.3 Secondary Endpoint – PCR / ELISA confirmed DENV and ZIKV cases–ITT fixed effect Poisson generalized linear model

There was a total of 96 DENV/ZIKV cases detected through PCR/ELISA. As such, the only analysis conducted was an ITT analysis. A Poisson regression was used to assess the impact of SR intervention on the number of acute DENV and ZIKV cases confirmed by PCR/ELISA, with an offset for the number of participant-days that participants spent in each cluster. Originally, there was an intent to add a random effect by cluster, but there were too few acute DENV and ZIKV cases detected by PCR or ELISA.

##### 1.5.4 Secondary Endpoint – Indoor adult female *Aedes aegypti* abundance

###### *1.5.4.1 Primary analysis – ITT mixed effects difference-in-difference (DID) analysis: '2016 baseline'.*

A mixed effects difference-in-difference (DID) Poisson regression was used to assess the impact of SR intervention on indoor adult female *Aedes aegypti* abundance, with factor-level monthly covariates to account for seasonality. Letting $y_{ijk}$ being the number of adult female *Aedes aegypti* collected at time $t_{ijk}$ at location $j$ in cluster $i$, we create a dummy variable $s_{ijk}$ that is 0 if $t_{ijk}$ is during 2016 and 1 otherwise. Further, we created a second summary variable, $d_{ijk}$, that is 1 if $s_{ijk}$ is 1 and cluster $i$ is in the treatment arm. Then, letting $m_{t_{ijk}}$ being the month of the year associated with $t_{ijk}$, we fit a mixed effects Poisson regression:

$$\log y_{ijk} = \beta_0 + \beta_s \cdot s_{ijk} + \beta_t \cdot treatment + \beta_{DID} \cdot d_{ijk} + \beta_{m_{t_{ijk}}} + W_i + \epsilon_{ijk}$$

where $W_i$ is a cluster-level random effect by cluster.

###### *1.5.4.2 Secondary analysis – PP (PP-1) mixed effects DID analysis: '2016 baseline'.*

To begin to assess the PP impact of SR intervention on indoor adult female *Aedes aegypti* abundance, we repeated the above analysis subsetting to houses that had at least as many intervention units as was specified by the manufacturer when the house was first assessed during enrollment. We repeated the analysis for houses that had any amount of intervention (i.e., PP-3) and found comparable results (results not shown).

We used the same model formulation as above. Letting $y_{ijk}$ being the number of indoor adult female *Aedes aegypti* collected at time $t_{ijk}$ at location $j$ in cluster $i$, we created a dummy variable $s_{ijk}$ that is 0 if $t_{ijk}$ is during 2016 and 1 otherwise. Further, we created a second summary variable, $d_{ijk}$, that is 1 if $s_{ijk}$ is 1 and cluster $i$ is in the treatment arm. Then, letting $m_{t_{ijk}}$ being the month of the year associated with $t_{ijk}$, we fit a mixed effect Poisson regression:

$$\log y_{ijk} = \beta_0 + \beta_s \cdot s_{ijk} + \beta_t \cdot treatment + \beta_{DID} \cdot d_{ijk} + \beta_{m_{t_{ijk}}} + W_i + \epsilon_{ijk}$$

where $W_i$ is a cluster-level random effect by cluster.

###### *1.5.4.3 Secondary analysis – ITT mixed effects DID analysis: 'cluster-specific baseline'.*

We conducted the same DID analysis as in 1.5.4.1 above specifying measurements from all collections that occurred after the date a cluster first received intervention as post-baseline.

###### *1.5.4.4 Secondary analysis - PP (PP-1) mixed effects DID analysis: 'cluster-specific baseline'.*

We conducted the same DID analysis as in 1.5.4.2 above specifying measurements from all collections that occurred after the date a cluster first received intervention as post-baseline.

##### 1.5.5 Secondary endpoint – Blood-fed adult female *Aedes aegypti* abundance

###### *1.5.5.1 Primary analysis – ITT mixed effects difference-in-difference (DID) analysis: '2016 baseline'.*

A mixed effects difference-in-difference (DID) Poisson regression was used to assess the impact of SR intervention on the frequency at which blood-fed adult female *Aedes aegypti* were collected, with factor-level monthly covariates to account for seasonality, and an offset for the number of adult female *Aedes aegypti* that were assessed for blood-fed status.

Letting $y_{ijk}$ be the number of blood-fed *Aedes aegypti* collected at time $t_{ijk}$ at location $j$ in cluster $i$, we created a dummy variable $s_{ijk}$ that is 0 if $t_{ijk}$ is during 2016 and 1 otherwise. Further, we created a second summary variable, $d_{ijk}$, that is 1 if $s_{ijk}$ is 1 and cluster $i$ is in the treatment arm and is 0 otherwise. Then, letting $z_{ijk}$ be the number of adult female *Aedes aegypti* that were assessed for blood-fed status at the time of collection and $m_{t_{ijk}}$ be the month of the year associated with $t_{ijk}$, we fit a mixed effects Poisson regression:

$$\log y_{ijk} = offset(z_{ijk}) + \beta_0 + \beta_s \cdot s_{ijk} + \beta_t \cdot treatment + \beta_{DID} \cdot d_{ijk} + \beta_{m_{t_{ijk}}} + W_i + \varepsilon_{ijk}$$

where $W_i$ is the cluster-level random effect.

##### *1.5.5.2 Secondary analysis – PP (PP-1) mixed effects DID analysis: '2016 baseline'.*
To begin to assess the PP impact of SR intervention on the frequency at which blood-fed adult female *Aedes aegypti* were collected, we repeated the above analysis subsetting to houses that had at least as many intervention units as was specified by the manufacturer when the house was first assessed during enrollment. We repeated the analysis for houses that had any amount of intervention (i.e., PP-3) and found comparable results (results not shown).

We used the same model formulation as above. Letting $y_{ijk}$ being the frequency at which blood-fed adult female *Aedes aegypti* were collected at time $t_{ijk}$ at location $j$ in cluster $i$, we created a dummy variable $s_{ijk}$ that is 0 if $t_{ijk}$ is during 2016 and 1 otherwise. Further, we created a second summary variable, $d_{ijk}$, that is 1 if $s_{ijk}$ is 1 and cluster $i$ is in the treatment arm. Then, letting $m_{t_{ijk}}$ being the month of the year associated with $t_{ijk}$, we fit a mixed effect Poisson regression:

$$\log y_{ijk} = \beta_0 + \beta_s \cdot s_{ijk} + \beta_t \cdot treatment + \beta_{DID} \cdot d_{ijk} + \beta_{m_{t_{ijk}}} + W_i + \epsilon_{ijk}$$

where $W_i$ is a cluster-level random effect by cluster.

##### *1.5.5.3 Secondary analysis – ITT mixed effects DID analysis: 'cluster-specific baseline'.*
Following the approach taken in 1.5.5.1 above, for the frequency at which blood-fed adult female *Aedes aegypti* were collected*,* we conducted the same DID analysis here specifying all collections that occurred after the date a cluster first received intervention as post-baseline.

##### *1.5.5.4 Secondary analysis – PP (PP-1) mixed effects DID analysis: 'cluster-specific baseline'.*
Following the approach taken in 1.5.5.2 above, or the frequency at which blood-fed adult female *Aedes aegypti* were collected, we conducted the same DID analysis here specifying all collections that occurred after the date a cluster first received intervention as post-baseline.

#### 1.5.6 Secondary endpoint – Adult female *Aedes aegypti* parity rate

##### *1.5.6.1 Primary analysis – ITT mixed effects difference-in-difference (DID) analysis: '2016 baseline'.*
A mixed effects difference-in-difference (DID) Poisson regression was used to assess the impact of SR intervention on adult female *Aedes aegypti* parity rate, with factor-level monthly covariates to account for seasonality and an offset for the number of mosquitoes assessed for parity by collection.
Letting $z_{ijk}$ being the number of parous adult female *Aedes aegypti* collected at time $t_{ijk}$ at location $j$ in cluster $i$, we created a dummy variable $s_{ijk}$ that is 0 if $t_{ijk}$ is during baseline and 1 otherwise. Further, we created a second summary variable, $d_{ijk}$, that is 1 if $s_{ijk}$ is 1 and cluster $i$ is in the treatment arm. Then, letting $m_{t_{ijk}}$ denote the month of the year associated with $t_{ijk}$ and $y_{ijk}$ denote the number of mosquitoes for which parity was assessed, we fit a mixed effect Poisson regression:

$$\log z_{ijk} = \beta_0 + \beta_s \cdot s_{ijk} + \beta_t \cdot treatment + \beta_{DID} \cdot d_{ijk} + \beta_{m_{t_{ijk}}} + W_i + \text{offset}\left(y_{ijk}\right) + \epsilon_{ijk}$$

where $W_i$ is a cluster-level random effect by cluster.

##### *1.5.6.2 Secondary analysis – PP (PP-1) mixed effects DID analysis: '2016 baseline'.*
As with mosquito abundance, we repeated the above ITT analysis for parity subsetting to houses that had at least as many intervention units as was specified by the manufacturer when the house was first

assessed at enrollment. We repeated the analysis for houses that had any intervention units (i.e., PP-3) and found comparable results (results not shown).

###### *1.5.6.3 Secondary analysis – ITT mixed effects DID analysis: 'cluster-specific baseline'.*

Again, following the approach taken in 1.5.6.1 for adult female *Aedes aegypti* parity rate, we considered a different baseline for each cluster based on when that cluster was first entered into the trial.

###### *1.5.6.4 Secondary analysis – PP (PP-1) mixed effects DID analysis: 'cluster-specific baseline'.*

Again, following the approach taken in 1.5.6.2 for adult female *Aedes aegypti* parity rate, we considered a different baseline for each cluster based on when that cluster was first entered into the trial.

### 2. Results and Interpretation

#### 2.1 Enrollment Statistics

Over the duration of the study (June 2015 to December 2018), 18,240 Iquitos residents in 2,438 houses were censused and enrolled in the disease surveillance cohort. Of these, a total of 12,852 residents were enrolled during the pre-intervention period, of which 29% were 2-17 years of age, the target age range for recruitment into the longitudinal cohort to measure seroconversion to DENV/ZIKV (primary endpoint). The remaining residents were < 2 years (4%) or $\geq$ 18 years (67%).

Of the 18,240 persons enrolled in the trial, 16,683 persons (8,235 in the SR arm and 8,448 in placebo arm) contributed person-time to the final analyses (**Figure 3 and Figure S2)**. Overall, LTFU was lower in the SR (20.6%) than placebo (23.5%) arm ($\chi2=20.4$, df=1 $p<0.0001$). Of the 4,892 individuals who were LTFU before the end of the study, 4.4% died, 89.8% left the study because of travel or change of residence, and 5.1% asked to dropout. There was a total of 1,557 persons (8.5%) whom did not contribute person-time due to LTFU before intervention implementation in their cluster (before 2017). For those individuals enrolled during the intervention phase of the trial, a total of 3,689 (22%) voluntarily withdrew prior to the end of the study (March 2019). Of these, a total of 1,700 and 1,989 resided in SR and placebo clusters, respectively.

Of the 2,215 participants in the longitudinal cohort providing blood samples to identify seroconversions to DENV/ZIKV, LTFU was 17.4% and 15.3% in the SR and placebo arms, respectively ($\chi2=2.01$, df=1 $p=0.1558$). Of the 367 participants that only provided a single blood sample for MNT testing, and thus labeled LTFU, 65% left the study because of travel or change of residence, 16.6% because their parents refused to let them continue, 17.1% because the child chose not to give another sample, and 1.3% because the participant could not be found, and 1 participant had insufficient sample volume, precluding MNT testing on paired sample. There were 162 samples that had insufficient serum volume for ZIKV testing.

#### 2.2 Participant Household Characteristics

The median footprint of study houses was 87$m^2$ (IQR: 61.5-115$m^2$, range 7-464$m^2$). There was a median of 6 rooms (IQR: 5-8, range 1,55) and 12 emanators (IQR-8-12, range 1,50) per household.

There were no statistically significant differences in the number of homes with electricity, having daily access to tap water, and access to indoor plumbing (> 99% in both arms of the trial). Residents using firewood for cooking was 30% and 32% in SR and placebo arms, respectively ($\chi2^2=0.9581$, df=1, p=0.3277).  The proportion of homes with open eaves was 73% and 76% in the SR and placebo arm, respectively ($\chi2$=2.74, df=1, p=0.0978).  Differences were observed among the housing materials used. The proportion of houses with concrete walls was nearly identical with 75% from the SR and 76% from the placebo arm ($\chi2$=0.56, df=1, p=0.4543), but houses with some brick walls were lower in the SR (16%) than placebo (24%) arm ($\chi2$=4.554, df=1, p=0.0328), and wood walls were rare overall, but slightly higher in SR (7.5%) than placebo (5.4%) arm ($\chi2$=27.34, df=1, p<0.0001).

The use of insecticides by residents was less than 5% of the households across all clusters, with 79% of reported use being hand-held pyrethroid spray cans, and 13% contracting someone to fumigate the house. There were only three households (all from placebo clusters) reporting the use of transfluthrin-based mosquito coils.

#### 2.3 Intervention Coverage

A summary of intervention coverage metrics is given in **Table S2 and Figure S3**. Of the houses registered in the GIS database the mean percentage of households per cluster (SR and placebo) that had intervention deployed at some point during the study period was of 44.2 % (SD = 9.7%). A non-significantly greater percentage of houses participated in SR versus placebo clusters (47.0% v 41.4%, p-value = 0.223). In those households consenting to receive intervention (SR or placebo), the mean percentage of days these were covered by an intervention at the cluster-level was 81.6% (SD = 3.9), with slightly higher coverage in households assigned to SR intervention (82.9%) compared to households within the placebo arm (80.3%), albeit insignificant (p-value = 0.153). For all enrolled households, the mean percentage of days with an adequate intervention application rate (1 product per 9 $m^2$) was 73.6% (SD = 9.1), with similar rates between SR and placebo clusters (72.7% versus 74.5%, p-value > 0.999).

#### 2.4 Intervention Quality Assurance

Data from the intervention manufacturing process control plan indicated no deviations from specifications, including all incoming inspections of transfluthrin purity, transfluthrin content in the formulation, and SR and placebo product filling weights for product used in the trial. The 64 samples of unused products randomly pulled from stock in Iquitos assigned to SR and placebo clusters had manufacturing dates ranging from July 2016 to March 2017 (4-month- to 12-month-old samples at the time of analysis). The average transfluthrin quantity for all sampled SR products (n=52) was 54.45 ± 1.15 mg, which was within the specified range (55.0 mg ± 2.75 mg). None of the sampled placebo products (n=12) contained transfluthrin.

#### 2.5 Epidemiological Indicators

##### 2.5.1. Primary Endpoint – Seroconversion to ABV infection

###### *2.5.1.1 Primary analysis –ITT survival analysis.*

Baseline characteristics for the covariates included in the survival analysis were balanced between SR and placebo arms **(Table 1)**. There were slightly fewer individuals residing in the SR clusters (both total and qualifying participants) with baseline ABV susceptibility also slightly lower on average.

Results for the primary analysis of the primary endpoint for PE of SR intervention against arbovirus infection in qualifying participants is presented in **Table 2 and Table S3**. The hazard ratio is 65.9% with an upper bound on the 1-sided 95% CI of 93.1%. This translates into a 34.1% reduction in the hazard risk by SR compared with placebo with the lower bound of the 1-sided 95% CI of 6.9%. The 34.1% PE estimate was statistically significant at the 5% level (Test statistic: $z = 1.98$, 1-sided p-value = $2.36\%$).

The Kaplan-Meier curves for SR and placebo clusters on arbovirus infection for qualifying participants by cluster show considerable between-cluster variation as evidenced by the wide spread of survival curves (**Figure S4).** As two examples, there were no arbovirus infections in qualifying participants in one placebo cluster (Cluster 7.2, which only had 18 qualifying participants). Conversely, in Cluster 8.2, almost half of those qualifying participants whose duration between tests exceeded 15 months became infected (5/11). The duration between tests varied by participant and across clusters (**Figure S5**), resulting in some Kaplan-Meier curves being estimated beyond 2 years. In many of those clusters, the only participants who went 2+ years between blood sampling were universally found to have had an arboviral infection.

**Table S3** presents the effect of each of the baseline covariates included in the statistical model on the hazard of arbovirus infection in qualifying participants. In summary, one covariate (age) had statistically significant effects on the hazard of arbovirus infection in qualifying participants, the hazard rate increases by 4.6% for every one-year increase in age. The other covariate (sex) did not have a statistically significant effect on the hazard of arbovirus infection in qualifying participants, the hazard rate decreases by 4.4% in males relative to females.

###### *2.5.1.2 Secondary analysis – ITT mixed effects logistic regression (original SAP).*

Ignoring any impact of the differential participation duration across qualifying participants, secondary analysis of the primary endpoint using an ITT mixed effects logistic regression model proposed in the original statistical analysis plan estimated a PE of 45.3%. Details, including covariate effects, are given in **Table S3** where the estimated odds ratio is 0.547 (1-sided 95% CI: $(-\infty, 0.883)$), with age having a significant impact on seroconversion.

###### *2.5.1.3 Secondary analysis – ITT mixed effects logistic regression (adjusted from original SAP).*

The estimated PE in those individuals who went “approximately” one year (9-15 months) between blood sampling (durations between 9 and 15 mo) was 42.3% where the estimated odds ratio is 0.577 (1-sided 95% CI: $(-\infty, 0.879)$) and age being the only covariate with significant impact on seroconversion (**Table S3**).

###### *2.5.1.4 Secondary analysis – Per protocol (PP) analysis.*

###### *2.5.1.4.1 Secondary analysis – PP survival analysis (PP-1).*

**Table S4** presents PE estimates and effect of the baseline covariates included in the statistical model on the hazard of arbovirus infection in qualifying participants based on our ≥75% threshold for adequate household coverage rate (at manufacturer’s specification) for the duration between blood sampling. The estimated PE was 37.4% with a corresponding hazard rate ratio of 0.626 (1-sided 95% CI: $(-\infty, 0.806)$), similar to the PE estimates from the primary analysis.

###### *2.5.1.4.2. Secondary analysis – PP survival analysis (PP-2).*

Having repeated the regression for coverage rates ranging between 5% and 100% to assess the robustness of the significance of our PP-1 analyses, we include the plot of PE with 90% two-sided confidence interval (**Figure S6**). Mean results from the primary analysis are included as a grey dashed horizontal line for reference. Results are relatively robust to the level of coverage required for “per protocol" analysis and only become substantially different when the requirements for inclusion exceed 95% coverage, and the number of qualifying participants drops considerably. Although we did not assess it, this may be due to a large indirect effect(38) of the intervention. A large indirect effect would indicate that this intervention would scale well to larger communities as only a fraction of the residents would need to actively participate. *A priori*, we expected the 5% test to closely resemble the results of the primary analysis. Because only 12 individuals met the criteria of 100% SR coverage for the entire trial with 2 seroconversions, one in each treatment arm, we expected the 100% test to not have a statistically significant outcome.

###### *2.5.1.4.3 Secondary analysis – PP survival analysis (PP-3).*

**Table S4** presents PE estimates and effect of the baseline covariates included in the statistical model on the hazard of arbovirus infection in qualifying participants that had any amount of intervention in their house for ≥75% of the duration between their sequential blood sampling. The estimated PE was 37.5% with a corresponding hazard rate ratio of 0.625 (1-sided 95% CI: $(-\infty, 0.831)$), similar to the PE estimates from the primary analysis.

###### *2.5.1.4.4 Secondary analysis – PP survival analysis (PP-4).*

Having repeated the regression for coverage rates ranging between 0 and 100% to assess the robustness of the significance of our PP-3 analyses, we included a plot of the PE with 90% two-sided confidence interval (**Figure S7**). Mean results from the primary analysis are included as a grey dashed horizontal line for reference. Similar to outcomes from PP-2, results were relatively robust to the level of coverage required for "per protocol" analysis and only become substantially different when the requirements for inclusion exceed 95% coverage and the number of qualifying participants drops considerably. As with the PP-2 analysis, because only 25 individuals had some amount of intervention units hanging in their house for 100% of the time, with 3 seroconversions (1 in the SR arm, 2 in the placebo arm), we expected the 100% test to not have a statistically significant outcome.

#### 2.5.2 Secondary endpoint – Laboratory confirmed DENV/ZIKV cases

##### 2.5.2.1 Primary analysis – ITT fixed effect Poisson generalized linear model.

Baseline characteristics of covariates included in the analysis of PCR/ELISA confirmed DENV and ZIKV cases were balanced between SR and placebo arms **(Table S5)**. Results from the ITT fixed effect Poisson generalized linear model indicate the rate ratio is 1.144 with an upper bound on the 1-sided 95% CI of 1.601. This translates into a 14.4% increase in the rate of PCR/ELISA confirmed arbovirus infections by SR intervention compared with placebo with the lower bound of the 1-sided 95% CI of –60.1%. The 14.4% rate reduction was in the opposite direction from the alternative hypothesis of a reduction in incidence and was not statistically significant at the 5% level (Test statistic: $z = -0.975$). It is important to note that there was a total of only 96 PCR/ELISA confirmed DENV/ZIKV cases in 10,802,926 participant days across all 26 study clusters. These positive tests were close to evenly split, with 51 originating from clusters allocated to receive SR intervention and 45 from placebo clusters.

There were no covariates used in this analysis.

#### 2.6 Entomological Indicators

Estimates of key entomological parameters depended on data from surveys designated as baseline and post-baseline.  The 2016 baseline approach summarized in **Table S6**, utilized surveys conducted within 2 weeks of initial intervention deployment as well as post-deployment surveys conducted throughout 2016. Given rolling enrollment of the deployment, some of these "baseline" collections were, in fact collected from houses already using the intervention, whereas others were from houses (or clusters) where the intervention had not been deployed yet. In contrast, the cluster-specific baseline approach (data summarized in **Table S7**), focuses the baseline surveys on collections made before any house in the cluster had the intervention deployed. That said, for the cluster-specific baseline approach, "post-baseline" for every house in a cluster is set on the first day the intervention was deployed in a cluster. **Figure S3** summarizes intervention activities including deployment over the course of the trial.

##### 2.6.1 Secondary endpoints: adult female *Aedes aegypti* abundance, blood-fed *Aedes aegypti* abundance, and *Aedes aegypti* parous rate from ITT and PP mixed effects DID analysis (2016 Baseline).

The number of 2016 baseline entomology collections (household surveys) per cluster, and female *Ae. aegypti* evaluated for blood-fed and parity status was lower in the SR compared to placebo arm. Estimates, however, of the number of female *Ae. aegypti* and those blood-fed per household collection as well as parous rate (proportion of older females) were balanced between treatment arms at baseline **(Table S6).** In the post-baseline period, the number of household collections and female *Ae. aegypti* evaluated for parity were higher in the SR than placebo arm, whereas the number of *Ae. aegypti* evaluated for blood-fed status was lower in the SR arm **(Table S6).**

Female *Ae. aegypti* abundance decreased overall in both the SR and placebo arms, but significantly more in the SR arm as with rate reductions in the ITT and PP mixed effects difference-in difference (DID) models of 28.6% and 26.3%, respectively **(Table S8)**.  Pre- and post-intervention entomology surveys indicated similar Ae. aegypti densities across treatment arms (**Figure S8**). Blood fed *Ae. aegypti* abundance increased overall between baseline and post-baseline surveys, but the increase was less in the SR clusters. This translates into a 12.4% reduction in blood-fed females in the SR compared to placebo arm for both ITT and PP models **(Table S8)**. Estimated parity rates showed similar rates between baseline and post-baseline periods and between treatments, illustrated by confidence intervals containing zero **(Table S8)**.

###### *2.6.1.1 Effects of covariates included in statistical models (Seasonality).*

To control for temporal variation, each month was included as a covariate in the ITT (primary analysis) and PP (secondary) mixed effects difference-in difference (DID) model for female *Ae. aegypti* abundance **(Table S9)**, blood-fed *Ae. aegypti* abundance **(Table S10)**, and parous rate **(Table S11)**.

Every covariate other than treatment had statistically significant effects on female *Ae. aegypti* abundance **(Table S9)**. As suggested by the balance in baseline measures, there is no significant difference in baseline mosquito abundance between SR and placebo arms. It is critical to note that in a

DID analysis, the main effect of treatment only estimates a difference to baseline. In contrast, blood-fed *Ae. aegypti* abundance showed no statistically significant difference between SR and placebo arms at baseline, but some effect of seasonality (2-month dummy variables) in both ITT and PP analyses and is consistent with the observation that female *Ae. aegypti* abundance increased significantly between the baseline and post-baseline periods. For parity rate, there was no effect of collection month in the ITT analysis and only two of the months were statistically significant in the PP analysis (**Table S9**).

##### 2.6.2 Secondary endpoints: adult female *Aedes aegypti* abundance, blood-fed *Aedes aegypti* abundance, and *Aedes aegypti* parous rate from ITT and PP mixed effects DID analysis (Cluster-specific Baseline).

Consistent with the 2016 baseline analysis, the number of baseline collections (household surveys) per cluster, and female *Ae. aegypti* evaluated for blood-fed and parity status was lower in the SR arm compared to placebo. Estimates of female *Ae. aegypti* per household collection for the cluster-specific baseline were not balanced between arms, with female abundance higher in the SR than placebo arm (**Table S7**); this difference was not statistically significant (**Table S9).** Estimates for blood-fed *Ae. aegypti* abundance and parity rate were balanced between treatment arms at baseline **(Table S7).**

When comparing cluster-specific baseline and post-baseline periods, the number of household collections and direction of change in entomological indicators were consistent with the 2016 baseline analysis (**Table S7**). Female *Ae. aegypti* abundance decreased overall in SR and placebo arms, but the difference in the SR arm was significantly greater than placebo, with rate reduction outcomes from ITT and PP mixed effects difference-in difference (DID) models of 40.1% and 39.3%, respectively **(Table S8)**. As observed with the 2016 baseline analysis, blood fed *Ae. aegypti* abundance increased between baseline and post-baseline surveys in both treatment arms, but a significant ITT rate reduction of 9.2% was estimated from the effect of SR intervention **(Table S8)**. Parity rate estimates did not significantly change overtime **(Table S8)**.

###### *2.6.2.1 Effects of covariates included in statistical models (Seasonality).*

To control for temporal variation, each month was included as a covariate in the ITT (primary analysis) and PP (secondary) mixed effects difference-in difference (DID) models for female *Ae. aegypti* abundance **(Table S9)**, blood-fed *Ae. aegypti* abundance **(Table S10)**, and parous rate **(Table S11)**. For female *Ae. aegypti* abundance, all covariates in the ITT analysis and 10 of 12 covariates in the PP analyses (other than treatment) had statistically significant effects. Although abundance appeared higher in the SR than placebo arm at baseline, those differences were not statistically significant **(Table S9)**. Blood-fed *Ae. aegypti* abundance showed no statistically significant difference between SR and placebo arms at baseline, although some effect of seasonality was indicated (2 months) in both ITT and PP analyses but these months varied between models and to the 2016 baseline analysis with covariates (**Table S9**). This is consistent with the observation that female *Ae. aegypti* abundance increased significantly between the baseline and post-baseline periods. For parity rate, only two of the month dummy variables were statistically significant in the ITT analysis compared to four in the PP analysis, with every other month covariate not being statistically significant.

##### *2.6.1.2 Interpretation*

The observation that female *Ae. aegypti* abundance was impacted by our SR intervention, supports the significant lower rate of ABV seroconversion observed in the SR treatment arm. This rate reduction was also consistent with self-reports from residents in the study clusters who reported perception of reduced mosquito biting (data to be presented in future publications). The difference in *Ae. aegypti* abundance observed could be due to a combination of repellency or contact irritancy resulting in host-seeking populations staying or exiting to the outdoors. Longer term impacts on the *Ae. aegypti* population could be caused by reduced fitness associated with less blood feeding, or acute chemical lethal effects caused by exposure to transfluthrin at the product point source. The study design to measure the contribution of each of these behavioral effects was outside the scope of our clinical trial, although results from semi-field studies with *Ae. aegypti* populations from our study area support the idea that transfluthrin has behavioral effects beyond mortality.(39) The apparent pyrethroid resistance phenotype in clusters containing SR suggests that exposure to transfluthrin may have had acute toxological impacts, although resistance was also indicated in mosquitoes from placebo clusters (see Section 2.7).

The difference in *Ae. aegypti* blood-fed female abundance between treatment arms was far less pronounced than overall abundance and no changes were estimated in the parous rate. The degree of variation observed in our study is consistent with expectations associated with seasonal differences in ambient temperature and precipitation, which are associated with development of mosquitoes that drives ABV transmission dynamics(40). Our results illustrate the inherent difficulties associated with measuring significant differences in *Ae. aegypti* entomological indicators that has been previously documented . Despite extensive sampling, temporal and spatial heterogeneity was expected among *Ae. aegypti* populations across our study clusters.

#### 2.7 Insecticide Resistance

Exposure of a total 590 *Ae. aegypti* females to 7.5 ug of transfluthrin in standard CDC bottle bioassays from Group A and B, representing mosquito pools originating from SR clusters, resulted in 39.5% (SD = 9.46, CV = 0.73) and 50.9% (SD = 7.7, CV = 0.59) mortality, respectively, at the diagnostic time (30-minutes) (**Figure S9**). Of the 297 mosquitoes evaluated from Group C, originating from clusters receiving placebo intervention, mortality at 30-minutes was 92.9% (Std = ±2.59, CV = 0.11), which was below the threshold for susceptibility and 100% mortality was observed in Group D, also originating from placebo clusters (Std = ±0.29, CV = 0.01; n = 299). One hundred percent mortality was observed for the GDLS population (Std = 0.29, CV = 0.01; n = 299) and USDA Rockefeller reference comparator (Std = 0.60, CV = 0.03; n=297) at the diagnostic time.

Our results indicate the presence of insecticide resistance in *Ae. aegypti* populations in our study area during the trial, although causality due to SR intervention cannot be determined from our study design. The reduced susceptibility in assayed adults from placebo clusters is consistent with the suggestion that other sources of selection pressure could have also contributed to the observed phenotype shift.

#### 2.8 Adverse events (AE) and Serious adverse events (SAE)

In addition to AE reporting directly from participants to our surveillance and entomological staff we analyzed addition sources of data that could measure potential adverse events associated with the SR intervention. Our strategies included: (1) reported reasons for household removing the intervention from their homes during the study; (2) recorded deaths in the active disease surveillance cohort to look for excess deaths in the treated clusters; and (3) reported hospitalization in the active disease surveillance cohort.

A total of 1,700 houses voluntarily requested temporary or permanent removal of the intervention from commencement of the study through last follow-up. Of these 1,700 houses, 855 no longer wanted to participate without giving a reason, 131 were planned to go under construction or be uninhabited, and 23 were based on claims of 'no effect". Only 4 houses specified 'allergy' as a reason for requesting removal, the remaining reasons for requesting removal were largely non-specific. Sixty-two households reported rash or asthma in our perception survey. A more detailed analysis of adverse events and severe adverse events results will be published in subsequent manuscripts.

A total of 203 SAEs were recorded during the trial. All of these were deaths of individuals within the cohort, whether or not the participant's house had an SR intervention or not when the person died. A total of 130 hospitalizations were reported from 18 clusters. There was a report of an infant death associated with a congenital defect in the SR arm and a resident with pityriasis rosea in the placebo arm. The DSMB found no reported SAEs associated with the SR intervention that required halting the trial.

### 3. Context and Significance of the Iquitos cRCT

#### 3.1 Evidence before this study

We searched the National Library of Medicine PubMed database on 17 June 2021 (date of manuscript package submission for initial peer-review) using the search term 'AEDES' in combination with 'SPATIAL REPELLENTS' (81 publications), 'CLINICAL TRIAL' (133 publications), and 'AEDES-BORNE VIRUS VECTOR CONTROL' (40 publications) with no restrictions on date or language. Of the resulting 254 identified publications, the vast majority represent vaccine studies, published protocols of upcoming trials including an Indonesian trial testing the use of *Wolbachia* infected *Ae. aegypt*i to prevent symptomatic dengue, randomized cluster trials evaluating entomological impacts of vector control interventions, and results from laboratory or small-scale field studies evaluating behavior of *Aedes* spp. exposed to vapor-phase pyrethroids. There were 3 reviews describing various alternative strategies to dengue control, but no randomized cluster clinical trials evaluating the effect of spatial repellents on human *Aedes*-borne virus infection. We identified six clinical trials using a rigorous cluster randomized clinical trial (cRCT) design for evaluating vector control interventions and measuring epidemiological endpoints. Four publications evaluated community-participation or mobilization strategies. One, the Camino Verde Study, reported a significant, but modest, reduction in dengue virus human infection. Of the remaining trials, one evaluated permethrin-impregnated school uniforms and another evaluated insecticide treated curtains, showed no human infection or disease protective effect of the intervention. Results from one of the cluster randomized trial (CRT) protocols were published since we conducted our literature search. The CRT with *Wolbachia* infected *Ae. aegypti*, which are less susceptible to dengue virus infection than mosquitoes not infected with *Wolbachia*, reported 77% treatment protective efficacy against symptomatic virologically confirmed human dengue virus infection. Thus, our search revealed a paucity of existing epidemiological evidence for *Aedes* vector control interventions and no existing information on spatial repellent efficacy, or any other chemical intervention, against human *Aedes*-borne virus infection or disease.

##### 3.2 Added value of this study

Our research demonstrates the first conclusive evidence from a cluster-randomized, controlled clinical trial (cRCT) for significant protective efficacy against human *Aedes*-borne virus infection by a chemical-based vector control intervention, which is the most commonly used intervention category among all World Health Organization (WHO) recommendations. Vector interventions are needed for prevention of *Aedes*-borne viral diseases (dengue, Zika, chikungunya, and yellow fever), but their application is hindered by the lack of evidence proving their protective efficacy. Results from our study will help fill this knowledge gap; guide public health authorities responsible for operational management and world-wide prevention of dengue, Zika, yellow fever, and chikungunya disease; and incentivize new strategies for *Aedes*-borne viral disease prevention.

#### 4. Relevance of Outcomes to U.S. Military

The NAMRU-6 was a partner in this research whose primary mission is to protect the U.S. Warfighter. For this reason, we describe the military relevance of our research findings. *Aedes*-borne viral (ABV) diseases such as dengue, chikungunya, Zika, and yellow fever pose a risk to deployed U.S. military service members throughout the tropics where these diseases are endemic. Beyond the direct impact on afflicted individuals, urban dengue epidemics overwhelm public health systems and destabilize societies.(41, 42) Currently, the primary means for ABV disease prevention is controlling the most important mosquito vector*, Aedes aegypt*i. This is achieved through reducing mosquito populations or interfering with mosquito-human contact. Spatial repellents represent a novel vector control intervention designed to reduce human-mosquito contact that may overcome shortcomings of current vector control tools. Our study was conducted in South America, where seroconversion rate to dengue in military personnel has been reported to be 12.4% for those deployed to South America.(43) Although the Department of Defense is actively developing a DENV vaccine that is safe for immunologically naïve individuals,(44) we expect vector control to remain a necessary component of effective integrated dengue, and ABV in general, prevention programs. Successful ABV control programs for the military, therefore, will require innovative strategies that include targeting adult mosquitoes.(45)

#### 5. Supplemental Figures

Supplemental Figure S1. Typical housing structure in the Iquitos, Peru study site (A) and intervention application inside an enrolled household (B).

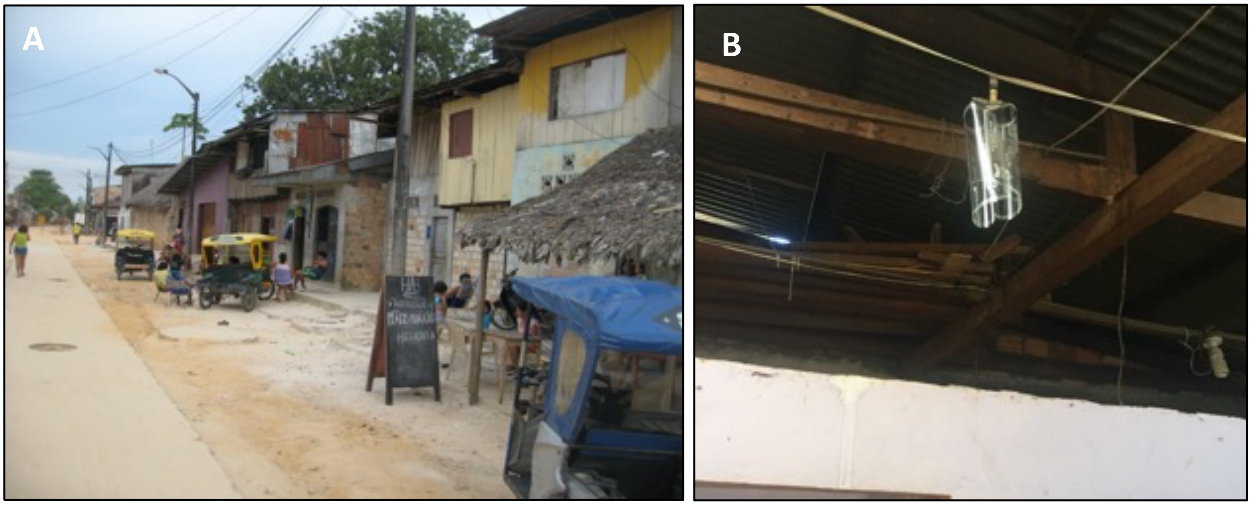

Supplemental Figure S2. Allocation and follow-up of the febrile/disease surveillance cohort for incidence of *Aedes*-borne virus (DENV/ZIKV) laboratory-confirmed cases (secondary endpoint) analysis.

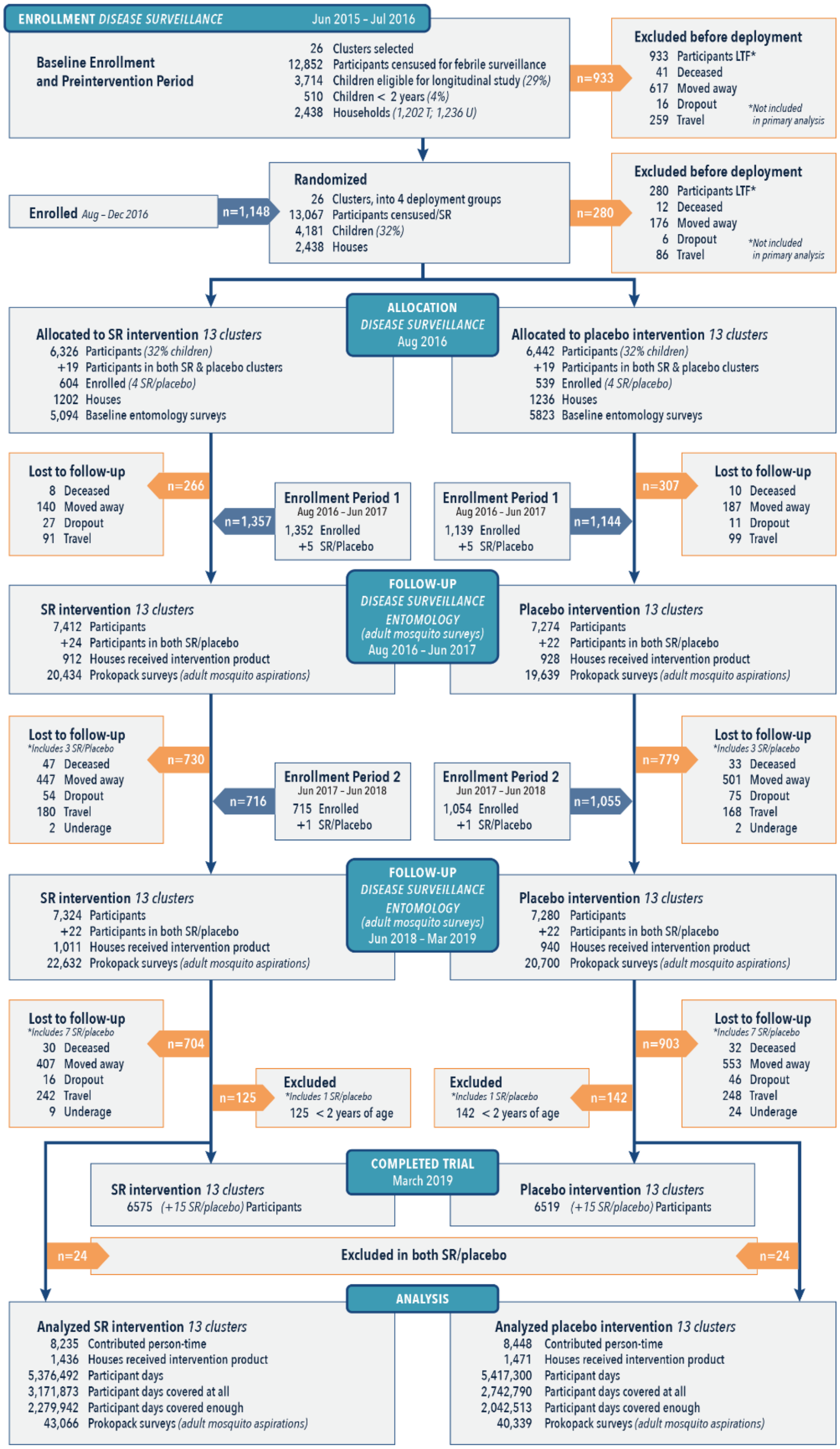

Supplemental Figure S3. Enrollment of households, intervention deployment, replacement, and removal over study period.

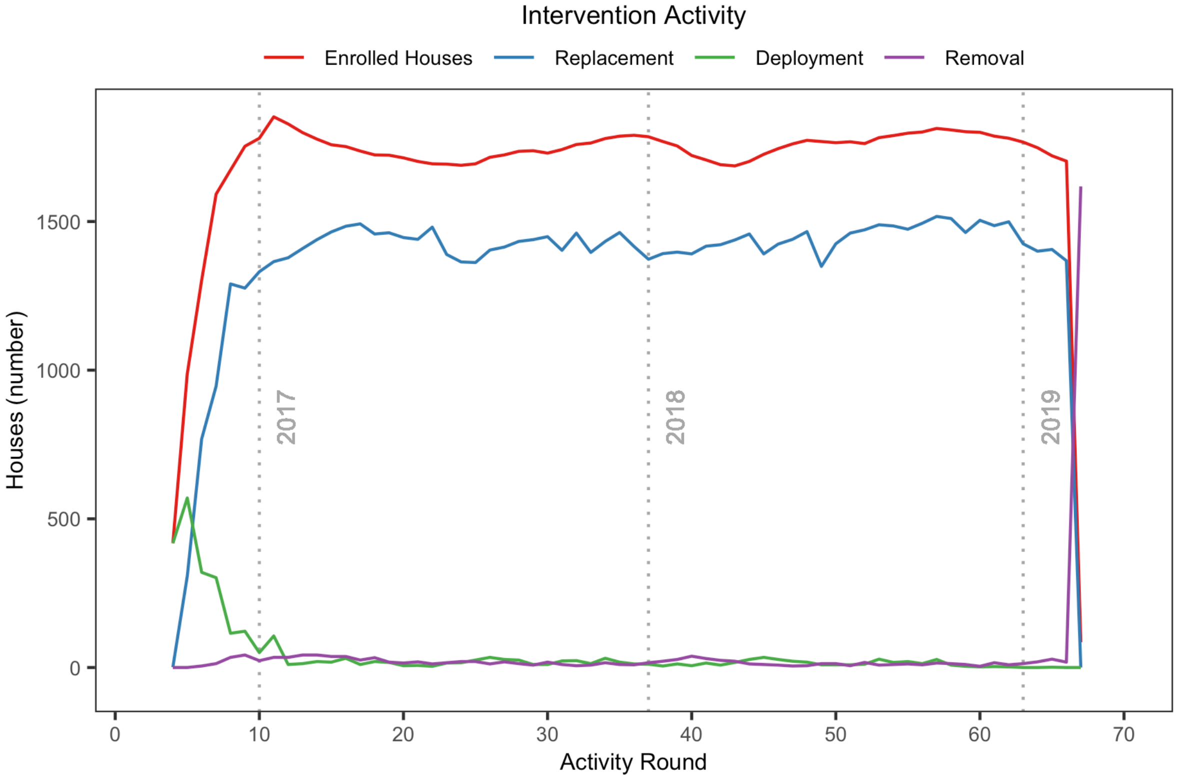

Supplemental Figure S4. Kaplan-Meier curves for placebo and spatial repellent (SR) clusters on arbovirus infection in qualifying participants measured by seroconversion (primary endpoint) by cluster.

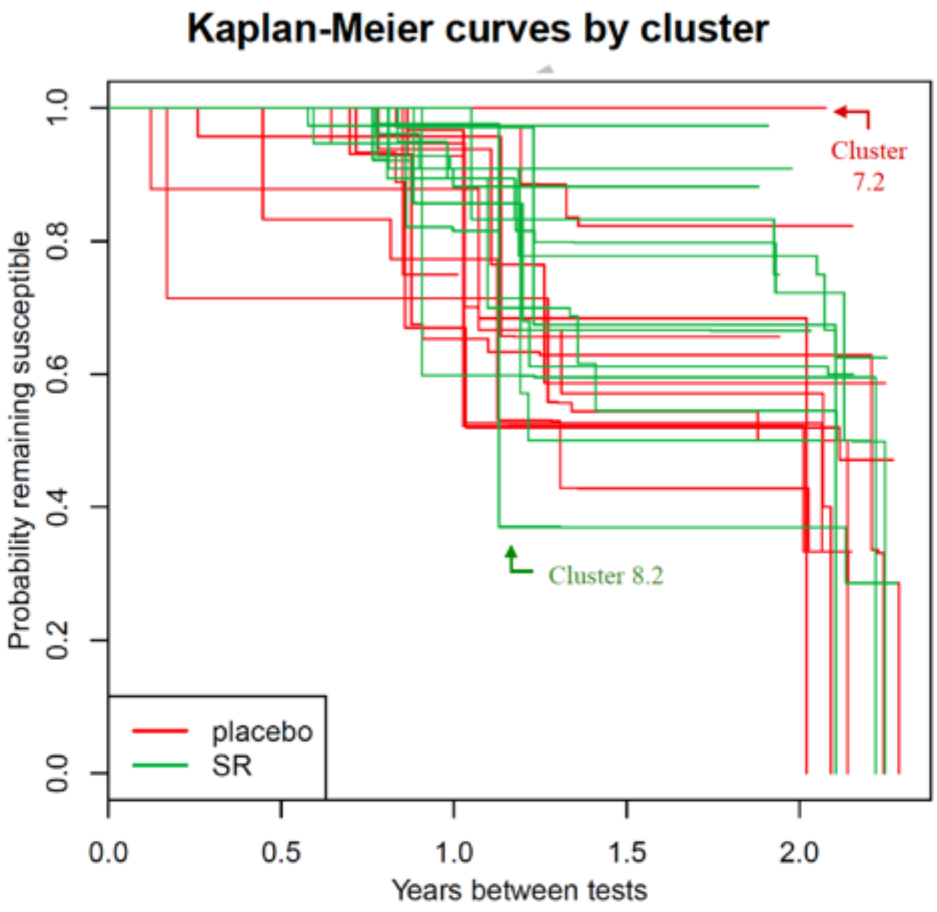

Supplemental Figure S5. Duration between tests (sequential blood samples) of qualifying participants across study clusters for the primary endpoint of seroconversion to *Aedes*-borne virus infection.

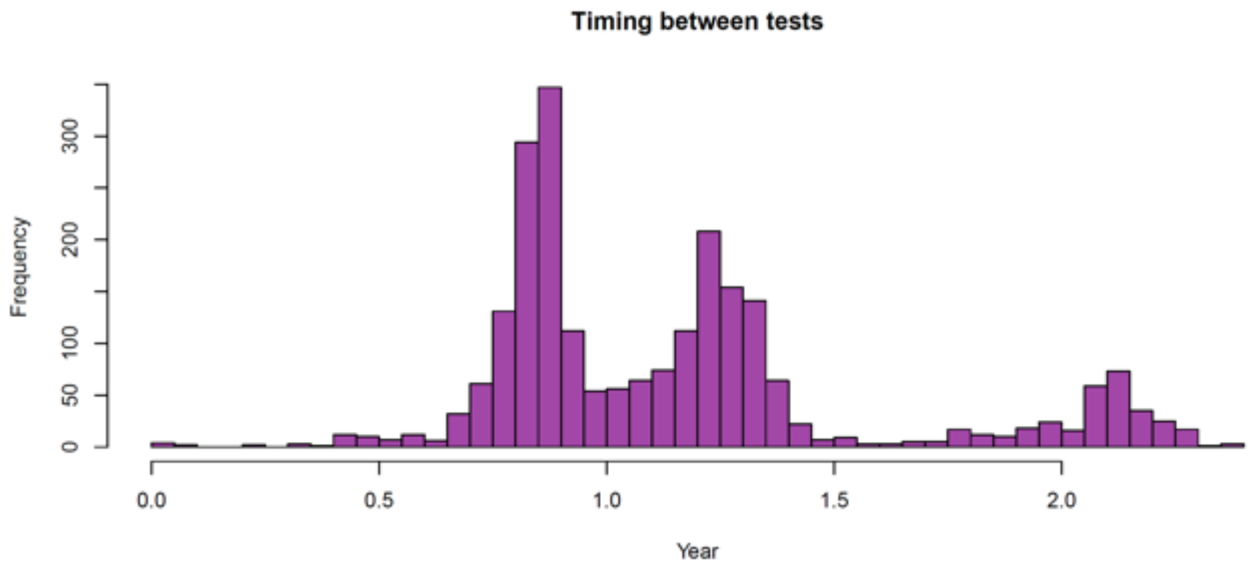

Supplemental Figure S6. Hazard rate reduction summary of PP-2 secondary analysis. Regression plot of hazard risk reduction based on the fraction of time a qualifying participant house had at least as many SR products as recommended between sequential blood sampling. This analysis reconsidered the threshold for inclusion in per protocol analysis and assessed the hazard risk reduction (fixed effect) as a function of inclusion thresholds (fraction covered), which ranged from 5% to 100%. As the coverage inclusion criteria became more strict (100%), fewer individuals were included in the analysis, resulting in fewer total seroconversions (primary endpoint) among all qualifying participants in SR and placebo clusters. Mean results from the primary analysis are included as a grey dashed horizontal line for reference.

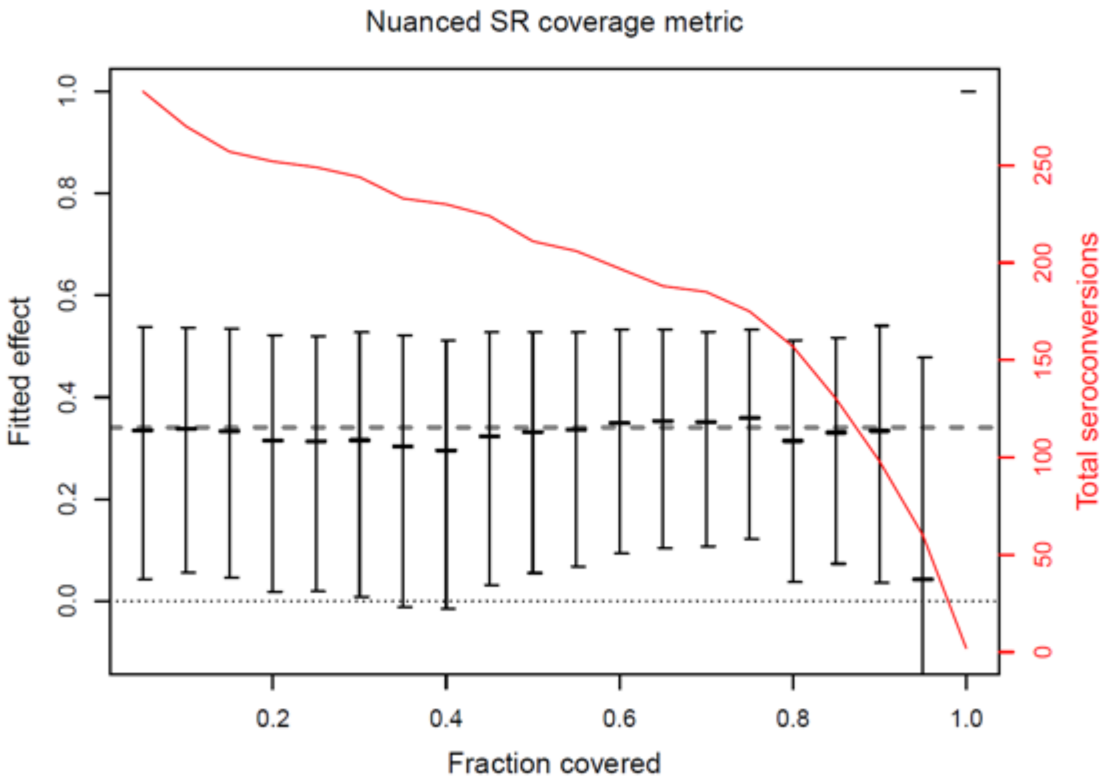

Supplemental Figure S7. Hazard rate reduction summary of PP-4 secondary analysis. Regression plot of hazard risk reduction based on the fraction of time a qualifying participant house had any SR products between sequential blood sampling. This analysis reconsidered the threshold for inclusion in per protocol analysis and assessed the hazard risk reduction (fixed effect) as a function of inclusion thresholds (fraction covered), which ranged from 5% to 100%. As the coverage inclusion criteria became more strict (100%), fewer individuals were included in the analysis, resulting in fewer total seroconversions (primary endpoint) between all qualifying participants in SR and placebo clusters. Mean results from the primary analysis are included as a grey dashed horizontal line for reference.

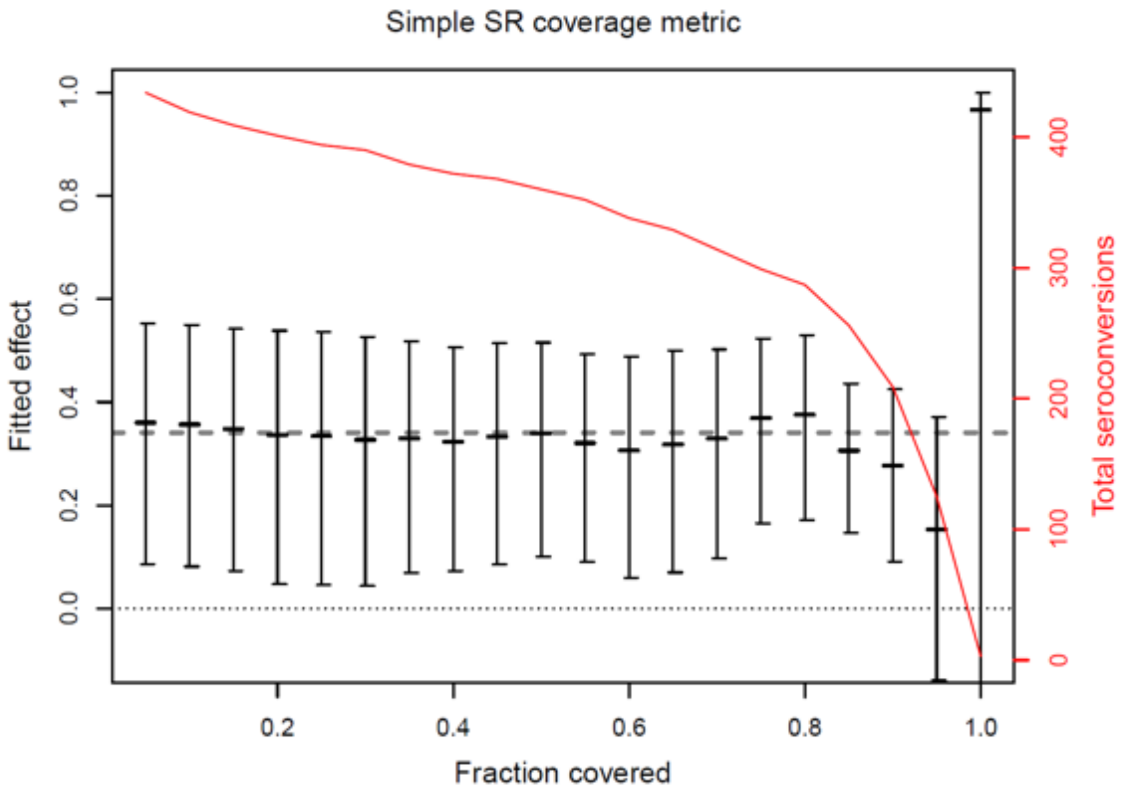

Supplemental Figure S8. Mean densities of adult female *Aedes aegypti* collected per household survey in 13 spatial repellent (SR) and 13 placebo clusters by study month during 2015 to July 2016 preintervention (cluster-based baseline) (panel A) and January-September 2019 post intervention surveys (panel B). Pre- and post-intervention full-pupal demographic surveys that included collection of adult mosquitos with Prokopack aspirator were carried out over 2–4-month intervals. Shaded areas represent the 95% Confidence Interval around the mean.

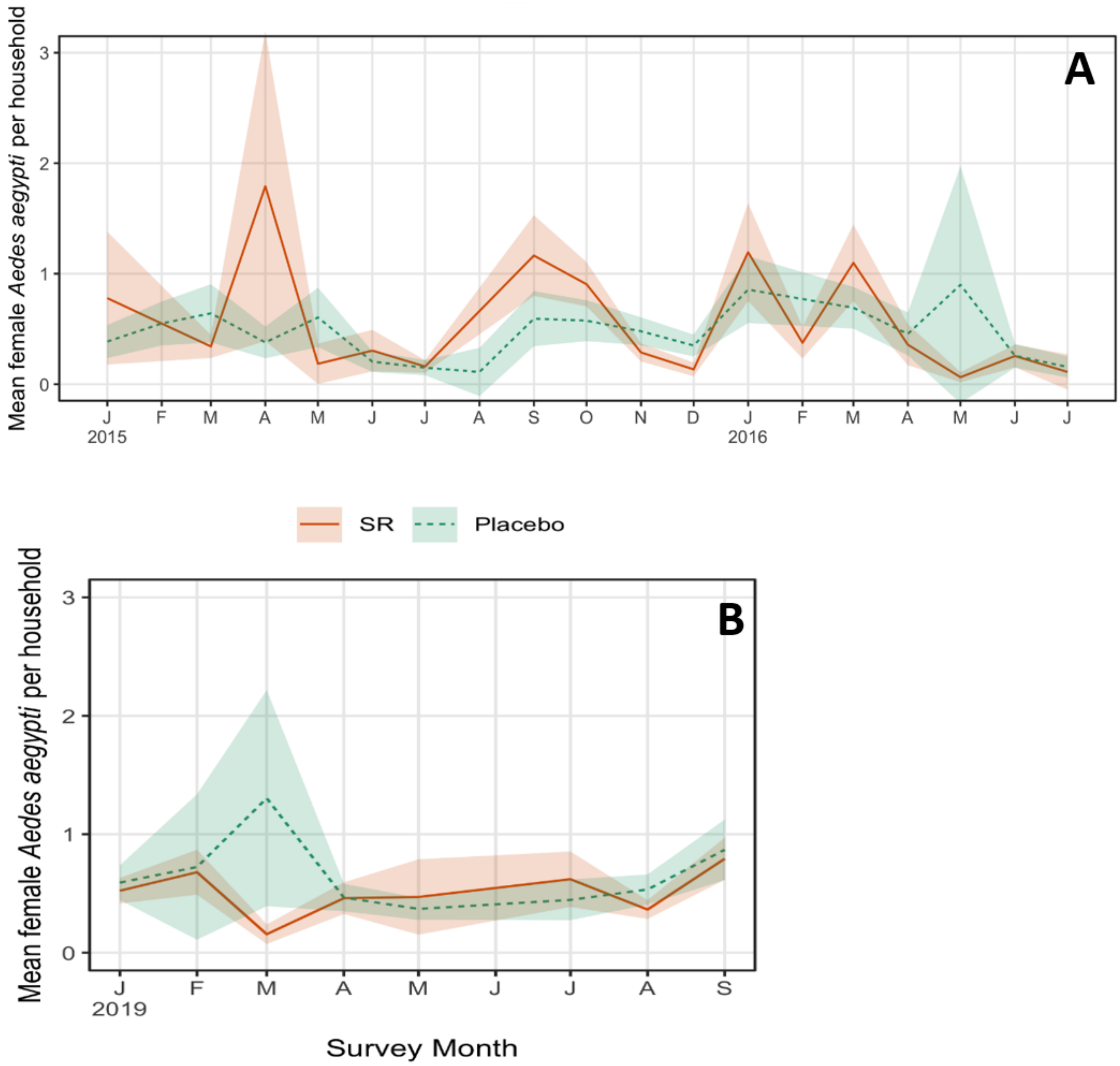

Supplemental Figure S9. Insecticide resistance status of female *Aedes aegypti*. Summary mortality rates from six populations originating in the study area to transfluthrin (7.5 ug) using the standard Centers for Disease Control (CDC) bottle bioassay. Note that Group D and the GDLS lines are obscured by the Rockefeller strain – all three strains are completely susceptible to transfluthrin.

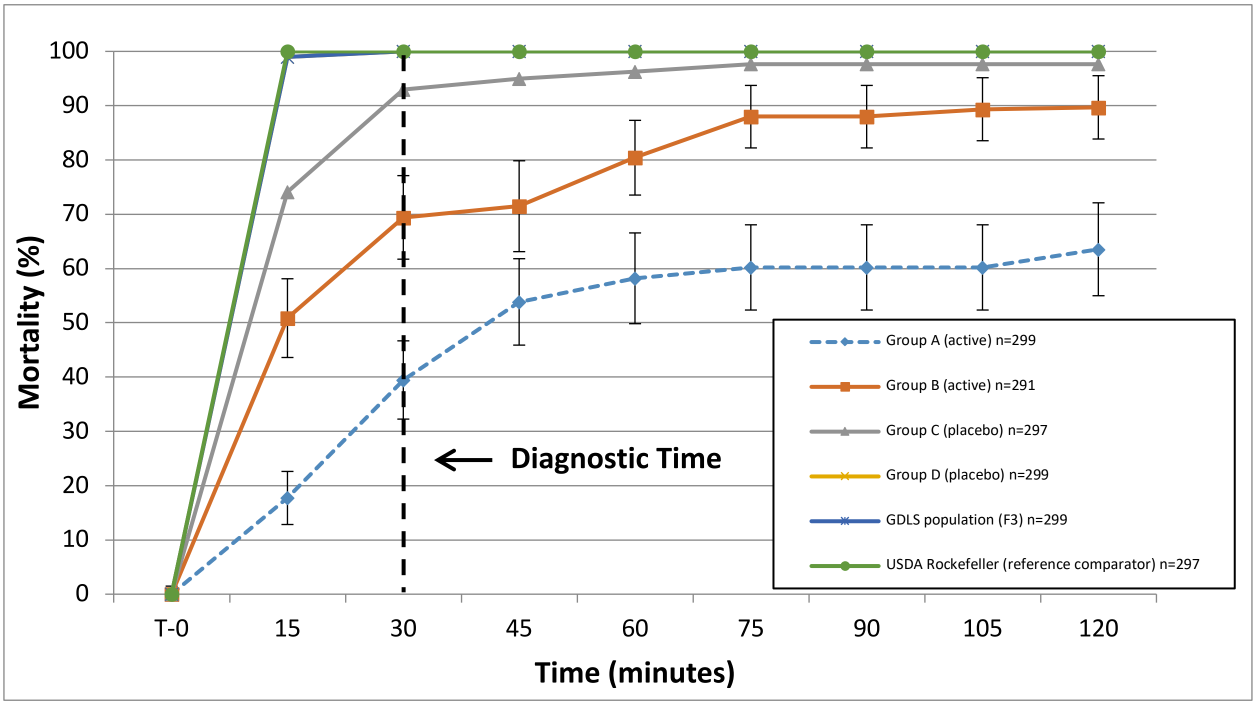

### 6. Supplemental Tables

#### Supplemental Table S1. Summary overview of trial endpoints, analysis, and location of reporting in SI document.

| | Description | Endpoint | Analysis Category | ITT /PP | Methods | Results | Table/ Figure | Analysis Description |
|---|---|---|---|---|---|---|---|---|
| *Epidemiological Indicators* | *Aedes*-borne virus (ABV) sero-conversion | Primary | Baseline | | 1.5.2 | 2.5.1 | Table1 | Baseline longitudinal cohort characteristics |
| | | | Primary | ITT | 1.5.2.1 | 2.5.1.1 | Table 2, S3, Figure S4 | Survival analysis with proportional hazards model, exponential distribution assumption for baseline hazard |
| | | | Secondary | ITT | 1.5.2.2 | 2.5.1.2 | Table S3 | Mixed effects logistic regression model (MELR) (original model SAP) |
| | | | | ITT | 1.5.2.3 | 2.5.1.3 | Table S3 | MELR model using only subjects with samples separated by 9-15 months |
| | | | Secondary | PP-1 | 1.5.2.4.1 | 2.5.1.4.1 | Table S4 | SA using subjects with 100% indoor spatial coverage (enough SR products) for $\geq$75% of days they participated |
| | | | | PP-2 | 1.5.2.4.2 | 2.5.1.4.2 | Figure S6 | Analysis in PP-1 varying coverage rate from 0 to 100% to assess robustness of 75% coverage |
| | | | | PP-3 | 1.5.2.4.3 | 2.5.1.4.3 | Table S4 | SA using subjects with some SR products (access to some bedrooms resulted in < 100% spatial coverage) for $\geq$75% of days they participated |
| | | | | PP-4 | 1.5.2.4.4 | 2.5.1.4.4 | Figure S7 | Analysis in PP-3 varying coverage rate from 0 to 100% to assess robustness of 75% coverage |
| | DENV/ZIKV cases | Secondary | Baseline | | 1.5.3 | 2.5.2 | Table S5 | Baseline disease surveillance cohort |
| | | | Primary | ITT | 1.5.3 | 2.5.2 | Table S5 | Fixed effect Poisson Generalized linear model |
| *Entomological Indicators* | Baseline/post-intervention | | | | 1.5.4 | 2.6.1 | Table S6 | Baseline characteristics using '2016 Baseline' approach |
| | | | | | 1.5.4 | 2.6.2 | Table S7 | Baseline characteristics using 'cluster specific Baseline' approach |
| | Indoor adult female *Aedes aegypti* abundance | Secondary | Primary | ITT | 1.5.4.1 | 2.6.1 | Table S8, S9 | Mixed effects difference-in-difference Poison regression (MEDIDPR), with factor-level monthly covariates to account for seasonality (2016 baseline) |
| | | | Secondary | PP | 1.5.4.2 | 2.6.1 | Table S8, S9 | MEDIDPR, with month covariate, using houses with 100% indoor spatial coverage (enough SR products) for $\geq$ 75% of days they participated (2016 baseline) |
| | | | | ITT | 1.5.4.3 | 2.6.2 | Table S8, S9 | MEDIDPR, with factor-level monthly covariates to account for seasonality (Cluster-based baseline) |
| | | | | PP | 1.5.4.4 | 2.6.2 | Table S8, S9 | MEDIDPR, with month covariate, using houses with 100% indoor spatial coverage (enough SR products) for $\geq$ 75% of days they participated (Cluster-based baseline) |
| | Blood-fed female *Aedes aegypti* abundance | Secondary | Primary | ITT | 1.5.5.1 | 2.6.1 | Table S8, S10 | MEDIDPR, with factor-level monthly covariates to account for seasonality (2016 baseline) |
| | | | Secondary | PP | 1.5.5.2 | 2.6.1 | Table S8, S10 | MEDIDPR, with month covariate, using houses with 100% indoor spatial coverage (enough SR products) for $\geq$ 75% of days they participated (2016 baseline) |
| | | | | ITT | 1.5.5.3 | 2.6.2 | Table S8, S10 | MEDIDPR, with factor-level monthly covariates to account for seasonality (Cluster-based baseline) |
| | | | | PP | 1.5.5.4 | 2.6.2 | Table S8, S10 | MEDIDPR, with month covariate, using houses with 100% indoor spatial coverage (enough SR products) for $\geq$ 75% of days they participated (Cluster-based baseline) |
| | *Aedes aegypti* parity rate | Secondary | Primary | ITT | 1.5.6.1 | 2.6.1 | Table S8, S11 | MEDIDPR, with factor-level monthly covariates to account for seasonality (2016 baseline) |
| | | | Secondary | PP | 1.5.6.2 | 2.6.1 | Table S8, S11 | MEDIDPR, with month covariate, using houses with 100% indoor spatial coverage (enough SR products) for $\geq$ 75% of days they participated (2016 baseline) |
| | | | | ITT | 1.5.6.3 | 2.6.2 | Table S8, S11 | MEDIDPR, with factor-level monthly covariates to account for seasonality (2015-2016 surveys with no SR product) |
| | | | | PP | 1.5.6.4 | 2.6.2 | Table S8, S11 | MEDIDPR, with month covariate, using houses with 100% indoor spatial coverage (enough SR products) for $\geq$ 75% of days they participated (Cluster-based baseline) |

Supplemental Table S2. Summary of intervention coverage metrics for spatial repellent (SR) and placebo treatment arms.

| **Coverage Metric** | **SR (n=13)** | **Placebo (n=13)** |
|---|---|---|
| Participating houses (%±sd)<br>(min, max) | 47.0 ± 10.4<br>(30, 68.3) | 41.4 +/- 8.3<br>(23.1, 55.8) |
| Location days participant covered at any house intervention application rate (%±sd)<br>(min, max) | 82.9 ± 3.7<br>(78.1, 90.3) | 80.3 ± 3.8<br>(70.3, 85.0) |
| Location days participant covered at manufactures' recommended house application rate (%±sd)<br>(min, max) | 72.7 ± 11.4<br>(52.5, 88.4) | 74.5 ± 6.3<br>(63.2, 86.3) |

Supplemental Table S3. Protective efficacy (PE) estimates from intent to treat (ITT) analyses for the spatial repellent intervention against *Aedes*-borne virus infection in qualifying participants measured by seroconversion (primary endpoint), including covariate effects.

| ITT analysis | Hazard rate ratio (95% CI) | PE (%) (95% CI) | One-sided p-value | Covariate | Odds ratio (95% CI) | Two-sided p-value |
|---|---|---|---|---|---|---|
| Survival analysis | 0.659 (−∞,0.931) | 34.1 (6.9,∞) | 0.024 | | | |
| | | | | Age | 1.046 (1.029-1.063) | $6.8 \times 10^{-6}$ |
| | | | | Male | 0.956 (0.821-1.112) | 0.62 |
| **ITT analysis** | **Odds ratio (95% CI)** | **PE (%) (95% CI)** | **One-sided p-value** | **Covariate** | **Odds ratio (95% CI)** | **Two-sided p-value** |
| Odds reduction | 0.547 (−∞,0.883) | 45.3 (11.7,∞) | 0.019 | | | |
| | | | | Age | 1.071 (1.042-1.100) | $6.9 \times 10^{-7}$ |
| | | | | Male | 0.950 (0.767-1.177) | 0.64 |
| Odds reduction on including 9–15-month sampling intervals | 0.577 (−∞,0.879) | 42.3 (12.1,∞) | 0.016 | | | |
| | | | | Age | 1.065 (1.028-1.103) | 0.0005 |
| | | | | Male | 0.970 (0.767-1.299) | 0.779 |

Supplemental Table S4. Protective efficacy (PE) estimates from per protocol (PP) analyses for the spatial repellent intervention against *Aedes*-borne virus infection in qualifying participants measured by seroconversion (primary endpoint), including covariate effects.

| Inclusion criteria (designated PP label) | Hazard rate ratio (95% CI) | PE (%) (95% CI) | One-sided p-value | Covariate | Odds ratio (95% CI) | Two-sided p-value |
|---|---|---|---|---|---|---|
| ≥75% of days meeting manufacturer's specifications for house intervention application rate between sequential blood sampling (PP-1) | 0.626 (−∞,0.806) | 37.4 (19.4,∞) | 0.001 | | | |
| | | | | Age | 1.039 (1.004-1.076) | 0.029 |
| | | | | Male | 0.877 (0.650-1.181) | 0.387 |
| ≥75% of days with any amount of intervention in house between sequential blood sampling (PP-3) | 0.625 (−∞,0.831) | 37.4 (19.4,∞) | 0.007 | | | |
| | | | | Age | 1.038 (1.012-1.064) | 0.032 |
| | | | | Male | 0.956 (0.759-1.120) | 0.702 |

Supplemental Table S5. Summary of baseline characteristics for spatial repellent (SR) and placebo treatment arms with incidence of *Aedes*-borne virus (DENV/ZIKV) laboratory-confirmed cases (secondary endpoint) and protective efficacy (PE) estimate from intent to treat (ITT) analyses using Poisson generalized linear regression. No correction for multiple testing was performed for secondary endpoint analyses and, as such, in accordance with CONSORT guidelines, we do not present p-values.

| **Treatment arm (number of clusters)** | **Participant-days per cluster (mean+sd) (min,max)** | **No. Laboratory-confirmed DENV/ ZIKV disease** | **DENV/ZIKV disease incidence per 100,000 p-days (mean+sd)** |
|---|---|---|---|
| SR (n=13) | 413,601 ± 136,789<br>(214,595, 624,045) | 51 | 8.64 ± 6.38 |
| Placebo (n=13) | 417,392 ± 125,860<br>(124,024, 592,326) | 45 | 7.61 ± 8.17 |
| | **Rate ratio (95% one-sided CI)** | **Rate reduction (%) (95% one-sided CI)** | |
| SR vs placebo comparison | 1.142<br>(−∞,1.599) | -14.2<br>(-59.9,∞) | |

Supplemental Table S6. Summary of baseline and post-baseline characteristics for entomological indicators (secondary endpoints) for intent to treat mixed effects difference-in-difference (DID) analysis using '2016 baseline'[1].

| Indicator | Characteristic | No. per Cluster (mean+sd) (min,max) | |
|---|---|---|---|
| | | **SR (n=13)** | **Placebo (n=13)** |
| Indoor adult female *Aedes aegypti* abundance | Baseline collections conducted | 428.7 ± 277.8 (48,926) | 483.2 ± 198.9 (73,785) |
| | Baseline indoor adult female *Ae. aegypti* captured per collection | 0.277 ± 0.153 (0.086,0.603) | 0.279 ± 0.122 (0.095,0.539) |
| | Number captured during post-baseline collections | 47,518 | 43,417 |
| | Post-baseline female *Ae. aegypti* captured per collection | 0.276 ± 0.091 (0.155,0.475) | 0.391 ± 0.142 (0.182,0.665) |
| Blood-fed *Aedes aegypti* abundance | Baseline collections evaluated for blood-fed status | 54.4 ± 37.4 (6,103) | 65.2 ± 35.7 (13,134) |
| | Baseline blood-fed *Ae. aegypti* per collection | 0.554 ± 0.130 (0.231,0.783) | 0.529 ± 0.083 (0.327,0.615) |
| | No. of post-baseline *Ae. aegypti* assessed for blood-fed status | 9,257 | 11,496 |
| | Post-baseline blood-fed *Ae. aegypti* per collection | 0.620 ± 0.038 (0.560,0.672) | 0.649 ± 0.032 (0.591,0.7022) |
| *Aedes aegypti* parity rate | Baseline *Ae. aegypti* evaluated for parity per collection | 21.9 ± 16.6 (1,46) | 23.2 ± 15.7 (2,56) |
| | Baseline *Ae. aegypti* parity rate per cluster | 0.754 ± 0.274 (0,1) | 0.749 ± 0.162 (0.429,1) |
| | No. of post-baseline *Ae. aegypti* evaluated for parity | 3,661 | 3,096 |
| | Post-baseline parity rate for female *Ae. aegypti* per cluster | 0.773 ± 0.016 (0.738, 0.812) | 0.787 ± 0.043 (0.707, 0.861) |

[1] Intent to treat (ITT) (primary analysis) mixed effects difference-in-difference (DID) Poisson regression specifying measurements made throughout 2016 as 'baseline', with post-2016 measurements as 'post-baseline'.

Supplemental Table S7. Summary of baseline and post-baseline characteristics for entomological indicators (secondary endpoints) for intent to treat mixed effects difference-in-difference (DID) analysis using 'cluster-specific baseline'[1].

| Indicator | Variable | No. per Cluster (mean±sd) (min,max) | |
|---|---|---|---|
| | | SR (n=13) | Placebo (n=13) |
| Indoor adult female *Aedes aegypti* abundance | Baseline collections conducted | 286.1 ± 121.9 (122,534) | 321.9 ± 90.5 (203,494) |
| | Baseline indoor adult female *Ae. aegypti* captured per collection | 0.713 ± 0.538 (0.178, 1.680) | 0.468 ± 0.162 (0.164, 0.766) |
| | Number captured during post-baseline collections | 46,699 | 53,091 |
| | Post-baseline female *Ae. aegypti* captured per collection | 0.278 ± 0.087 (0.155, 0.464) | 0.379 ± 0.139 (0.185, 0.646) |
| Blood-fed *Aedes aegypti* abundance | Baseline collections evaluated for blood-fed status | 59.6 ± 20.8 (35,109) | 65.7 ± 27.8 (31,126) |
| | Baseline blood-fed *Ae. aegypti* per collection | 0.545 ± 0.063 (0.46,0.634) | 0.498 ± 0.09 (0.366,0.632) |
| | No. of post-baseline *Ae. aegypti* assessed for blood-fed status | 9,727 | 12,927 |
| | Post-baseline blood-fed *Ae. aegypti* per collection | 0.62 ± 0.037 (0.573,0.676) | 0.637 ± 0.031 (0.589,0.696) |
| *Aedes aegypti* parity rate | Baseline *Ae. aegypti* evaluated for parity per collection | 22.1 ± 10.9 (10,43) | 23.5 ± 10.3 (9,49) |
| | Baseline *Ae. aegypti* parity rate per cluster | 0.764 ± 0.133 (0.55,1) | 0.720 ± 0.164 (0.326,0.9) |
| | No. of post-baseline *Ae. aegypti* evaluated for parity | 3,239 | 4,075 |
| | Post-baseline parity rate for female *Ae. aegypti* per cluster | 0.789 ± 0.047 (0.698, 0.861) | 0.771 ± 0.02 (0.741, 0.805) |

[1] Intent to treat (ITT) (primary analysis) and DID Poisson regression specifying measurements made in 2016 up to the first date of intervention deployed in each cluster as 'baseline', with all measurements following that date as 'post-baseline' for that cluster, even for houses that did not enroll until 2017 or later.

Supplemental Table S8. Rate reduction summary of indoor adult female *Aedes aegypti* abundance, blood-fed female abundance, and parity rates (secondary endpoints) for primary (ITT) and secondary analyses (PP). No correction for multiple testing was performed for secondary endpoint analyses and, as such, in accordance with CONSORT guidelines, we do not present p-values.

| **Indicator** | **Statistics** | **2016 Baseline[1]** | | **Cluster-specific Baseline[2]** | |
|---|---|---|---|---|---|
| | | **ITT** | **PP** | **ITT** | **PP** |
| Indoor adult female *Aedes aegypti* abundance | Rate ratio (95% one-sided CI) | 0.714 (−∞,0.759) | 0.737 (−∞,0.784) | 0.599 (−∞,0.633) | 0.607 (−∞,0.641) |
| | Rate reduction (%) (95% one-sided CI) | 28.6 (24.1,∞) | 26.3 (16.2,∞) | 40.1 (36.7,∞) | 39.3 (35.9,∞) |
| Blood-fed female *Aedes aegypti* abundance | Rate ratio (95% one-sided CI) | 0.876 (−∞,0.958) | 0.876 (−∞,0.958) | 0.908 (−∞,0.985) | 0.929 (−∞,1.06) |
| | Rate reduction (%) (95% one-sided CI) | 12.4 (4.2,∞) | 12.4 (4.2,∞) | 9.20 (1.49,∞) | 7.10 (-6,∞) |
| *Aedes aegypti* parity rate | Rate ratio (95% one-sided CI) | 0.922 (−∞,1.057) | 0.921 (−∞,1.056) | 0.928 (−∞,1.058) | 0.909 (−∞,0.986) |
| | Rate reduction (%) (95% one-sided CI) | 7.75 (-5.70,∞) | 7.87 (-5.60,∞) | 7.24 (-5.80,∞) | 9.10 (-1.2,∞) |

[1] Intent to treat (ITT) (primary analysis) and per protocol (PP) (secondary analysis) mixed effects difference-in-difference (DID) Poisson regression specifying measurements made throughout 2016 as 'baseline', with post-2016 measurements as 'post-baseline'; PP analysis only considering houses with SR application rates meeting manufactures specifications ≥75% of the days between sequential blood sampling (designated PP-1).

[2] ITT and PP mixed effects DID Poisson regression specifying measurements made in 2016 up to the first date of intervention deployed in each cluster as 'baseline', with all measurements following that date as 'post-baseline' for that cluster, even for houses that did not enroll until 2017 or later; PP analysis only considering houses with SR application rates meeting manufactures specifications ≥75% of the days between sequential blood sampling (designated PP-1).

Supplemental Table S9. Covariate effects for indoor adult female *Aedes aegypti* abundance mixed effect difference-in-difference (DID) Poisson regression models for intent to treat (ITT) and per-protocol (PP) analyses. No correction for multiple testing was performed for secondary endpoint analyses and, as such, in accordance with CONSORT guidelines, we do not present p-values.

| **Covariate** | **2016 Baseline[1]** | | **Cluster-specific Baseline[2]** | |
|---|---|---|---|---|
| | **ITT** | **PP** | **ITT** | **PP** |
| | **Hazard rate ratio (95% CI)** | **Hazard rate ratio (95% CI)** | **Hazard rate ratio (95% CI)** | **Hazard rate ratio (95% CI)** |
| Treatment | 1.005 (0.781-1.294) | 0.989 (0.771-1.271) | 1.246 (0.973-1.595) | 1.247 (0.97 - 1.604) |
| Post-baseline | 1.800 (1.711-1.893) | 1.812 (1.723-1.906) | 0.790 (0.754-0.828) | 0.793 (0.756 - 0.831) |
| February | 0.714 (0.664-0.768) | 0.711 (0.673-0.752) | 0.682 (0.648-0.717) | 0.692 (0.656 - 0.729) |
| March | 0.698 (0.661-0.736) | 0.754 (0.715-0.796) | 0.721 (0.689-0.755) | 0.756 (0.721 - 0.793) |
| April | 0.713 (0.678-0.751) | 0.749 (0.708-0.791) | 0.787 (0.751-0.825) | 0.82 (0.781 - 0.861) |
| May | 0.714 (0.677-0.753) | 0.437 (0.409-0.466) | 0.409 (0.385-0.435) | 0.416 (0.391 - 0.442) |
| June | 0.428 (0.402-0.455) | 0.479 (0.449-0.510) | 0.437 (0.412-0.463) | 0.456 (0.429 - 0.483) |
| July | 0.455 (0.428-0.484) | 0.529 (0.498-0.562) | 0.471 (0.445-0.499) | 0.491 (0.463 - 0.52) |
| August | 0.503 (0.474-0.534) | 0.659 (0.624-0.696) | 0.607 (0.577-0.639) | 0.633 (0.6 - 0.667) |
| September | 0.628 (0.596-0.662) | 1.201 (1.147-1.257) | 0.996 (0.955-1.038) | 1.034 (0.99 - 1.079) |
| October | 1.143 (1.093-1.195) | 1.252 (1.198-1.308) | 1.058 (1.017-1.100) | 1.088 (1.045 - 1.133) |
| November | 1.200 (1.500-1.252) | 1.171 (1.112-1.225) | 0.935 (0.897-0.974) | 0.962 (0.923 - 1.004) |
| December | 1.120 (1.072-1.170) | 0.856 (0.815-0.899) | 0.693 (0.663-0.724) | 0.712 (0.68 - 0.745) |

[1] Intent to treat (ITT) (primary analysis) and per protocol (PP) (secondary analysis) mixed effects difference-in-difference (DID) Poisson regression specifying measurements made throughout 2016 as 'baseline', with post-2016 measurements as 'post-baseline'; PP analysis only considering houses with SR application rates meeting manufactures specifications $\geq$75% of the days between sequential blood sampling (designated PP-1).

[2] ITT and PP mixed effects DID Poisson regression specifying measurements made in 2016 up to the first date of intervention deployed in each cluster as 'baseline', with all measurements following that date as 'post-baseline' for that cluster, even for houses that did not enroll until 2017 or later; PP analysis only considering houses with SR application rates meeting manufactures specifications $\geq$75% of the days between sequential blood sampling (designated PP-1).

Supplemental Table S10. Covariate effects for blood-fed female *Aedes aegypti* abundance mixed effect difference-in-difference (DID) Poisson regression models for intent to treat (ITT) and per-protocol (PP) analyses. No correction for multiple testing was performed for secondary endpoint analyses and, as such, in accordance with CONSORT guidelines, we do not present p-values.

| Covariate | 2016 Baseline[1] | | Cluster-based baseline[2] | |
|---|---|---|---|---|
| | ITT | PP | ITT | PP |
| | Hazard rate ratio (95% CI) | Hazard rate ratio (95% CI) | Hazard rate ratio (95% CI) | Hazard rate ratio (95% CI) |
| Treatment | 1.101 (0.995-1.219) | 1.101 (0.995 – 1.219) | 1.076 (0.982-1.179) | 1.097 (0.946 - 1.273) |
| Post-baseline | 1.224 (1.134-1.322) | 1.226 (1.135 – 1.324) | 1.250 (1.163-1.342) | 1.148 (1.021 - 1.29) |
| February | 0.837 (0.768-0.975) | 0.842 (0.772 – 0.918) | 0.865 (0.824-0.934) | 1.03 (0.907 - 1.17) |
| March | 0.952 (0.877-1.032) | 0.942 (0.867 – 1.025) | 0.898 (0.798-0.965) | 0.918 (0.816 - 1.032) |
| April | 0.911 (0.838-0.990) | 0.912 (0.838 – 0.993) | 0.944 (0.835-1.016) | 1.072 (0.953 - 1.207) |
| May | 0.988 (0.905-1.079) | 0.990 (0.905 – 1.083) | 0.985 (0.877-1.072) | 0.915 (0.797 - 1.05) |
| June | 0.933 (0.852-1.022) | 0.936 (0.853 – 1.026) | 0.942 (0.905-1.027) | 0.841 (0.727 - 0.972) |
| July | 1.005 (0.923-1.096) | 1.007 (0.923 – 1.100) | 1.004 (0.864-1.091) | 0.914 (0.798 - 1.046) |
| August | 1.009 (0.936-1.089) | 1.008 (0.933 – 1.089) | 1.006 (0.924-1.083) | 0.953 (0.841 - 1.081) |
| September | 1.010 (0.947-1.078) | 1.013 (0.948 – 1.082) | 0.971 (0.934-1.031) | 0.974 (0.881 - 1.076) |
| October | 0.964 (0.904-1.028) | 0.967 (0.906 – 1.033) | 0.935 (0.914-0.992) | 0.971 (0.88 - 1.07) |
| November | 1.001 (0.940-1.067) | 1.011 (0.938 – 1.068) | 0.925 (0.925-1.041) | 0.939 (0.849 - 1.038) |
| December | 0.953 (0.890-1.020) | 0.956 (0.891 – 1.025) | 0.932 (0.874-0.994) | 0.899 (0.81 - 0.997) |

[1] Intent to treat (ITT) (primary analysis) and per protocol (PP) (secondary analysis) mixed effects difference-in-difference (DID) Poisson regression specifying measurements made throughout 2016 as 'baseline', with post-2016 measurements as 'post-baseline'; PP analysis only considering houses with SR application rates meeting manufactures specifications ≥75% of the days between sequential blood sampling (designated PP-1).

[2] ITT and PP mixed effects DID Poisson regression specifying measurements made in 2016 up to the first date of intervention deployed in each cluster as 'baseline', with all measurements following that date as 'post-baseline' for that cluster, even for houses that did not enroll until 2017 or later; PP analysis only considering houses with SR application rates meeting manufactures specifications ≥75% of the days between sequential blood sampling (designated PP-1).

Supplemental Table S11. Covariate effects for *Aedes aegypti* parity rate mixed effect difference-in-difference (DID) Poisson regression models for intent to treat (ITT) and per-protocol (PP) analyses. No correction for multiple testing was performed for secondary endpoint analyses and, as such, in accordance with CONSORT guidelines, we do not present p-values.

| Covariate | 2016 Baseline[1] | | Cluster-specific baseline[2] | |
|---|---|---|---|---|
| | ITT | PP | ITT | PP |
| | Hazard rate ratio (95% CI) | Hazard rate ratio (95% CI) | Hazard rate ratio (95% CI) | Hazard rate ratio (95% CI) |
| Treatment | 1.095 (0.940-1.276) | 1.096 (0.940-1.276) | 1.099 (0.948-1.275) | 1.074 (0.98 - 1.178) |
| Post-baseline | 1.020 (0.904-1.152) | 1.019 (0.903-1.151) | 1.149 (1.022-1.292) | 1.247 (1.161 - 1.34) |
| February | 1.012 (0.890-1.151) | 1.010 (0.883-1.155) | 1.031 (0.912-1.165) | 0.87 (0.802 - 0.943) |
| March | 0.950 (0.835-1.079) | 0.938 (0.823-1.071) | 0.927 (0.826-1.040) | 0.889 (0.826 - 0.957) |
| April | 1.085 (0.952-1.237) | 1.084 (0.948-1.239) | 1.074 (0.956-1.206) | 0.945 (0.877 - 1.019) |
| May | 0.907 (0.791-1.041) | 0.901 (0.782-1.038) | 0.920 (0.805-1.051) | 0.987 (0.905 - 1.076) |
| June | 0.826 (0.713-0.958) | 0.819 (0.704-0.952) | 0.847 (0.735-0.977) | 0.944 (0.864 - 1.031) |
| July | 0.902 (0.787-1.033) | 0.900 (0.783-1.034) | 0.914 (0.801-1.044) | 1.006 (0.924 - 1.095) |
| August | 0.950 (0.839-1.076) | 0.944 (0.831-1.073) | 0.958 (0.848-1.083) | 1.004 (0.931 - 1.083) |
| September | 0.957 (0.862-1.062) | 0.949 (0.852-1.056) | 0.981 (0.889-1.081) | 0.972 (0.913 - 1.033) |
| October | 0.965 (0.870-1.070) | 0.961 (0.864-1.070) | 0.973 (0.885-1.070) | 0.936 (0.882 - 0.994) |
| November | 0.920 (0.829-1.020) | 0.917 (0.823-1.020) | 0.941 (0.853-1.037) | 0.98 (0.922 - 1.041) |
| December | 1.095 (0.940-1.276) | 0.878 (0.787-0.980) | 0.902 (0.815-0.998) | 0.933 (0.874 - 0.996) |

[1] Intent to treat (ITT) (primary analysis) and per protocol (PP) (secondary analysis) mixed effects difference-in-difference (DID) Poisson regression specifying measurements made throughout 2016 as 'baseline', with post-2016 measurements as 'post-baseline'; PP analysis only considering houses with SR application rates meeting manufactures specifications ≥75% of the days between sequential blood sampling (designated PP-1).

[2] ITT and PP mixed effects DID Poisson regression specifying measurements made in 2016 up to the first date of intervention deployed in each cluster as 'baseline', with all measurements following that date as 'post-baseline' for that cluster, even for houses that did not enroll until 2017 or later; PP analysis only considering houses with SR application rates meeting manufactures specifications ≥75% of the days between sequential blood sampling (designated PP-1).
