## Supplementary material for "Efficacy of a Spatial Repellent for Control of *Aedes*-Borne Virus Transmission: A Cluster Randomized Trial in Iquitos, Peru": S2. Initial IRB Approved Study Protocol

### `Human Subject Research Protocol

#### Part I – COVER PAGE

1. **HRPP Number:** NAMRU6.2014.0021

2. **Title of Protocol:** Spatial Repellent Products for Control of Dengue Virus transmission in Iquitos, Peru.

3. **Sponsor or other support:** *(List industry sponsor, government support, etc.)*

   **Work Unit Title And Number:**
   **Grant Title And Number:** Spatial Repellent Products for Control of Vector-borne Diseases (Bill and Melinda Gates Foundation)
   **Other:**

4. **Principal Investigator:**
   a. **Name, degree, affiliation, address, telephone number, email:**
      **Nicole Achee**, Ph.D., Eck Institute for Global Health, College of Biological Sciences, South Bend, Indiana, USA 001-202-459-1896,
   b. **Human Subject Protection Education (date completed):**
      19 May 2013
   c. **Curriculum Vitae attached**

5. **NAMRU-6 Lead Investigator:**
   a. **Name, degree, affiliation, address, telephone number, email:**
      Robert Hontz, Lt., Ph.D., Naval Medical Research Unit No. 6, 511-6144-210,
   b. **Human Subject Protection Education, certificate**
      6 March 2013
   c. **Curriculum Vitae, attached**

6. **Associate Investigators:**
   *(For each please provide (1) name, degree, affiliation, address, telephone number and email, (2) institutional affiliation and (3) date human subject protection education was completed)*:
   a. **Name, degree, affiliation, address, telephone number, email:**
      **John Grieco,** Ph.D., Department of Preventive Medicine, USUHS, Bethesda, MD 20814 USA, phone 301.295.3731,
      **Amy Morrison**, Ph.D., M.S.P.H., NAMRU-6, Clinica-Naval, Av. La Marina con C/Trujillo #951, Punchana, Peru, 965-604279,
      **Thomas Scott**, Ph.D., Department of Entomology, University of California, Davis, One shields Ave. Davis, CA95616, USA, phone 530.754.4196,
      **Neil Lobo,** Ph.D., Department of Biological Sciences, University of Notre Dame, Notre Dame, IN 46556 USA, phone 574.631.4155,

   b. **Human Subject Protection Education, certificate**
      Greico: 19 June 2014
      Morrison: 1 Jan 2012
      Scott: 18 Aug 2013
      Lobo: 01 Jan 2014

c. **Curriculum Vitae attached**

7. **Additional Key Study Personnel**
(e.g., individuals listed in the protocol, research coordinators, etc.)*(For each please provide (1) name, degree, affiliation, address, telephone number and email, (2) institutional affiliation and (3) date human subject protection education was completed)*:

a. **Name, degree, affiliation, address, telephone number, email:**
**Helvio Astete**, Department of Virology, NAMRU-6 Iquitos, 377 Callao, Iquitos, Peru, 51-65-600102,
**Steven Stoddard**, Ph.D., UC Davis, Department of Entomology, Assistant Project Scientist, Phone: 530-752-0565, Fax: 530-752-1537,
**Stalin Vilcarromero**, Department of Virology, NAMRU-6, Clinica-Naval, Av. La Marina con C/Trujillo #951, Punchana, Peru, 965-885174,
**Clara del Aguila**, B.S., Dirección General de Saneamiento Ambiental (DIGESA), Calle Alzamora 410, Iquitos, Peru

b. **Human Subject Protection Education, certificate**
Astete: 31 May 2013
Stoddard: 05 June 2014
Vilcarromero: 31 May 2013
Del Aguila: 31 May 2013

c. **Curriculum Vitae attached**

8. **Research Study Site(s):**

a. **Research Study Site(s):** Iquitos, Peru (Annex 1)

b. **Lead Research Institution (name of site with lead or primary IRB responsibility):**
NAMRU-6

c. **Collaborating Institutions (list all institutions):**

University of Notre Dame
University of California, Davis
DIRESA/DIGESA Loreto

d. **Host Country Approval Agency (ies) (list all MoH department/offices, local DoD component/command, etc. as applicable):**
DIRESA/DIGESA

e. **Does a written agreement exist to formalize this collaboration?**

**No:** ☐ **Yes:** ☒ (*If yes, please provide a copy of the agreement with this application)*:
IRB Relying agreements: Notre Dame/NAMRU-6, UC Davis/NAMRU-6, USUHS/NAMRU-6; CRADA: UC Davis and NAMRU-6 (Pending)

9. **Dates of Research** *(Proposed start and end dates):* 15 July 2014 to 14 September 2018

10. **Medical Monitor:**
**Name, degree, affiliation, address, telephone number, email:**
Not Applicable (minimal risk study)

11. **CRADA number:**
Pending

**12. Program Director:**
**Name, degree, affiliation, address, telephone number, email:**
Daniel Bausch, MD, MPH & TM., Head, Virology and Emerging Infections Department, U.S. Naval Medical Research Unit No. 6 (NAMRU-6) Lima-Peru, phone: 511-6144189,.

**13. COMMANDING OFFICER:**
**Name, degree, affiliation, address, telephone number, email:**
Kyle Petersen, CAPT, MC, USN; US Naval Medical Research Unit No. 6 (NAMRU-6), Peru, 511 614-4111,

### Part II – Record of Changes

### Abbreviations

| | |
|---|---|
| AI | Active Ingredient |
| CPD | critical path of development |
| DENV | dengue virus |
| DOD | Department of Defense |
| ED50 | Effective Dose for 50% repellency |
| ELISA | Enzyme-Linked Immunosorbent Assay |
| GIS | Geographic Information System |
| HIN | Household Identification Number |
| HITSS | High Throughput Screening System |
| HRP | Horseradish peroxidase |
| IgG | Immunoglobulin G |
| IgM | Immunoglobulin M |
| IRB | Institutional Review Board |
| IRS | Indoor Residual Spraying |
| LLIN | Long Lasting Insecticide treated Net |
| LTFU | Loss to Follow up |
| MFIR | Minimum Field Infection Rate |
| MOH | Ministry of Health |
| OD | Optical Density |
| PCR | Polymerase Chain Reaction |
| PE | Protective Efficacy |
| RCT | Randomized Cluster Trial |
| RDT | Rapid Diagnostic Test |
| RT-PCR | Reverse Transcription Polymerase Chain Reaction |
| SAE / AE | Serious Adverse Events / Adverse Events |
| SFS | Semi Field System |
| SFS | Semi-field Systems |
| SI | Subject Identity |
| SOP | Standard Operating Procedure |
| SR | Spatial Repellent |
| UIC | Unique Identifier Code |
| WHOPES | WHO Pesticide Evaluation Scheme |

### Part III – Summary of Research in Lay Terms

*(No more than 500 words)*

Dengue viruses are the most medically important arthropod-borne pathogens worldwide, with transmission occurring in most tropical and sub-tropical regions. An estimated 390 million infections occur yearly. Although, there are considerable ongoing efforts to develop a vaccine, vector control remains the only option for reducing DENV transmission and disease burden. The goal of this project is to determine the efficacy of a spatial repellent (SR) product (active ingredient transfluthrin) for reducing contact between household residents and vector mosquitos and as a result reduce DENV transmission. Spatial repellency is used here as a general term to refer to a range of insect behaviors caused by airborne chemicals that reduce contact between people and disease vectors. This can include movement away from a chemical stimulus, and interference with host detection (attraction-inhibition) and/or feeding response.

Protection provided by this product will be measured using entomological and virological approaches, comparing entomological indices will be measured through standard household monitoring of *Aedes aegypti* population densities, while DENV transmission will be measured through door-to-door surveillance for active dengue disease and through serological monitoring for DENV exposure in a randomized cluster trial. We will establish a cohort of 3,400 persons, primarily children 2-12 years of age and adults who have not been previously infected with DENV who will provide annual blood samples when they are healthy, whereas in the same clusters we expect to monitor 5,000-6,000 residents for active dengue disease. The cohort will be monitored for a period of 2 years. The use of spatial repellents has never been tested on a large scale to reduce disease and could change vector control practices worldwide, reducing the amount of chemical insecticides applied and also prevent the development of insecticide resistance.

The project will be carried out in the Amazonia City of Iquitos, Peru, which has a well-established infrastructure for studying urban dengue fever. This trial is the only site, of a multi-center project that is testing SR against dengue fever and is thus part of a larger study with sites in Africa and Asia that will evaluate the impact of SR against malaria. The study, therefore, will generate rigorous evidence, documenting and evaluating the impact of SR products on human infection rates, to be considered and used by academia, industry and public health key stakeholders at the global, regional, national and/or local level and drive efforts to acquire full recommendation of SR products for inclusion in disease control programs.

### Part IV – Scientific Background and Military Relevance

**1. Background:** Dengue is a problem for over 3.97 billion people in 128 countries where the disease is actively transmitted and recent estimates put the number of symptomatic infections around 400,000 per year [Brady, 2012]. A mosquito-borne arbovirus belonging to the family Flaviviridae, the virus is capable of causing dengue fever, dengue hemorrhagic fever and dengue shock syndrome (Gubler and Kuno 1997). The predominant vector of dengue virus, *Aedes aegypti* (L) (Diptera: Culicidae), is a peri-domestic, day-biting mosquito species found throughout the tropical and semi-tropical world. Adult stages are commonly found in secluded areas of a dwelling, such as underneath beds and resting on clothes in the lower portions of closets. *Aedes aegypti* also vectors other arboviral infections including Chikungunya and urban yellow fever. Dengue prevention currently relies entirely on vector control and is focused on routine larval source management and reactive space spray or other adult-focused campaigns [Erlanger et al. 2008].

Two general approaches have been used to control *Ae. aegypti*: (1) elimination or treatment of larval habitats to prevent adult development and (2) insecticidal space spraying to reduce adult populations (Gubler 1989, Reiter and Gubler 1997). Although a mainstay of most *Ae. aegypti* programs (WHO/TDR 2009), spraying is expensive and unless done inside houses it is often ineffective because *Ae. aegypti* rest in places that are not penetrated by outdoor sprays (e.g., closets inside houses) (Reiter and Gubler 1997). In addition, as the scope of dengue continues to grow, new tools will be needed to compliment the limited number of available interventions and/or further optimization of current products will be required to meet public health demands. This includes new paradigms for *Ae. aegypti* control targeting adult mosquitoes (Morrison et al. 2008). The need for spatial repellents for vector control can be highlighted as they are effective when considering outdoor/day/early-evening biting mosquitoes – the area of transmission that long lasting insecticide treated nets (LLINs) and indoor residual sprays (IRS) have low effect. SRs have been demonstrated to show effect against insecticide resistant populations and have the potential to limit the spread and/or emergence of insecticide resistance alleles due to low/no selection pressure when considering non-lethality of effect. SRs, in combination with other interventions, have the potential to demonstrate added protective benefit, especially in areas where traditional LLINs or IRS interventions may not offer full protection or have reached their efficacy limits, especially in areas with residual transmission or areas where elimination is proposed. Control and/or elimination of disease in these areas will require new approaches and this may be where spatial repellency would be most effective [Ogoma et al. 2012; Achee et al. 2012]. Spatial repellents could be offered as stand-alone tools where no other interventions are currently in use; or, most likely, combined with existing interventions to augment efficacy of these other tools (i.e., a combination strategy) thereby tackling residual transmission, managing the spread of insecticide resistance and intervening in areas of the vector life-cycle where other interventions do not reach.

Spatial repellency is used here as a general term to refer to a range of insect behaviors induced by airborne chemicals that result in a reduction in human-vector contact. This can include movement away from a chemical stimulus, and interference with host detection (attraction-inhibition) and/or feeding response. Spatial repellency can be measured and distinguished from other chemical actions, to include contact irritancy and toxicity, in laboratory studies [Grieco et al. 2005; Achee et al. 2009]. Repellents drive mosquitoes away from a treated space. Toxics kill mosquitoes and irritants are compounds that rely on contact between the mosquito and a treated surface to prevent resting. Many compounds exhibit two or more modes of action, but they can be distinguished by the concentration or dose needed to achieve them [Grieco et al. 2007]. Spatial repellency occurs at low vapor phase concentration, contact irritancy requires higher doses and killing requires absorption of still higher levels. In semi-field conditions Phase II (experimentally controlled conditions) mosquitoes in natural environments in Belize [Grieco et al. 2000; Grieco et al. 2007] and Thailand [Ojo et al., unpublished data] do not enter experimental huts that contain a spatial repellent source. Spatial repellents have been demonstrated to prevent human-vector contact using vapor-active pyrethroids. Kawada et al., (2004a, 2004b, 2005) showed both indoor and outdoor protection (i.e., fewer bites) against *Anopheles* spp. and *Culex* spp. when using metofluthrin-impregnated paper strips as compared to untreated sites in field conditions in Lombok, Indonesia. Evaluations of transfluthrin have also shown up to 90% bite protection from *Anopheles* spp. inside houses of Dar es Salaam, Tanzania compared to control after volatizing using a kerosene oil lamp (Pates et al. 2002). Outdoor field trials in North America have demonstrated paper emanators impregnated with metofluthrin to reduce landing rates of *Aedes vexans* by up to 95% compared to pre-treatment conditions (Lucas et al. 2005) and *Ae. canadensis* by 85-100% compared to untreated controls (Lucas et al. 2007); both species are nuisance biters and incriminated in vectoring West Nile Virus. These mosquitoes include both major genera (e.g. *Anopheles spp.* and *Aedes spp.*) as well as a range of insecticide susceptibility statuses. There is also evidence from Phase III studies that spatial repellents can impact pathogen transmission. A proof-of-principle study showed that communities (village clusters) with high coverage (90%) of coils had lower biting pressure from mosquitoes and reduced malaria transmission by 60% [Syafruddin et al., unpublished data]. Replication and extension of these studies will be the core of the work outlined in the current proposal. In particular for the dengue site, our objective is to provide proof-of principle that SRs can reduce dengue transmission.

Over the past several years, formal national and international meetings have been convened to bring together academics, industry and global public health experts, including representatives from the WHO and the WHO Pesticide Evaluation Scheme (WHOPES), to discuss the role of SR in the reduction of arthropod-borne diseases. A critical aspect of these meetings and subsequent efforts has been to establish a critical path of development (CPD) for these products based on expert advice. This includes measures related to scientific, regulatory and social parameters. In part, these criteria outline the endpoints of a target product profile (i.e., optimum product characteristics) for a spatial repellent product. A major hurdle in the CPD is the generation of sufficient epidemiological data demonstrating significant protective efficacy (PE) to influence policy-makers to recommend the incorporation of spatial repellents into current multi-lateral disease control programs confidently. Realizing this vision demands credible demonstration of protective efficacy (PE) against new human infections and estimates of SR PE based on vector bionomics, intervention coverage and/or disease transmission rates to define optimum patterns of use.

This protocol is designed to measure PE against DENV transmission in a single site. It is part of a larger multi-country trial testing PE for Malaria transmission with multiple sites in Asia and Africa. For this protocol, all field experiments proposed will be carried out in the Amazonian city of Iquitos, Peru, where dengue is endemic. During 1990, this town was the center of the first (laboratory-confirmed) dengue epidemic in Peru (serotype I) (Phillips et al. 1992, Hayes et al. 1996, Forshey et al. 2010). Major epidemics of dengue also occurred in 1995 (serotype 2), 2002, 2004, 2006-2007 (serotype 3), 2008-2010 (serotype 4) and most recently from December 2010-March 2011, Iquitos experience the largest dengue outbreak in its history with nearly 20,000 reported cases (confirmed and probable) of which 2,042 presented warning signs, 104 required intensive care, and 14 fatalities were documented associated with Lineage II of the Asian-American strain of dengue 2 (Forshey et al. 2009b, Forshey et al. 2009a, Forshey et al. 2010, Morrison et al. 2010, Durand Velazco et al. 2011, Mamani et al. 2011, Rodriguez Ferrucci 2011).

Although not a specific objective of this project, an overall goal is to allow for official recommendations (or not) from WHO or in- country specific authorities of presently available spatial repellent products and the eventual evaluation of these products that are not being regulated, thereby increasing quality, overall use and a consequent reduction in disease. The use of a standardized core protocol will facilitate this process. In essence, we will generate data that can be reviewed by WHO to recommend (or not) the use of SR products. Formal recognition of a spatial repellent paradigm for disease prevention will 1) facilitate the discovery and development of such products by industry manufacturers, 2) drive the expansion of new mosquito control approaches and 3) allow for the recommendation of existing SR tools that can provide substantial impact on disease transmission worldwide – something that is vital in the fight against arthropod-borne diseases. This translates to protection from a broad range of diseases and disease vectors that could extend beyond dengue-endemic countries.

#### 2. Military relevance *(Relevance of research study to operational/Navy medicine)*:

The cosmopolitan distribution of *Aedes aegypti* and its close association with anthropogenic environments enables it to successfully vector several arboviruses, most notably dengue and Chikungunya viruses (Delatte et al. 2008). *Aedes* larvae inhabit small, localized habitats (i.e., artificial containers, tires, water storage containers, disposal plastic containers) thus are difficult to control before adulthood (Service 1992, Delatte et al. 2008). Of arboviral diseases, dengue virus (DENV) infections cause the most human morbidity and mortality worldwide (Monath 1994, Kuno 1995, Gubler and Kuno 1997, Gubler 2002). It is estimated that 2.5-3.0 billion people are at risk of infection each year. Millions have been infected during recent epidemics. Beyond the direct impact on afflicted individuals, urban dengue epidemics overwhelm public health systems and destabilize societies (Gubler 2002b, a). Dengue occurs throughout the tropics where deployed troops are at risk. Furthermore, incidence rates have steadily increased since the 1950s (Rigau-Perez et al. 1998) with severe economic consequences for endemic countries that negatively impact economic development and contribute to political instability.

At present, reducing the density of *Ae. aegypti* is the only option for prevention and control of DENV infections in human populations. Even when licensed vaccines or chemotherapeutics become available for the treatment of this disease, we expect vector control to remain as a necessary part of the most effective integrated dengue prevention programs. Successful dengue control programs will require innovative strategies that include targeting adult mosquitoes (Morrison et al. 2008). Application methodologies implemented during this project may provide significant insight into the effectiveness of novel spatial repellent products directed against adult stages of mosquitoes near US troop encampments and urban environments. Other vector species, such as *Anopheles* spp. and *Culex* spp., may be similarly affected. Our study builds on a strong

Department of Defense (DOD) commitment to operational research on spatial repellent strategies conducted in Iquitos and other locations.

### Part V – Objectives or Hypotheses

The **primary objective** of the study is to quantify the protective efficacy (PE) of a spatial repellent product in reducing the incidence of dengue infection in a large-scale randomized cluster trial (RCT) in the city of Iquitos, Peru. Project objectives will be met through pilot studies carried out between September 2014-June 2015 and a RCT conducted between September 2014 and September 2017.

#### 1. Primary objectives:

a. Compare incidence of dengue in SR treated and control household clusters, receiving standard entomological surveillance and control procedures by the local ministry of health. **The null hypothesis is that there is no difference in measured endpoints (dengue seroconversion and clinical attack rates) between intervention and control arms.**
b. Confirming and measuring the entomological correlates of reduced infection. (e.g., a reduction in adult densities, mosquito infection and parity rates).
c. Determine if the indoor:outdoor ratio of adult *Aedes aegypti* collected is lower in intervention compared to control clusters.

#### 2. Secondary objectives:

a. Evaluate the adoption, maintenance, and sustainability of the SR intervention in the trial clusters.
b. Determine the impact of the SR intervention use on population densities of non-target mosquito species.

### Part VI – Study Design and Experimental Methods

#### 1. Study type:

The study will be a prospective, cluster-randomized, participant and observer-blinded, placebo controlled trial in a site endemic for dengue to measure the impact of a spatial repellent (SR) product on new dengue virus infections. Clusters of households, containing 100-120 residents testing negative, or positive for antibodies against single DENV, will be selected from two separate areas of 13 of 34 Ministry of Health (MOH) zones (Fig. 1). All participating houses in each cluster will be monitored entomologically using standardized *Aedes aegypti* surveys immediately before deployment of the SR intervention and at 2 periods after the intervention is in place. A post-intervention survey will also be conducted 1 month following the last subject follow-up. Entomological surveys will include monitoring of both *Ae. aegypti* population densities, age structure, location (indoors versus outdoors), and mosquito infection rates. DENV transmission will be assessed by monitoring active disease rates during the study and by comparing seroconversion rates from baseline (pre-intervention) and follow-up (post-intervention) serum samples between intervention and control clusters.

**2. Study population:** We will address our research objectives in the context of the following research activities. (Annex 1). Also see Figure 2 for flow chart.

**Baseline serostatus to identify longitudinal cohort participants (n=9,000):** Our objective is to recruit longitudinal participants whose baseline serostatus against DENV is seronegative, or positive to a single serotype, indicating they have only had a primary dengue infection. For this reason our primary target group will be children 2-12 years of age who have a higher probability of no previous infection with DENV. More importantly, this age group typically is less mobile than adults spending more time at home where the SR intervention would presumably provide protection. To keep house cluster sizes to a minimum (for logistical reasons) we will also screen all willing adult residents by IgG ELISA to identify seronegative individuals who would then be included as longitudinal cohort participants. We estimate that we would need to screen up to 9,000 residents to achieve a total longitudinal cohort size of 3,400 which with loss to follow-up of 28% would yield 2,400 participants that could be used for seroconversion estimates (see experimental methods below for rationale).

| Individual Level (Written consent-ICD-Longitudinal Cohort; Information Sheets: Explanation of Study and Blood Samples and Febrile Illness Visits) | |
|---|---|
| Inclusion Criteria | Exclusion Criteria |
| • ≥ 2 years of age<br>• Plans to stay in study area for minimum of 12 months<br>• Resident of household or frequent visitor (~20% of day hours in house/mo) | • Less than 2 years of age<br>• Temporary visitor to household<br>• Plans to leave study area within next 12 months |

**Longitudinal cohort (n=3,400):** Follow-up serum samples will be requested from eligible participants after screening at the same time the SR products are installed (May-July 2015, time 0), and 12 and 24 months later, primarily during the months of May-July. At each annual bleed, we will recruit participants who have stayed in the study areas under febrile surveillance to participate in the longitudinal portion, replacing individuals lost to follow up, to ensure that the study power is maintained. Identification of seroconversions in participants experiencing 3$^{rd}$ or 4$^{th}$ DENV infections can often be difficult requiring that data be excluded. By restricting our population to seronegative or monotypic we maximize the probability of generating interpretable data. Seroconversion rates will be compared between intervention and control clusters.

**Febrile surveillance (~9,000 residents):** All households on the blocks where longitudinal participants are recruited will be invited to participate in a household census and allow health workers to visit their homes 3x/week to ask if any residents have a fever. When an individual is identified with a fever, in a separate consent process they will be invited to provide acute and convalescent (14-21 days later) blood samples and receive daily medical exams (physical exam, temperature, vital signs, and tourniquet test) for the course of their illness. While we will be focusing on dengue virus transmission for this study, we also plan to take advantage of the labor-intensive door-to-door surveillance study design to address the objectives of a concurrent study of respiratory pathogen transmission in the region (NMRCD.2010.0010). Human infection by a range of pathogens (such as dengue virus or influenza virus) can lead to non-specific disease presentation, such as fever, headache, and malaise. Thus, arbovirus (dengue) and respiratory pathogen (influenza) surveillance can be integrated seamlessly, maximizing personnel and infrastructure and minimizing inconvenience for the participant. To achieve this alternative objective, during the household febrile surveillance consent process we will obtain permission from the head of household to invite residents with febrile illness compatible with respiratory virus infection, identified during 3x/week home visits, to participate in the separate study. Respiratory samples will be collected from participants under a separate consent process. Confirmed dengue cases will also be invited to participate in separate protocols that include feeding mosquitoes on infected people (NAMRU-6.2011.0002), collection of PBMCs for immunological studies (NMRCD2005.0009 and NMRCD2007.2007), or evaluation of novel rapid diagnostic tests (NAMRU-6.2014.003). Incidence of dengue disease and the symptomatic to asymptomatic case ratio will be calculated between the intervention and control clusters.

| Household Level (Consent without written documentation, Information Sheet – Explanation of Study) | |
|---|---|
| Inclusion Criteria | Exclusion Criteria |
| • Adult head of households agrees to health visits and census<br>• Individuals spend a minimum of 4 hrs per week during the daytime hours or sleep in the house. Temporary residents can be included | • Households where study personnel identify a security risk (i.e., site where drugs are sold, residents are always drunk or hostile)<br>• Sites where no residents spend time during the day (i.e. work 7d a week outside the home) |
| Individual Level – Febrile cases (Written consent) | |
| • Individual who spends a minimum of 4 hours per week within the household.<br>• ≥ 2 years of age<br>• <u>Fever at the time of presentation or report of feverishness within the previous 24 hours</u> | • Less than 2 years of age<br>• Individuals who spend have spent less than 4 hours in the household during the week prior to illness. |

**<u>Entomological Monitoring (Pilot ~100 households; Efficacy trial ~3,380 households)</u>:** Entomological monitoring will be conducted for both the pilot and the RCT. For the *Pilot study* full pupal/demographic surveys will be carried out in households at the beginning and end of the nine-week monitoring period. In addition adult mosquito collections will be carried out up to 2x per day. For the *RCT* monitoring, surveys will be conducted at regular intervals in each cluster.

| Household Level (Consent without written documentation, Information Sheets-Explanation of Study, Entomological Surveys) | |
|---|---|
| Inclusion Criteria | Exclusion Criteria |
| • Adult head of households agrees to surveys | • Properties where study personnel identify a security risk (i.e., site where drugs are sold, residents are always drunk or hostile) |

**<u>SR Intervention (Pilot ~100 households per replicate; Efficacy trial ~3,380 households)</u>:** All households will receive one sheet for every 8m$^2$ area (i.e. a household 100 m$^2$ would require 6-8 sheets) that will be hung from the ceiling. Sheets treated with transfluthrin will be placed in the intervention clusters whereas the control clusters will receive untreated placebo cards. Field personnel will be blinded. Sheets will be collected and replaced at 3 week intervals.

| Household Level (Consent without written documentation, Information Sheets-Explanation of Study, Transflutrin Pamphlet) | |
|---|---|
| Inclusion Criteria | Exclusion Criteria |
| • Adult head of households agrees to SR deployment and to provide access to team member at 3 week intervals to change cards. | • Properties where study personnel identify a security risk (i.e., site where drugs are sold, residents are always drunk or hostile) |

**3. Number of subjects to be enrolled/or charts/records to be reviewed/number of biological specimens to be used in this study:** Pilot studies: ~100 households
RCT: ~3,380 households (ask for permission for up to 5,000 households); Longitudinal participant screening and active surveillance (up to 9,000); Longitudinal participants eligible for follow-up samples, ~3,400 (we ask for permission up to 5,000 individuals since if we identify more seronegative individuals than expected it will only improve the power of the study).

**If only biological specimens will be used, please complete the following information:**

**a) Biological specimens obtained under a NAMRU-6 IRB approved study; if so, please provide the name and number of the protocol.** N/A
**Specimens to be used have a future use authorization:** Yes

**b) Biological specimens obtained from clinical care/collaborative efforts; if so, please provide the source.** N/A

**4. Duration of individual subject participation** *(State, not Applicable for chart/record reviews)***:**
Longitudinal and febrile surveillance: 2 years, but with an extension to 5 years (the study is currently slated for 2 years, but we put a clause in saying if we need additional data we will ask if we can continue). A amendment would be submitted before an extension took place. For example, we may try new product formats in subsequent years or need to collect data for additional years if study endpoints are marginally significant or close to significance.

**5. Age range of subjects** *(adults and/or children)***:** $\geq$ 2 years of age.

**6. Types of populations involved,** *please describe (pregnant women, lactating women, neonates, military, dependent employees, individuals with economical, social, physical or cognitive disadvantages, individuals sedated or traumatized, healthy volunteers)***:**

This study aims to enroll all households located in the study area. There will be no exclusion criteria related to gender, ethnic or racial category. Based on previous studies by UC Davis/NAMRU-6, we expect that 52% of the study population will be female. We have also found that adult females are more willing to participate than males. In similar previous studies all participants were Peruvian citizens, Spanish speakers, and of mestizo ethnicity (mixed Amerindian and white). The health status of this population will be mostly healthy, however, we expect that pregnant woman, some individuals with physical and mental disabilities, and individuals with other acute and chronic illnesses and active duty military will be living within the study area; all will be residents of Iquitos, Peru. Since all of these groups are at risk of contracting dengue it would be inappropriate to exclude them if they wish to participate.

Studies requiring blood samples have been limited to individuals $\geq$ 2 years of age. We excluded younger children because of the inherent difficulties obtaining blood from small children. Pregnant women will be present on the study blocks and allowed to participate in all aspects of the study, because they represent a group at risk for dengue. Little information is available on the effects of DENV infection on fetuses and the procedures for our study do not represent additional risk to the mothers. Children are a significant risk group and represent the population with the highest number of susceptible individuals, thus their inclusion is essential to understand the overall transmission dynamics of dengue. Although active duty military members might be living on the study block, their superior officers would have no knowledge of their participation. They are also at risk for dengue and exclusion in this context would be inappropriate and discriminatory.

**7. Will subjects be compensated or paid an incentive for participating?**
**If YES,** *please indicate how much, the form of payment and whether this is a reimbursement for expenses incurred or an incentive.*

We will provide a small gift (2 bags of powdered milk) of food or vitamins (30 vitamin pills) as recognition of providing a blood sample, which we consider an appreciation for participation. Although these items may provide a small incentive to participate in the study, they are only mentioned after a procedure has been completed and are not used to actively recruit participants. Although the risks of the procedures involved are of minimal risk and discomfort, it is only appropriate that participants, especially children, receive a small token for their participation. These items have never represented undue inducement, because many residents, independent of socio economic condition, have elected not to participate in previous research of a similar nature in Iquitos, when similar gifts have been offered.

Mention of this appreciation is not included in the consent or assent forms in order to insure proper focus of the informed consent process. That is, we want potential individuals to focus on the information provided during the consent process rather than be thinking about what they will receive at the end of the project. We also feel that it is important to reduce the perception of “buying” people’s blood.

**8. Will treating physicians, clinicians, or researchers be compensated or paid an incentive for referring or enrolling subjects?**
**If YES***, (please explain)*:
The investigators will not be compensated or provided other incentives for enrolling participants in the study. This project is funded by the Bill and Melinda Gates Foundation which has no personal or financial interest in this research. A partnership agreement with SC Johnson, Inc., the manufacturer of the transfluthrin sheets, is in place for product support throughout the study.

**9. Experimental procedures:** *please explain*

The proposed studies will be conducted in urban sections of Iquitos City in the Amazon region (73.2’W longitude, 3.7°S latitude, 120 m above sea level) in the Department of Loreto, northeastern Peru. The site has been described in detail previously (Hayes et al. 1996, Getis et al. 2003, Morrison et al. 2004a,b, Rocha et al. 2009, Morrison et al. 2010). The current population is approximately 380,000 distributed among 4 districts: San Juan, Maynas, and Punchana running from South to North and Belen on the East (INEI, 2008). Pilot studies examining only entomological outcomes will be carried out on selected blocks and include up to 1,000 households for experiments from 2-9 weeks of duration.

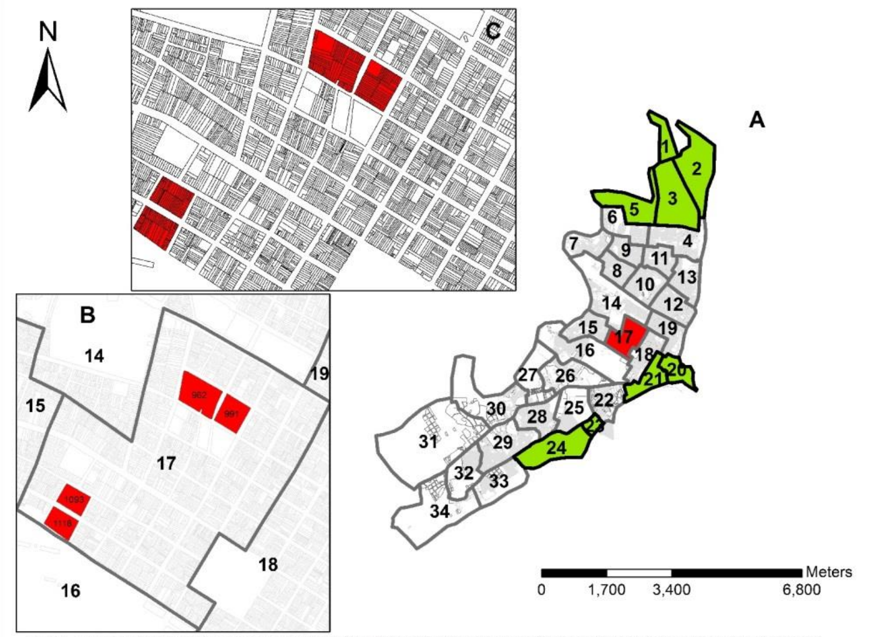

Figure 1. Description of cohort rcrutitment scheme. Panel A: City of Iquitos, Peru divided into 34 Ministry of Health Zones. Green indicates zones that will not not be included, red indicates location of panel B a close up of zone 17, with a single block where recruitment would be initiated shown in panel C.

The proposed RCT will be carried out in 26 MOH zones covering all 4 districts (Fig. 1A). Based on previous cohort studies conducted in Iquitos, the number of houses per block ranged from 20-88 (average = 32) and the number of residents per household ranged from 0-114 residents (average=6.6). In our experience, 80-90% of houses on a block are willing to participate in some study activity (i.e., wellness visits and/or entomological surveys) and of these individuals 30-40% of residents will agree to provide longitudinal blood samples when they are healthy. Our aim is to establish a cohort of 100-140 longitudinal participants (seronegative or evidence of a single previous infection with DENV) in each of 13 MOH zones that are not effected by season flooding which results in a highly unstable human population (migrate to higher ground annually) and also lower DENV transmission rates (eliminated zones are shown in Figure 1 in green). So if we assume that there are 6.6 persons per house, 90% houses participate in someway, 40% of the residents in those houses are willing to provide longitudinal samples, and 33% of these are children, then we would need to visit approximately 130 households to recruit 100 children. Thus, assuming 40% of residents participate we would screen up to 9,000 residents to achieve a total cohort size of approximately 3,400. Allowing for a loss to follow-up of 28% we expect paired samples from approximately 2,400 participants for seroconversion estimates. Our target population will be children 2-12 years of age because they are most likely to be seronegative and they are also less mobile than adults and more likely to benefit from the protective efficacy afforded by the SR intervention. To maximize cluster participants however, we will screen adults at baseline to include any who are negative by IgG-ELISA. Therefore in each of 13 MOH zones, we will select two blocks distant from one another where recruitment will begin (Fig. 1B). Adjacent blocks would be invited until the target of 100 children was reached (Fig. 1C, shows lots on each individual block).

##### a. Field work:

We will address our research objectives in the context of (1) preliminary pilot experiments and (2) the RCT. We present sections on entomological monitoring methods, household census, time in house monitoring and Monitoring of DENV transmission and a description of the SR intervention and deployment.

**Entomological Surveys (see Entomology Information Sheet and Annex 2):** Our research team has developed a standard pupal and demographic survey over the last 13-years by 6 two-person collecting teams using, a survey methodology based on Focks et al. (1993) described previously (Getis et al. 2003, Morrison et al. 2004a, Morrison et al. 2004b, Schneider et al. 2004). Included in our pupal-demographic survey is an adult mosquito collection carried out simultaneously using a Prokopec Aspirator for indoor and outdoor capture (Vazquez-Prokopec et al. 2009). Permission is obtained from adults in the household at each visit. Three information sheets will be supplied to residents during the first visit along with an explanation of the project (see section VIII.1.b).

Age Structure assays: Up to 30 adult *Ae. aegypti* per day will be dissected to remove ovaries. The physiological state of these mosquitoes will be scored as unfed, blood fed (fresh, old), gravid. Ovaries will be scored as nulliparous, gravid or parous.

Household Questionnaires: Prior to performing entomological and serological surveys, we will administer a household questionnaire to the head of the household. Information will be obtained on the number, age and sex of occupants, dimensions of the property, housing materials, method of cooking, water use patterns, type of sewage disposal, and insecticide use. The number of occupants and dimensions of the property are essential to calculate mosquito indices using persons or hectares in the denominator. The remaining information will be useful to control for confounding and account for differences in SR efficacy.

**Household Census:** A census form (Annex 3) listing all persons residing or regular visitors will be recorded. Each household will be assigned a unique geographic code (HIN) and each person appearing on the census will be assigned a participant code based on the HIN. This allows for field personnel carrying out febrile surveillance to monitor all members of each household. Individual participants are also assigned a unique numeric participant code by our database that links to the participant code. Therefore each participant will have a unique participant identification number but can have multiple participant codes that are defined as primary or secondary residence and include an estimate of time spent in each location. For subsequent visits census forms are generated from database (Annex 3 – Sample Census Form) and movement of individuals in and out of their households are monitored using a Register of Withdrawl and Address Change Form (Annex 3).

**Time in House:** As part of the initial household census, 1x per month each resident will be asked to estimate the time spent at the residence during daytime hours for weekdays and weekends separately (see Annex 4). This will allow us to control for difference in household exposure to infected mosquito bites among participants.

**Monitoring of DENV Transmission**.

The true impact of the SR vector control intervention will be a reduction in DENV transmission. This will be measured in 2 ways: through serotype-specific seroconversion rates in longitudinal cohort participants providing blood samples at baseline, 12 and 24 months and by identification of active dengue cases through febrile surveillance.

Febrile illness capture. This will be implemented at the initiation of the study and will continue for the duration of the study. Public health workers employed by the study will visit each house three times per week. They will record households where no one is home and record any subjects in the house who have febrile illnesses. They will also record changes in census data, noting individuals who move away and individuals who move into the study area. When a participant reports a fever, study personnel will take their temperature with a thermometer and administer the informed consent process for dengue diagnosis. If the febrile subject meets the inclusion criteria described above and provides consent (parental consent and assent for minors) the project physician will be informed, a brief medical exam (vital signs, tourniquet test) and medical history (see Annex 5 -Dengue Case Investigation Form that includes vaccination and dengue history and lists signs and symptoms) will be performed, and an acute phase blood specimen obtained. If necessary the project physician will evaluate the participant and daily medical exams will be carried out for the course of the illness (usually 3-7 days). If medical exams yield suspicion of DHF/DSS patients will be transported to a local public hospital where their treatment for dengue is free. Ill subjects will be referred to

LRHD clinics or hospitals for treatment. Convalescent blood samples will be requested at the participants home 14-21 days after the acute sample. Virus isolation will be attempted from acute samples, and both acute and convalescent samples will be tested for anti-dengue antibodies by IgM. In addition, if the participant has a febrile illness with respiratory symptoms such as cough, sore throat, or runny nose, they will be invited to participate in a study of respiratory pathogens circulating in the region (NMRCD.2010.0010). A separate informed consent process will be conducted under that protocol if the participant is willing to provide respiratory samples (nasopharyngeal swabs). Full details of sampling and laboratory procedures are available in the NMRCD.2010.0010 study protocol.

Blood Sample Collection. Blood samples for both the longitudinal and febrile surveillance components of the study will be collected by venipuncture or fingerstick procedures using standard aseptic techniques. An experienced phlebotomist (or study physician) will take the blood sample from an antecubital vein. For the longitudinal measuring of DENV-neutralizing antibodies, up to 5 ml (1 teaspoon) of whole venous blood will be obtained from each volunteer. For febrile participants, up to 7 ml (1.4 teaspoons) of whole venous blood will be obtained. Blood will be collected in one Vacutainer® collection tube (red top) without anticoagulant. If venipuncture is unacceptable to a study subject, the finger prick method will be used for collection of a person's first sample. Providing each participant with a choice of methods will increase compliance and reduce the drop-out rate during subsequent collections. For finger pricks, we will use a BD genie safety lancet and microtainer tube system. After cleaning a finger with 70% alcohol, the sterile lancet will be used to the puncture skin. The finger will be squeezed by the phlebotomist forming a large drop of blood which is held over the microtainer tube. Capillary action draws the blood into the 1 ml tubes. Sera will be separated by centrifugation at 2500 rpm for 10 minutes at $4^{0}$C, transferred to cryo-vials and stored at $-20^{0}$C.

#### SR Description and Deployment

The SR product will be evaluated pending confirmation of minimum performance specifications (**Annex 6,7,8**) against target vector species according to WHO guidelines for efficacy testing of spatial repellents [WHO 2012] to include:

- Effective dose for repellency ($ED_{50}$ and $ED_{95}$) established in a High Throughput Screening System (HITSS) laboratory assay [Grieco et al. 2005; Achee et al., 2009];
- Repellency for a given protected area ($m^2$) established in semi-field systems (SFS);
- AI registered by a stringent regulatory authority;
- Formulation to support constant evolution (duration) of a minimum effective dose throughout the area proposed for protection during expected time of exposure to infective bites. The duration of effect of the specific SR intervention product has to be determined so that constant dispersion of the SR AI can be calculated for controlled product use.

One SR product will be used at all sites (**Annex X**). If the formulated product is not registered in the study area an experimental use permit will be obtained before the study begins. Proprietary protection, for evaluation of AI and/or product formats still under development, will follow industry specifications.

The intervention will be applied in houses delineated into geographical clusters based on balancing of study arms (demographics, existing interventions, baseline mosquito exposure). Individual clusters within the site will be randomly assigned to treatment or control arms. Each cluster will be assigned to receive either active repellent product in its standard formulation and duration of activity or a placebo containing no chemical repellent and being otherwise identical to the active product. All households within a cluster will have the same treatment. All houses enrolled in the study will be assigned a unique code to correspond with epidemiology and entomology surveys to facilitate the linkage of data during the intervention.

Industry partners will manufacture, package and deliver SR products according to site-specific sample size and objective demands. A preparatory time period (ramp-up phase) will be factored into the study to facilitate product acquisition in-country and distribution within clusters. Active and placebo product, formulation and packaging will be indistinguishable. Only the study monitor and industry partner will have access to the code sheet that identifies the package as an active repellent or placebo. Existing national standard vector control interventions (i.e., bed nets, fogging, etc.) will continue without interruption in study clusters throughout the trial.

Applications of Spatial Repellent Product (see Spatial Repellent information sheet)**:** Spatial repellent products will be placed inside consented homes at the start of the intervention trial according to manufacturer's specifications. Application (at initial and replacement at 2 week intervals) will be conducted by a dedicated SR product monitoring team.

Product Management**.** Each SR product received directly from industry partner(s) will have a Unique Identifier Code (UIC). Industry partners will ensure that the listing of UICs (identifying the contents as having an active ingredient (AI) or blank/placebo is accurate and kept strictly confidential. All consenting households within the intervention arm (study cluster) will be assigned active products, with all consenting houses within the control arm (randomly chosen) receiving blank/placebo.

Product Monitoring for Compliance**.** An individual SR product monitoring team will be assigned several houses for monitoring product compliance. Monitoring will include documentation of product disbursal and replacement according to predetermined schedule (based on product life), collection and documentation of damaged product, used product, packaging and/or verification that the UIC of used product matches that assigned to the HIN. Mismatching UIC and HIN will be promptly reported at the local level of protocol management.

The monitoring team will record compliance at time of product replacement. The inspector will record the household as "compliant" if SR product is properly positioned at time of inspection. If the inspector finds deviation from prescribed application, the household is recorded as "non-compliant" and a report detailing the nature of non-compliance is submitted on the same day. The product monitoring team will also record product integrity at time of replacement. This will include discolor and/or damage. This information will be recorded using smart phones in real time.

Product compliance summary reports will be produced and reported to industry partner at end of trial. The report will summarize compliance in the distribution, and integrity of product to guide marketing strategy and ensure end-user compliance and acceptability.

##### b. Data collection:

In this section we describe the schedule of activities described in the previous section and refer to data collection forms.

Study Schedule

1. *Pilot Study*. Small trials will be carried out for 9 weeks between October 2014-April 2014.
   Baseline assessments/procedures
   - Trial blocks will be selected based on historical entomological indices obtained in previous entomological studies conducted in Iquitos.
   - House visits, distribution of information sheets and study explanation.
   - Complete Pupal/demographic entomological survey.
   - Deployment of SR intervention on designated study blocks.

   Ongoing assessments/procedures
   - Adult mosquito collections using a Prokopack aspirator will be carried out inside and outside each household up to 2 times per day, 1 week prior and up to 3 weeks to SR product deployment and removal, respectively.
   - Every 3 weeks, SR products will be replaced and rotated to untreated houses. Note intervening housesholds will receive placebo sheets.
   - Another pupal demographic survey will be conducted at the conclusion of the trial.

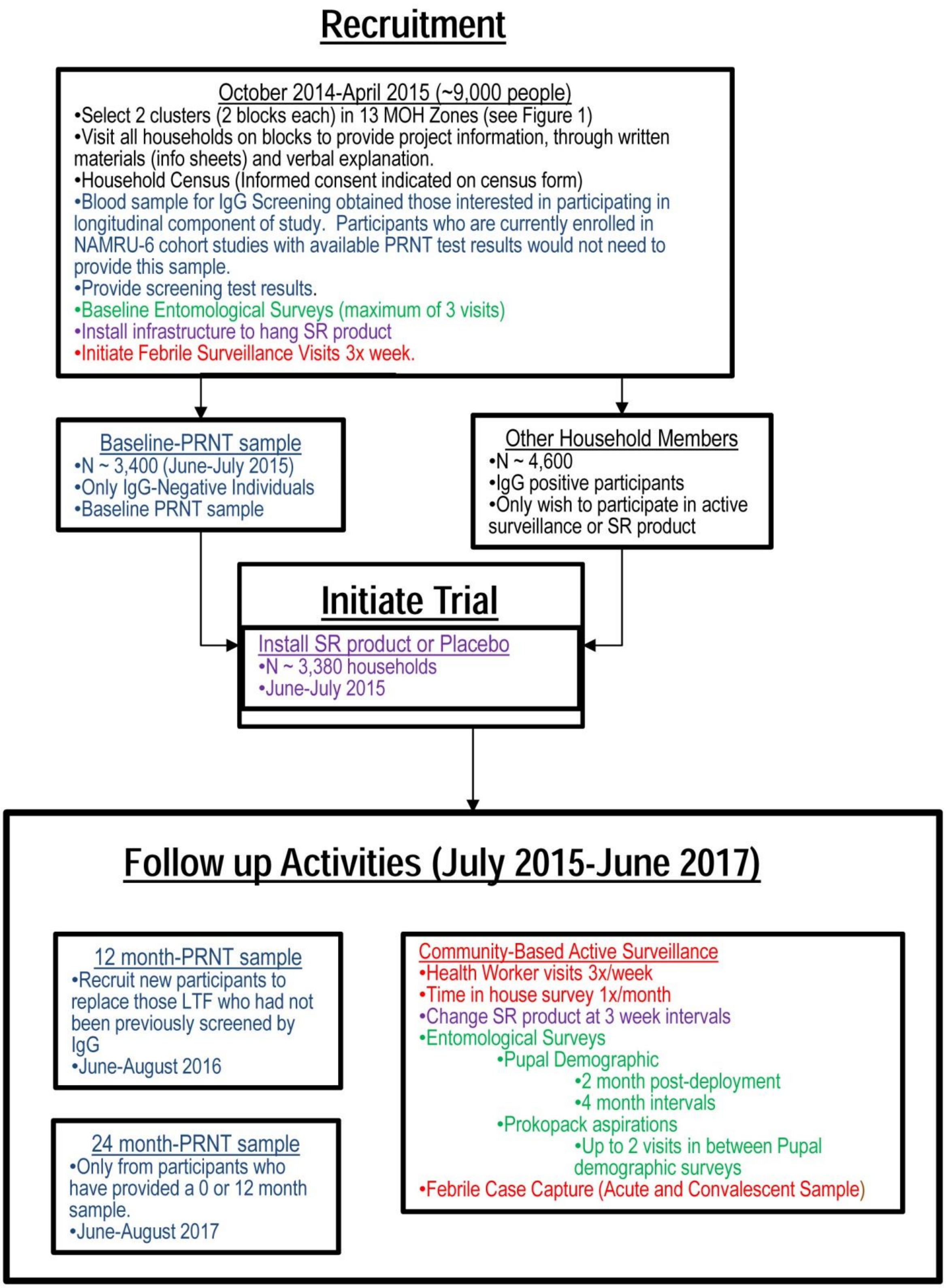


**Figure 2. Flow chart showing RCT activities.**

2. *RCT.* The trial will be carried out from June 2015 to June 2018, but pre-intervention activities could begin as early as October 2014. Data analysis is expected to take until December 2018.

Pre-intervention assessments/activities

- House visits, community meetings if necessary, and distribution of study information.
- Pupal/Demographic Survey and adult mosquito monitoring (2 month intervals).
- Carry out Household Census
- Informed consent procedures (documentation for surveillance, entomological monitoring, and SR deployment; written consent for longitudinal blood samples)
- Time in house surveys
- Assign unique household codes from GIS maps of the study area
- Randomize clusters (i.e., block(s) of houses within a single zone) to receive either active or placebo treatment using random number generator.
- IgG Screening Blood sample for those interested in participating in the longitudinal cohort.
- Determine SR placement

Baseline assessments/procedures

- Informed consent procedures (documentation for surveillance, entomological monitoring, and SR deployment; written consent for longitudinal blood samples)
- Baseline PRNT blood sample.
- Pupal/demographic entomological survey.
- Deployment of SR intervention cards with baseline entomology survey.

Ongoing assessments/procedures

- 3 visits from project health worker per week to ask adult family members if anyone in the household has a fever or had a fever within the previous 5 days.
- SR cards will be removed and replaced at 3 week intervals by study personnel.
- Time in house surveys 1x per month
- Follow up blood samples 12 and 24 months post-baseline sample for longitudinal participants.
- Pupal demographic surveys carried out at 2 month post deployment then at 4 month intervals for the remainder of the project (24 months).
- Adult mosquito collections using Prokopack aspirator in a sample of study households at 1 week intervals (as part of pupal demographic surveys when performed).
- Febrile illness capture and monitoring.

Data Collection Formats

- **Geographic Information System (GIS) for Iquitos:** All household data will be managed in a GIS developed for the city of Iquitos using ARC/GIS software (ESRI, Redlands CA), which allows visualization of different *Ae. aegypti* indices across different scales of time and space, and management of project activities.
- **Data collection Forms:** We have a standardized form for pupal demographic surveys and adult mosquito aspirations that has been in use for many ongoing NAMRU-6 projects. They are included in this package. With the exception of our GIS code the information on these forms has no personal identifiers.
- **Smart Phone Monitoring of Card Status:** Samsung Android telephones are being used to keep track of visits to replace SR cards.

##### c. Laboratory diagnosis:

All blood samples collected in the longitudinal arm of the study will be assayed by Plaque Reduction Neutralization test using standard NAMRU-6 protocols (Comach et al., 2008, Morrison et al. 2010) to identify seroconversions between paired samples from the same individual. PRNT assays will also be conducted on acute and convalescent samples from study participants manifesting with febrile illness detected through surveillance activities. Acute-phase serum samples will be tested for DENV infection either by virus isolation in cell culture or by detection of viral RNA by reverse transcription polymerase chain reaction (RT-PCR). Both acute- and convalescent-phase samples will be screened for DENV IgM antibody by IgM-capture enzyme-linked immunosorbant assay (ELISA). If appropriate, acute and convalescent samples will be tested by DENV IgG ELISA. Febrile episodes will be classified as DENV infections based on the isolation of virus, RT-PCR, IgM serology (elevated IgM antibody titers [≥1:100] in the acute sample, convalescent sample, or both), or IgG antibody serology (four-fold rise in titers between acute and convalescent samples).

**Plaque Reduction Neutralization Test (PRNT).** A modified protocol of Morens *et al.* (1985) will be followed. Test sera will be diluted two fold in media (EMEM + Pen./Strep.) from 1:40 to 1:640. Two hundred μl media containing 40 to 80 PFU of assay virus will be mixed with 200 ul diluted test serum and then incubated at $4^0$ C for 15 hours. In triplicate, 100 ul virus-serum mixture will be added to 0.5 ml media containing 1.5 x $10^5$ BHK21 cells and then added to a well of a 24 well tissue culture plate and incubated at $37^0$ C with 5% $CO_2$ for 3 hrs. The cells will then be overlaid with 0.5 ml of overlay media (0.6% Carboxymethyl Cellulose, MEM w/o Phenol Red, 10% FBS, 0.075% $NaHCO_3$ and Pen./Strep.) and incubated at $37^0$ C with 5% $CO_2$ for 5 days. The media will be removed, and the cells rinsed with $H_2O$ and stained with 0.5 ml/well stain solution (0.1% (w/v) Naphthol Blue Black, 1.36% (w/v) Sodium Acetate, and 6% (v/v) Glacial Acetic Acid) for 30 min. The stain will be removed and the plaques will be counted. The results will be expressed as the serum dilution, determined by probit analysis that reduced the number of plaques by 70% compared to that of normal human serum at the same dilution.

**IgM-Capture ELISA.** Dengue-specific IgM antibody titers will be determined by an IgM-capture ELISA adapted from published protocols (Innis et al. 1989). Briefly, plates (96-well format) are coated with anti-human IgM antibody to capture participant IgM antibody. Virus-specific IgM are detected by the addition of dengue viral antigen, followed by virus-specific hyperimmune ascitic fluid and horseradish peroxidase (HRP)-labeled anti-mouse IgG. Following the addition of colorimetric substrate, plates are read at 410 nm. All acute- and convalescent-phase samples are initially screened at a 1:100 dilution. Samples exceeding the reference cut-off value, calculated as the mean of seven antibody-negative samples plus three standard deviations, are considered IgM antibody-positive. Positive samples will be subsequently re-tested at four-fold serial dilutions to determine end-point antibody titers.

**IgG-ELISA.** Dengue-specific IgG titers will be determined by an IgG ELISA adapted from Ansari et al. (1993). Plates (96-well format) are coated with DENV antigen (cocktail of serotypes 1, 2, 3 & 4) produced from infected Vero cell culture lysates or uninfected cell lysates as controls. Aliquots of diluted participant's serum samples (1:100) are added to two dengue antigen coated wells and to two control wells. After addition of HRP-conjugated mouse anti-human IgG, OD values are recorded at 410 nm. Adjusted OD values are calculated by subtracting the OD of the uninfected antigen coated well from that of the corresponding viral antigen coated well. The cutoff OD value for determining antibody positivity will be calculated as the mean adjusted OD plus 3 standard deviations of antibody negative control sera.

**RT-PCR.** Viral RNA will be prepared from 140 μl of each sera sample using QIAamp Viral RNA Mini Kits following the manufacturer's instructions (Qiagen Inc., Valencia, CA 91355). Nested DENV RT-PCR will be performed following the protocol of Lanciotti et al. (1992) on serum samples for dengue viral RNA detection.

**Virus Isolation.** Isolation of DENV serotypes will be attempted on all acute serum specimens. Using standard virological techniques, cell cultures (C6/36 and Vero cells) are inoculated with serum and monitored for cytopathic effects. All cells inoculated for virus isolation will be tested by immunofluorescence assay using DENV serotype-specific monoclonal antibodies whether or not the cells show cytopathic effects.

**Testing for other Viral Pathogens.** It is estimated that dengue virus is responsible for approximately 30% of the febrile illness observed in Iquitos, Peru. Although the focus of the protocol is dengue, the SR intervention could potentially impact other arthropod-borne diseases. In addition, viral culture inherently detects viruses other than dengue. Thus, blood samples from febrile participants will be screened for a minimum of 6 other arboviruses circulating in and near Iquitos (Yellow Fever virus, Oropouche virus, Venezuelan equine encephalitis virus, Mayaro virus, Group C viruses). Additionally, based on the clinical signs and symptoms, samples will be tested for other pathogens including Leptospirosis, rickettsia, and other viral pathogens (influenza viruses, flaviviruses, alphaviruses, hantaviruses and other bunyaviruses, and arenaviruses). No screening for sexually transmitted pathogens will be done and consent forms will reflect that we may be screening for more than dengue virus. Finally, longitudinal samples may also be screened for evidence of past infection by other viral pathogens, excluding HIV, HTLV, or other sexually transmitted pathogens, if there is evidence of their circulation. Based on our experience with previous cohorts, participants show active interest in knowing the cause of their febrile illness and their past exposure to other non-DENV viral pathogens. Thus, results from all assays will be made available to study participants when feasible.

##### d. Risks to subjects associated with the research: (Range of possible risk of harms: e.g. psychological, social and economic

Risks to study participants are minimal. The only *invasive* procedure used to collect biological specimens from human participants is venipuncture (blood/serological sample). As such the potential risks to participants are:

*Immediate risks*: Occasional bruising and a slight risk of infection at the site of the venipuncture are the most serious injuries that a volunteer can receive from participating in this part of the study.

*Longer term risks:* None are anticipated for this part of the project.

Risks associated with the SR Intervention. There is no unusual risk for households and their occupants participating in this study. Adverse toxicological effects are not likely since we will be following the manufacturer's label instructions (SCJohnson, USA) and trained personnel will provide oversight for SR application. Transfluthrin is registered for public health use in Peru ((1628-2011/DEPA/DIGESA/SA). The active ingredient is approved for human use by WHO (WHO 2006) and used in numerous consumer products throughout the world such as emanators. When pyrethroids are applied in other formulations (Sprays) at doses that are toxic for mosquitoes, itching, pricking sensations, numbness, burning of the skin and the eyes or tingling of the skin are the symptoms most commonly reported after contact with pyrethroids (6 studies cited in WHO 2005). Symptoms usually start 1-6 hours after exposure (3 studies cited in WHO 2005) and last not more than 24 hours, but in some cases they may last up to 3 days. Pyrethroids are not carcinogenic, genotoxic or toxic to reproduction in experimental animals. While data from humans are very limited, it is unlikely that these insecticides pose a carcinogenic or reproductive toxicity hazard to humans (WHO 2005). Overall, when WHO evaluated different uses for public health purposes they state that the onset of parasethesis and upper respiratory tract sensory irritation, especially in asthmatics cannot be ruled out but no long-term or serious health risks are anticipated from pyrethroids at WHO recommended levels (WHO 1998, 2005, 2006, 2012). No special recommendations are need for pregnant woman or children (see Annex 12). *It should be noted that the formulation we are using is a doses significantly lower than for spray formulations noted above and therefore the likelihood of these symptoms are significantly lower.*

Although, label instructions advise not to touch the cards and keep them out of reach of children, consequences of exposure or ingestion of the cards are considered minimal; some people may experience coughing, sneezing, eye irritation and may also have some skin sensitivity to the insecticide. This is likely to be only mild and temporary. Any health related problems would be reported to a physician at a local health center and we will provide contact information so that a NAMRU-6 project physician could visit any participant with symptoms. No psychological, social, or legal consequences are anticipated for this study. Participation in this study will not affect residents' normal access to medical care and treatment through the local health department, or access to vector control administered by the MOH. Pamphlets outlining these risks will be provided to house residents.

For other study procedures such as the entomological surveys, questionnaires/interviews, or surveillance visits the only potential immediate risks are: Participants may find that the entomological visits, questionnaires, and focus group discussions are inconvenient or take up too much time.

Rationale for the necessity of such risks:

In terms of the procedures employed in the project, risk is minimal and with proper training and quality control can be become negligible. Venipuncture will be performed by qualified phlebotomists from NAMRU-6. Universal standard safety precautions including using aseptic technique and single-use sterile needles will be followed when handling blood samples. No significant adverse effects are expected from the phlebotomy. Participation in this study will not affect residents' normal access to medical care and treatment through the local health department, or access to vector control administered by the Loreto Regional Heath Department. Furthermore, no psychological, social, or legal consequences are anticipated for this study. To date, there is no stigma associated with having dengue, and community members are interested and willing to participate in projects that might help improve the health and dengue prevention situation. We will emphasize that individuals are not obligated in any way to participate and can retire from the project at any time without repercussions. The risks to subjects are reasonable in relation to the anticipated benefits to research subjects because the risks associated with venipuncture are negligible.

#### d. Benefits to subjects associated with the research:

Households who participate will potentially benefit from application of mosquito control measures. Repellent applications should supplement the national control program and are likely to result in a decrease of mosquito biting activity and a reduction in DENV transmission.

Subjects with febrile illness will receive appropriate case management from the onset of symptoms, per the national guidelines. Should a study physician determine that the subject is suffering from severe dengue using WHO standard definitions [WHO, 2009] or that, even in the absence of criteria for severe dengue, the subject should be hospitalized, the subject will be referred to the local hospital and treated according to the medical judgment of the local physician.

For those in control areas we will provide the same treatment after the conclusion of the 2 year study. Finally, information from these studies will guide the MOH in their selection of dengue control strategies.

Without the type of data generated from this study, Public Health officials are unable to justify the use of new potentially efficacious products. What is most exciting about this strategy is its superior logistic advantages for both non-epidemic and emergency control. Although maintaining cards in households continuously might be challenging for a MOH, distribution of cards could be carried out rapidly and sustained during an outbreak. This could provide a cost-effective alternative to emergency ULV spray campaigns with longer product duration per deployment (i.e. up to 14 days). In addition, the SR product may also be marketed as a consumer product thereby homeowners will contribute to use and coverage during non-epidemic time periods thereby potentially offering protection to individuals throughout the year.

Risk – Benefit Balance

As stated previously, the risk in this study is minimal and can be reduced further with good clinical and operational practices. Because risk to participating households is minimal, and the knowledge gained from the project could provide more cost-effective *Ae. aegypti* control strategies to the MOH, the benefits clearly outweigh any risks involved in this project. (Annex 11).

### Part VII – Data Analysis and Sample Size Calculation

**Sample Size Calculations:**

A cohort of up to 9,000 participants will be enrolled for febrile surveillance of mild apparent disease. A subset of this cohort ~ 3,400 subjects will also be enrolled from the febrile surveillance cohort for measuring dengue virus infection. A distribution of 2,880 subjects (~1,636 houses) will occur among 24 2-block clusters (12 2-block clusters per study arm - treatment and placebo). This will provide 80% power to detect a minimum of 30% PE at an estimated force of infection (risk) of 10% over 2 years, an anticipated 65% participation rate and $p < 0.10$. These thresholds are based on expert opinions. A design effect factor of k=0.25 was applied to sample size estimates based on assumptions of moderate to high intercluster spatial-temporal heterogeneity in transmission. A 28% attrition rate (loss to follow up; LTFU) will be applied during recruitment to ensure minimum cohort size thresholds are met.

*1. Sample Size Calculations*

**Dengue Seroconversion**

Dengue virus transmission is characterized by considerable spatial and temporal variation so that predicting typical transmission rates for the study areas is problematic. The best strategy to combat this problem is to maximize serological monitoring to the limits of the personnel and financial resources available which are monitoring approximately 1,200 households and 2,400 persons longitudinally (1,200 in both treatment and control areas). Previous studies from Iquitos show that under conditions of endemic transmission (prior to 2002) overall seroconversion rates ranged from 3-4 conversions for every 100 susceptible people per year. The 2002 epidemic began as a combined DENV-1 and DENV-3 epidemic with a significant increase in rates of DENV transmission in January-April 2002 (18.4 conversions/100 person-years at risk). Peak seroconversion rates were observed in September-December 2002 (62.34 conversions/100 p-years at risk). Most recently DENV-4 was introduced into Iquitos in 2008. At the end of 2009 seroprevalence rates for DENV-4 were about 10% and by May 2010 seroprevalence rates to DENV-4 reached 20-30%.

**Annex 9** outlines the process for sample size calculations and the number of clusters, participants and households needed to find a significant reduction in transmission (at 0.90 in a two-sided test) at a given percent reduction (effect size) for an estimated 10% seroconversion rate (risk of infection) with a power of 80%.

**Entomological Indices**

*Aedes aegypti* population densities are highly variable overtime and space. Previous studies in Iquitos showed that the distribution of pupae and adult *Ae. aegypti* were highly aggregated, with the mean (+standard deviation) number of pupae and adults collected per household was 2.99 (+ 22.9) and 0.545 (+4.19), respectively, based on a 3.6 years of collection data. We used a sample size calculation formula for a one-sided, two-sample test for a reduction of counts associated with treatment for organisms showing a negative binomial distribution. The negative binomial distribution is appropriate for data with low average counts but high variably, as observed for these mosquito collections. The dispersion parameter k was estimated by fitting adult *Ae. aegypti* counts to the negative binomial distribution using PROC GENMOD in SAS (SAS 1990); k was determined to be 0.1486 for this data. Table 2 illustrates the sample sizes needed to detect different size reductions in the *Ae. aegypti* entomological indices. Detection of a 50% reduction in adult and pupal indices would require 400-500 households in each group.

**Table 2.** Sample sizes necessary to achieve an expected % reduction, assuming a negative binomial count distribution with dispersion factor, k = 0.1486, a 95% level of significance (□=0.05) and power of 90% (□=0.10). The column highlighted in green reflects historical rates for adult collections; the column in yellow are those for pupal collections.

| % Reduction | Average Counts per household (untreated) | | | | |
|---|---|---|---|---|---|
| | 0.20 | 0.5 | 1.25 | 3.0 | 7.5 |
| 30% | 2020 | 1487 | 1274 | 1191 | 1155 |
| 40% | 1101 | 809 | 692 | 646 | 627 |
| 50% | 684 | 502 | 429 | 401 | 388 |
| 60% | 462 | 339 | 290 | 270 | 262 |
| 70% | 331 | 243 | 208 | 194 | 188 |
| 80% | 248 | 182 | 156 | 146 | 142 |
| 90% | 192 | 142 | 122 | 114 | 111 |

*2. Data Analysis*

Data analyses associated with the longitudinal and febrile surveillance components of the study will identify quantitative relationships between entomological parameters (mosquito population densities and age structure) and dengue incidence (overall, asymptomatic, and symptomatic).
<u>Incidence calculations and case classification.</u> In this section we define (1) predictive and response variables for pattern and regression analyses and (2) the sources of data from the longitudinal study (longitudinal bleeds) and active surveillance. After each longitudinal sample, all participants in the project will be placed into one of the following categories: lost to follow up (LTF), not DENV infected, inapparent infection, symptomatic, or symptomatic with hemorrhagic manifestations. Demographic and entomological household characteristics for people in the LTF group will be compared to individuals who remain in the study. Serologic data on people who are LTF will be excluded from incidence calculations for the sample period prior to leaving the project.

| Case Classification | Description |
|---|---|
| Lost to Follow-up (LTF) | Participants who move from the study area or choose to stop participation in febrile surveillance |
| Not DENV infected | No change in PRNT results between longitudinal bleeds |
| Inapparent Infection | Change in PRNT results consistent with a seroconversion between longitudinal bleeds (i.e. Baseline=Negative, 12 month sample= positive for Dengue 4 antibody) |
| Symptomatic infection | Clinical case indentified in active surveillance with laboratory evidence of dengue virus infections (virus isolation, elevated or seroconversion of IgM ELISA) |

Symptomatic infections identified in the active surveillance study will be used to calculate incidence—that is, laboratory confirmed clinically apparent cases divided by the number of person-days of weekly surveillance. Using a person-time approach we can adjust for households that were closed during visits or when people leave the area for work or vacation and return later. Seroconversion rates will be calculated from those participating in the longitudinal studies, by identifying individuals whose sero-status changes between blood draws. The asymptomatic to symptomatic case rate will be calculated from this group of participants. Illness and seroconversions will be linked to the home residence. We recognize that participants can be infected away from their homes.

Entomological Analyses. With the information generated from entomological surveys and household questionnaires we will calculate the following indices of *Ae. aegypti* density to compare between active and placebo treatment arms: house index (HI, % of houses infested with immature *Ae. aegypti*), container index (CI, % wet containers positive for *Ae. aegypti* larvae or pupae), Breteau index (BI, No. of containers positive for immature *Ae. aegypti*/100 households surveyed), household pupal index (HPI = % houses infested with *Ae. aegypti* pupae), pupae per household (Pu/Hse = No. pupae collected/No. households surveyed), pupae per person (Pu/per = No. pupae collected/No. people living in households surveyed), pupae per hectare (Pu/Ha = No. pupae collected/No. hectares surveyed [sum of lots surveyed]), adult index (AI = % houses infested with adult *Ae. aegypti*), adults per household (AA/Hse = No. adult *Ae. aegypti* collected/No. households surveyed), adults per person (AA/per = No. adult *Ae. aegypti* collected/No. people living in households surveyed), adults per hectare (AA/Ha = No. adult *Ae. aegypti* collected/No. hectares surveyed [sum of lots surveys]), adult female index (AFI = % houses infested with adult female *Ae. aegypti*), adult females per household (AAF/Hse = No. adult *Ae. aegypti* females collected/No. households surveyed), adult females per person (AAF/per = No. adult *Ae. aegypti* females collected/No. people living in households surveyed), and adult females per hectare (AAF/Ha = No. adult *Ae. aegypti* females collected/No. hectares surveyed).

Efficacy Evaluations. Seroconversion rates between treatment and control areas can be compared directly by Chisquare analysis, but more complex multivariate logistic regression models will be developed between predictive variables (mosquito indices, spatial repellent product coverage, treatment area) and response variables (seroconversions, symptomatic disease). Additional analyses with epidemiological data will include comparing cumulative infection incidence (number of events divided by the total person-time at risk) in the cohort with spatial repellent product to the cohort with placebo. Analysis will be carried out at multiple geographic scales starting at the individual household to block to zone level. Protective efficacy (PE) will be measured based on number of dengue infections powered for statistical significance. Analyses will be conducted to compare PE on dengue virus infections between control and treatment arms using the following equation:

*Equation 1:* $PE = = [1-(I_c/I_0)] * 100\%$.

Where, $I_0$= the number of new infections in placebo houses, $I_c$ = the number of new infections in active households (www.OpenEpi.com).

To identify differences in mean abundance of numbers of adult *Ae. aegypti* mosquitoes (broken into age groups = parous versus nulliparous) and pupae, as well as numbers of water-holding, positive, and pupae positive containers we will use negative binomial regression models procedure (PROC GENMOD) of SAS (SAS Institute, 1990). Models will be constructed for the dependent variables (i.e., AD/HA. PU/PER) and treatment area (SR or Control) will be the primary independent variable. Possible confounders such as lot size, human population density, block coverage rates or other household characteristics will be included if potential confounding is observed. Least square means (LSMeans) will be used to test differences among mean rates within main effects and interactions terms; when appropriate main effect variables were then classified into groups and differences were tested using contrast statements in PROC GLM (SAS Institute, 1990).

Role of Human Movement. Because *Ae. aegypti* is a daytime biting mosquito, it is expected that the daytime activity patterns of human hosts will profoundly affect their risk of exposure to infective mosquitoes. People's movement patterns, despite being hard to measure, clearly contribute to the geographic scale at which DENV transmission occurs impacting our ability to evaluate an interventions efficacy, as well as determine an individual's exposure to DENV transmission. Thus, there is an urgent need to quantify time spent in and outside an area where an intervention has been deployed.

Preliminary analysis of Activity Space pilot study data reveals people have a fairly good recall of locations visited, but their recall of time spent in places is poor. Thus, creating a linear variable to describe amount of time spent away from study area is not ideal. However, a categorical variable, representing low, medium and high amounts of time spent away from home will be created. Time spent away from home per week will be summarized into a bivariate variable where "high movers" represent the tercile that spends most time away from home. The "high mover" variable will then be added to logistic regression models estimating the association between living in an intervention household and dengue incidence. A logistic regression analysis will be conducted to examine the association between the treatment households and dengue incidence among individuals in those households, adjusting for the extent of movement of the different individuals. The model can be simplistically represented by:

**Dengue incidence$_i$ = $\beta_0 + \beta_1$(treatment)$_h$+ β2(extent of movement)$_i$ + […] + $\mu_i + \mu_h$**
**(where i=individual level and h=household level)**

**Methods for recording, and coding of data and/or specimens**
Database management will meet or exceed all standards for data accuracy and security required by participating IRBs, Peruvian Instituto National de Salud, and LRHD. Drs. Morrison will supervise onsite collection and initial verification of all entomological data; serology and virology data will be managed at NAMRU-6 by LTC. Hontz. Databases will be shared electronically among key project personnel for analyses, publications, oral presentations, and project development.

The information from these studies is not expected to be sensitive or lead to any negative ramifications such as legal or employment risks, and standard measures to protect confidentiality should be sufficient. Household codes that are georeferenced in our GIS for the city will provide x,y coordinates for spatial statistical analysis, which will be linked to entomological and serological data at these sites. No participant identifiers, beyond the participant's code will be included within our GIS or shown on printed maps or in publications of the study area. Databases generated from these studies will only contain code identifiers and never names. As stated above, it will be clearly explained to participants that information from these studies will be kept strictly confidential.

### Part VIII – Informed Consent

1. **Type of Consent,** ***please explain the reasons for using each and verify that all the elements are present.***

In response to discussions with representatives of the NAMRU-6 IRB, they indicated that for minimal risk studies, alternatives to written consent forms should be sought. We are proposing the following consent format for this study: written consent for blood samples and consent without written documentation for household participation (census information, entomological monitoring and placement of SR intervention).

##### a. Written Informed Consent:

Written consent will be obtained for participation in components of the study that requires a blood sample: longitudinal blood samples at baseline, 12 months and 24 months; and acute and convalescent blood samples when a person has a febrile episode. Although minimal risk, taking blood is invasive and is associated with discomfort and many Iquitos residents have fear of needles. In addition, rumors about the use of blood samples (i.e., pooling for transfusions and receiving payment) are common. For these reasons we believe written consent is warranted and provides important credibility to the project for the specific procedure of taking a blood sample.

##### b. Verbal consent:

We are proposing consent without written documentation for household participation (census information, entomological monitoring and placement of SR Intervention). Our field technicians will continue to provide a rigorous consent process where information sheets, which contain all key elements of a consent document, will be reviewed with participants and verbal consent documented with the letters "CI" placed next to the head of household providing consent on census forms.

The use of information sheets for these less invasive procedures has been particularly useful for improving comprehension of our projects. Because there are many components, this strategy allows us to break the components of the project into smaller, "digestible" bites and contrasts well with blood samples that require written consent. There is distrust overall for signing documents, especially for simple activities that are carried out by local health authorities.

Of particular importance to this protocol is a clear explanation that the SR cards may or may not be treated and that study personnel will not know the status of the cards (treated or untreated). We will explain why it is necessary to have control house houses to test the product.

For all studies it will be emphasized that if at any time the family no longer wants to participate in any of these components they simple can tell the field worker not to visit anymore and say no to the entomological surveys. This is already the case operationally.

**c. Waiver of Informed Consent:**

2. **Will anyone other than the subject be authorized to provide consent or permission for the subject's involvement in the research** *(e.g., parents, court ordered guardian, spouse, etc.)***?** Yes

   If YES,*( please explain)*:

   Parental informed consent will be obtained from children aged 3-17 and both assent to participate and a parentally signed ICD will be obtained for 8-17 year old persons.

3. **Will all adult subjects have the capacity to give informed consent? If NO, describe the likely range of impairment and explain how, and by whom, their capacity to consent will be determined** *(NOTE: In research involving more than minimal risk, capacity to consent should be determined by a psychiatrist, clinical psychologist, or other qualified professional not otherwise involved in the research. Individuals who lack the capacity to consent may participate in research only if a legally authorized representative gives consent on their behalf and if there is prospect of direct benefit to the subject)***:**

   Although, not common, mentally challenged residents often show interest in providing blood samples. In these cases we require assent and parental consent from a responsible relative

4. **Will you obtain consent for the future use of specimens?** Yes
   If YES*, please indicate where the option in the document is*:
   The consent forms for participation in the febrile surveillance and longitudinal cohort will have a separate section for requesting permission to store the participant's blood/serum for future use in studies either on dengue or other arboviruses, in view of the fact that a) it may be necessary to revalidate the original laboratory findings at some point in the future; b) new and improved serological/diagnostic tests may become available whilst the project is in progress which would be beneficial to the project; c) new viruses may be discovered over the course of the project for which it would be advantageous to screen the study

participants retrospectively. The participants will be able to refuse consent for future use of their blood samples, without any repercussions. Should the participants agree to the future use of their specimens, the blood/serological samples will be stored indefinitely at secure laboratory facilities at NAMRU-6 and will not have any participant identifiers, beyond the participant's code.

5. **Please explain the circumstances surrounding obtaining informed consent, with respect to privacy and confidentiality:** *Enrollment plan, details of place, time, and types of advertisements, whether it is group consent or individual and any other information to help your IRB better assess the process*:

Consent will be obtained in participants home. Study personnel will visit prospective households in a door to door fashion, distributing information sheets, and providing an overview of the study. Written consent documents will be obtained 1 week to 1 month after these initial visits at the time blood samples are obtained. Study explanations are usually provided to small groups of adults present in the household, whereas written consent is obtained from each individual participant. In our 15 years of experience conducting these kinds of studies in Iquitos we have found absolute privacy is rarely necessary. If our study staff senses that an individual wants or needs privacy they have been trained to set up a meeting outside the household.

#### Part IX – Confidentiality and Privacy Safeguards for Maintenance of Records, Data Analysis, Sharing Data and Presenting Results

1. **Please describe how/where informed consent documents and study data will be stored, and identify the persons that will have access to them:**

See description of data protection plan in the next section and consent forms will be registered in our database. The information from these studies is not expected to be sensitive or lead to any negative ramifications such as legal or employment risks, and standard measures to protect confidentiality should be sufficient. All paper data forms will be stored in locked files or cabinets in the UC Davis Iquitos Field Laboratory in a specified storage facility with limited access. All data will be stored on secure servers in Iquitos and UC Davis which allow access only to appropriate study personnel. All files sent to consultants, collaborators and co-investigators will have names and address removed. Household codes that are georeferenced in our GIS for the city will provide x,y coordinates for spatial analysis and model parameterization. No household identifiers, beyond the code will be included within our GIS or shown on printed maps or in publications of the study area.

Pupae, larvae, and adult mosquitoes collected during entomological surveys are labeled with unique household codes for transport to our Iquitos field laboratory at 377 Callao Street where specimens will be counted as described in section VI. Immature mosquitoes are either destroyed or added to laboratory colony and data recorded on the entomology survey sheet (described in the previous paragraph). All information will be entered into a database stored on a secure server (described below). Adult mosquitoes are placed in plastic vials labeled by geographic code and date, and then stored at -70 C.

Blood samples will be transported from field sites every 2-3 hours to our field laboratory in Iquitos (address: Clinica Naval, Esquina Avenida La Marina y Avenida Trujillo, Punchana), where the tubes will be stored at 4ºC refrigeration until separation. After separation serum will be stored at -70ºC in labeled Nunc polypropylene vials. Blood samples will be transported for testing on dry ice to NAMRU-6 laboratories in Lima by plane.

2. **Please describe procedures to ensure the privacy of subjects and to maintain the confidentiality of data/information** *(e.g., how/where data will be stored, who will have access to the data/codes, what will happen to the data when the research is complete)***:**

All entomological, serological, and clinical data will be stored on secure data servers and kept strictly confidential. Households are assigned codes unique to our database, which are then used to identify all subsequent data we collect. Outside of our database these codes will not be interpretable, rendering the data effectively unidentifiable without access to our servers. Access to the database will be primarily administered through a custom, web-based interface with restricted access privileges and encrypted data transfer (SSL). Access will be limited to certified project personnel and certified associates, who will be provided unique login and password combinations. Database servers will be protected by multiple layers of security. Physical access will be limited to study personnel, with all servers in secure, locked, limited access locations (our database servers in Iquitos are in locked facilities under 24 hour surveillance and our server in Davis is in a locked, limited access server room on the campus of UC Davis). All database servers are protected by firewalls limiting incoming access to secure ports (SSH, HTTPS). Data backups will be encrypted and kept on another secure, limited access backup server at the UC Davis Center for Vectorborne Diseases (CVEC) and University of Notre Dame with tape backups kept at a secure, off-site location. Our server at Davis will be the host of our public access website. Most data collected by this project will be synchronized among servers in Davis and Iquitos though encrypted transmission of binary logs on a nightly basis.

Database management will meet or exceed all standards for data accuracy and security required by participating IRBs, Peruvian Instituto National de Salud, and LRHD. Drs. Morrison will supervise onsite collection and initial verification of all entomological data; serology and virology data will be managed at NAMRU-6 by Drs. Hontz. Databases will be shared electronically among key project personnel for analyses, publications, oral presentations, and project development.

Reports on aggregated data, especially *Ae. aegypti* indices, will be provided to MOH in a timely manner to aid with ongoing city surveillance and control activities, however this data will not contain any identifiers relating to the individual households or participants involved in the study (i.e. the entomological indices are calculated with reference to a certain area, block, or zone). The results of the study will be analyzed and published in scientific, peer-reviewed journals and may be presented in the form of oral or poster presentations at national or international scientific meetings; however, the data and results will be presented such that participant anonymity and confidentiality will be maintained.

It will be made clear during the consent process that no information can be shared with anyone other than designated study personnel, the paper and computer files will be well protected, and we will ask that interviews be carried out one-on-one to prevent other family members listening in. We will take all necessary measures to ensure confidentiality. It will also be made clear to study personnel that any violation of confidentiality would be a fireable offense.

3. **If data with identifiers will be released, specify the person(s) or agency to which this information will be released:**

Subject confidentiality is held strictly in trust by the consultants, collaborators and co-investigators, the sponsors and their agents. This confidentiality extends to the testing of biological samples i.e. blood samples in addition to other information relating to participating subjects. Data and all other information generated during the course of the study will be held in strict confidence. No information concerning the study or the data will be released to any unauthorized third party without prior written approval of the PI.

We will permit access to all documents and records that may require inspection by the sponsor or its authorized representatives.

**4. Please indicate how long research records will be kept and how they will be discarded.**

All paper data forms will be stored in locked files or cabinets in the UC Davis/NMRCD Iquitos field office or NAMRU-6 Lima in a specified storage facility with limited access. Access to computer data files will be password protected to allow exclusive access to appropriate study personnel. The paper data forms associated with the project e.g. consent forms, questionnaires, census, etc. will be stored indefinitely in accordance with U.S. Navy regulations. Following completion of the project if appropriate storage space is unavailable permission to destroy the documents would need to be obtained from the U.S. Navy.

Should consent be given for future use then serological samples will be stored indefinitely at the laboratories of the United States Naval Medical Research Center Detachment, Lima, Peru. The samples will not have any participant identifiers, beyond the participant's code. If, however, consent for future use is not given, the blood samples will be destroyed immediately following completion of the project.

### Part X – Collaborative relations (local capacity building and/or strengthening, technology transfer)

1. **Describe your collaborative relations with the local MoH collaborators in the conduct of this study and your present or future plans for capacity building, addition of local researchers to your team, technology transfer and others.**

This project is being carried out in collaboration with the Loreto Regional Health Department. We have worked closely with colleagues in DIGESA, Iquitos to develop the study protocol, and inform local health workers about the project and the purpose of the traps. DIGESA personnel participated in focus groups and provided significant input on the potential for new vector products in Iquitos (Paz Soldan et al. 2010). Drs. Achee and Morrison have participated in scientific meetings and workshops. The DIGESA has a strong interest in the use of SR products as emergency and non-emergency control tools and are following results of the project closely.

### Part XI – Provision of results to the study participants

**1. Describe your plans for providing results to study participants, including a table with timelines; Also, provide details on any mechanisms that will be used to make sure that results will be made available to all participants.**

Individual Results. All participant providing a blood sample receives a small written report presenting and explaining their results (Annex 12). These are provided to the participant by the member of our phlebotomist team who is responsible for the active surveillance visits for the participants' household. In general, households under surveillance are well known to our team as we have a minimum contact of 3 visits per week, and many of those visits are significant interactions, most head of households and many members are known by name and have built up significant trust. With the exception of children where results are presented in the presence of parents, results are provided to individual participants. Reactions range widely. For results from febrile samples, interpretation has been rather straightforward and, in these case,s is often presented by a project physician. If a participant is positive for dengue, special emphasis is placed on the warning signs and the importance to go to a health facility first and contact us second. Because we provide daily clinical follow up through defervesensce we can reinforce this concept and ensure proper care. In the case that the person is negative in the acute sample we emphasize the fact that we can't know if they have had dengue until testing a convalescent sample and that clinical follow-up is important which our project fortunately provides. As for longitudinal sample results, we provide a summary interpreting their collective PRNT results: infections they had previous to, and then during, the study (based on their tests). We always also explain the fact that they might be infected with dengue again. This explanation requires training of the phlebotomy team, careful explanation in the field and also one-on-one during our "barridos" (6-12 weeks). We have found that the concept of previous infection can be challenging and requires discussion and our team will often ask the participant to explain their results back to them to ensure they understand. Nurse supervisors and physicians often carry out this exercise separately to reinforce the significance of their results. A key message is that many people with dengue do not have obvious disease and that this is an important objective of the study. Finally, when participants have questions the phlebotomist or team member cannot answer, Dr. Morrison often has a conversation with the participant over the phone or in person. What is critical to understand is our team has developed a relationship and trust with our participants, we have constant contact and conversation, and that receiving results is important to our participants even with though we are quite transparent about the fact that the longitudinal results are of no direct therapeutic value to a participant. Because of inherent difficulties, cross-reactions and assay variability in PRNTs we emphasize this and how important these studies are to understanding the value of these assays overall.

Group Results. The pamphlet submitted in the amendment package would be provided to heads of households and other family members present at the time. In addition the same findings presented in that pamphlet have been presented by the phlebotomy teams. Because this pamphlet has not been approved as of yet, it will be distributed and explained during normal febrile surveillance visits to participating households. Truthfully, participants have been asking all along and the phlebotomy teams have been briefed periodically on the progress of the study and this is discussed with participants, many of whom inquire. We are in constant dialog with study participants, the project is interactive. We provided a results pamphlet during our last barrido and results were well received. This document clearly provides more ownership to participants in the project.

#### Part XII – Medical Monitor *(if applicable)*

1. **Describe Medical Monitor procedures *(if applicable)*:**

   N/A

2. **If no Medical Monitor, please provide a justification:**
   Because this study is minimal risk, a waiver for the medical monitor is requested for this study. All study physicians are licensed to practice medicine in Iquitos. Only study physicians or trained phlebotomists will collect specimens.

#### Part XIII – Funding Information

1. **Funding Sources** *(check all that apply)*:

   ☐ DoD Agency *(name)*:
   ☐ Other Federal Agency (name):

☐ Commercial *(name)*:
☒ Other Foundation *(name)*: Bill and Melinda Gates Foundation
☐ Other *(describe)*:

**2. Grantee** *(name)***:** University of Notre Dame

**a. Grant / Contract No.** *(If available)***:**

**b. Grant / Contract or Project Title:** Spatial Repellent Products for Control of Vector-borne Diseases (Bill and Melinda Gates Foundation)

#### Part XIV – Conflict of Interest Disclosure, *(please also see the investigator assurance form)*

**1. Do members of the investigative team have any conflict, potential conflict or perceived conflict of interests, with research sponsors, or that may otherwise reasonably appear to affect or be affected by the research?** Yes

**If so***, please describe*.
Some key personnel receive a portion of their salary for work on this project. The project requires a corporate partner to develop and produce the SR cards, which are patented by SC Johnson. No one on the investigative team is named on these patents and could eventually benefit from commercialization of the SR cards.

**2. List any steps taken to eliminate, manage or minimize potential for conflict of interest, if any, to harm research subjects or research objectivity**

Recognition and training in good research practices mitigate the issue of personnel receiving salary support. This study is a proof-of-principle of the SR strategy and does not endorse a particular product. Furthermore, the Gates Foundation requires a product development plan that ensures accessibility of the SR product to communities in need. All key research team members must successfully complete the CITI Conflict of Interest module.

#### Part XV – Roles and Responsibilities of the Research Team

Dr. Nicole L. Achee **(Lead Principal Investigator, University of Notre Dame)** will be responsible for contributing to study design and general management of the research, ensure compliance with IRB requirements, reporting to funding agency, and participation in manuscript preparation.

Dr. Neil Lobo (**co-Principal Investigator, University of Notre Dame**) will be responsible for contributing to study design, ensure compliance with IRB requirements, manuscript preparation as well as reporting to funding agency.

Dr. John P. Grieco (**Scientific Advisor, Uniformed Services University of the Health Sciences**) will be responsible for providing support during study design and will participate in manuscript preparation and routine site visits to discuss results.

Dr. Robert Hontz **(NAMRU-6-lead Investigator)** is the Director of the arbovirus and febrile illness section of Virology and Emerging Infections Program of the U.S. Naval Medical Research 6 -Lima (NAMRU-6). They will be responsible for the coordination of all laboratory analyses, including PRNT.

Prof. Thomas Scott and Dr. Amy Morrison have over 15 years experience in researching dengue vectors and epidemiology in Iquitos. They are co-investigators who will act in advisory roles as well as in coordination of Iquitos Field activities.

Dr. Steven Stoddard (**External Statistician**) will participate in sample size estimation during study design and data analyses throughout the study.

Dr. Stalin Vilcarromero (**NAMRU-6 Project Physician and Head of Iquitos Virology and Emerging Infections Department)** supervise clinical activities and surveillance teams.

Helvio Astete **(Supervisor-Project Dengue Field Team)**: Will supervise *Aedes aegypti* field team, supervise SR card replacement, and coordinate with DIGESA collaborators.

Clara de Aguila will serve as collaborator and contact person in the Department of Environmental Health, Loreto Regional Health Department (responsible for Vector Control). Although she will not be responsible for the procedures outlined in the protocol, we will coordinate our activities with her to ensure that we have all Ministry of Health control efforts registered and she is receiving regular reports on *Aedes aegypti* indices in our study areas. As stated in the protocol, our study will not interfere with normal Health Department interventions, but we will coordinate closely with this organization to ensure that participants continue to cooperate with Health Department efforts and they will do their best to support our project.

### Part XVI – References


Achee NL, Bangs MJ, Farlow R, Killeen GF, Lindsay S, Logan JG, Moore SJ, Rowland M, Sweeney K, Torr SJ, Zwiebel LJ and Grieco JP. 2012. Spatial repellents: from discovery and development to evidence-based validation. Malaria Journal. 11(1):164. doi:10.1186/1475-2875-11-164.

Achee, N.L., Sardelis, M.R., Dusfour, I., Chauhan, K.R. and Grieco, J.P. 2009. Characterization of spatial repellent, contact irritant, and toxicant chemical actions of standard vector control compounds. J. Am. Mosq. Control Assoc. 25(2): 156-167.

Ansari MZ, Shope RE, Malik S (1993) Evaluation of vero cell lysate antigen for the ELISA of flaviviruses. J Clin Lab Anal 7: 230-237.

Comach G, Blair PJ, Sierra G, Guzman D, Soler M, et al. (2008) Dengue Virus Infections in a Cohort of Schoolchildren from Maracay, Venezuela: A 2-Year Prospective Study. Vector Borne Zoonotic Dis 9: 87-92.

Forshey BM, Morrison, AC, Cruz C, Rocha C, Vilcarromero S, et al. (2009a) Dengue virus serotype 4, northeastern Peru, 2008. Emerg Infect Dis 15: 1815-1818.

Erlanger, T. E., Keiser, J. Utzinger, j. (2008), Effect of dengue vector control interventions on entomological parameters in developing countries: a systematic review and meta-analysis. Medical and Veterinary Entomology, 22: 203–221. doi: 10.1111/j.1365-2915.2008.00740.x

Getis A, Morrison AC, Gray K, Scott TW (2003) Characteristics of the spatial pattern of the dengue vector, Aedes aegypti, in Iquitos, Peru. Am J Trop Med Hyg 69: 494-505.

Grieco, J.P., Achee, N.L., Chareonviriyaphap, T., Suwonkerd, W., Chauhan, K., Sardelis, M., and Roberts, D.R. 2007. A new classification system for the actions of IRS chemicals traditionally used for malaria control. PlosOne. 2(1): e716.

Grieco, J.P., Achee, N.L., Sardelis, M., Chauhan, K. and Roberts, D.R. 2005. A novel high- throughput screening system to evaluate the behavioral response of adult mosquitoes to chemicals. J. Am. Mosq. Control Assoc. 21(4): 404-411.

Grieco, J.P., Achee, N.L., Andre, R.G. and Roberts, D.R. 2000. A comparison study of house entering and exiting behavior of *Anopheles vestitipennis* (Diptera: Culicidae) using experimental huts sprayed with DDT or deltamethrin in the southern District of Toledo, Belize, C.A. J. Vector Ecology 25 (1): 62-73.

Gubler, D. J. *Aedes aegypti* and *Aedes aegypti*-borne disease control in the 1990's: top down or bottom up: Am. J. Trop. Med. Hyg 1989. 40:571–578.

Gubler DJ (1997) Dengue and dengue hemorrhagic fever: its history and resurgence as a global public health problem.In: Gubler DJ, Kuno, G., editor. Dengue and dengue hemorrhagic fever. London, UK: CAB International. pp. 1-22.

Gubler DJ (2002) Epidemic dengue/dengue hemorrhagic fever as a public health, social and economic problem in the 21st century. Trends Microbiol 10: 100-103.

Hayes CG, Phillips IA, Callahan JD, Griebenow WF, Hyams KC, et al. (1996) The epidemiology of dengue virus infection among urban, jungle, and rural populations in the Amazon region of Peru. Am J Trop Med Hyg 55: 459- 463.

Innis BL, Nisalak A, Nimmannitya S, Kusalerdchariya S, Chongswasdi V, et al. (1989) An enzyme-linked immunosorbent assay to characterize dengue infections where dengue and Japanese encephalitis co-circulate. Am J Trop Med Hyg 40: 418-427.
Kochel TJ, Watts DM, Halstead SB, Hayes CG, Espinoza A, et al. (2002) Effect of dengue-1 antibodies on American dengue-2 viral infection and dengue haemorrhagic fever. Lancet 360: 310-312.

Kawada, H., Maekawa, Y., Tsuda, Y. & Takagi, M. (2004a) Laboratory and field evaluation of spatial repellency with metofluthrin-impregnated paper strip against mosquitoes in Lombok Island, Indonesia. J. Amercian Mosquito Control Association. 20:292-298.

Kawada, H., Maekawa, Y., Tsuda, Y. & Takagi, M. (2004b) Trial of spatial repellency of metofluthrin-impregnated paper strip against Anopheles and Culex in shelters without walls in Lombok, Indonesia. J. American Mosquito Control Association. 20:434-437.

Kawada H, Maekawa Y, Takagi M. 2005. Field trial on the spatial repellency of metofluthrin-impregnated plastic strips for mosquitoes in shelters without walls (beruga) in Lombok, Indonesia. J Vector Ecol,30(2):181-5.

Kuno G (1995) Review of the factors modulating dengue transmission. Epidemiol Rev 17: 321-335.

Lanciotti RS, Calisher CH, Gubler DJ, Chang GJ, Vorndam AV (1992) Rapid detection and typing of dengue viruses from clinical samples by using reverse transcriptase-polymerase chain reaction. J Clin Microbiol 30: 545-551.

Lucas, J.R., Shono, Y., Iwasaki, T., Ishiwatari, T., Spero, N. & Benzon, G. (2007) U.S. Laboratory and field trials of metofluthrin (SUMIONE®) emanators for reducing mosquito biting outdoors. J. American Mosquito Control Association. 23: 47-54.

Monath TP (1994) Dengue: the risk to developed and developing countries. Proc Natl Acad Sci USA 91: 2395-2400.

Morens DM, Halstead SB, Repik PM, Putvatana R, Raybourne N (1985) Simplified plaque reduction neutralization assay for dengue viruses by semimicro methods in BHK-21 cells: comparison of the BHK suspension test with standard plaque reduction neutralization. J Clin Microbiol 22: 250-254.

Morrison AC, Minnick SL, Rocha C, Forshey BM, Stoddard ST, Getis A, Focks DA, Russell KL, Olson JG, Blair PJ, Watts DM, Sihuincha M, Scott TW, Kochel TJ. (2010) Epidemiology of dengue virus in Iquitos, Peru 1999 to 2005: Interepidemic and epidemic patterns of transmission. PLoS Negl Trop Dis 4(5): e670. doi:10.1371/journal.pntd.0000670.

Morrison AC, Zielinski-Gutierrez E, Scott TW, Rosenberg R (2008) Defining challenges and proposing solutions for control of the virus vector *Aedes aegypti*. PLoS Med 5: e68.

Morrison AC, Astete H, Chapilliquen F, Ramirez-Prada C, Diaz G, et al. (2004a) Evaluation of a sampling methodology for rapid assessment of *Aedes aegypti* infestation levels in Iquitos, Peru. J Med Entomol 41: 502-510.

Morrison AC, Gray K, Getis A, Astete H, Sihuincha M, et al. (2004b) Temporal and geographic patterns of *Aedes aegypti* (Diptera: Culicidae) production in Iquitos, Peru. J Med Entomol 41: 1123-1142.

Ogoma SB, Moore SJ, Maia MF. A systematic review of mosquito coils and passive emanators: defining recommendations for spatial repellency testing methodologies.
Parasit Vectors. 2012, 5:287. doi: 10.1186/1756-3305-5-287.

Pates, H.V., Line, J.D., Keto, A.J. & Miller, J.E. (2002) Personal protection against mosquitoes in Dar es Salaam, Tanzania, by using a kerosene oil lamp to vaporize transfluthrin. Medical and Vetinerary Entomology. 16:277-84.

Phillips I, Need J, Escamilla J, Colon E, Sanchez S, et al. (1992) First documented outbreak of dengue in the Peruvian Amazon region. Bull Pan Am Health Organ 26: 201-207.

Reiter P, Gubler DJ (1997) Surveillance and control of urban dengue vectors. pp: 425-462, In: DJ Gubler and G Kuno (eds.), Dengue and Dengue Hemorrhagic fever. CAB International, New York, New York.

Rocha C, Morrison AC, Forshey BM, Blair PJ, Olson JG, et al. (2009) Comparison of two active surveillance programs or the detection of clinical dengue cases in Iquitos, Peru. Am J Trop Med Hyg 80: 656-660.

SAS Institute Inc. 1990. SAS/STAT Guide for Person Computers, SAS Insitute Inc., Cary, NC, USA.

Schneider, J. R., A. C. Morrison, H. Astete, T. W. Scott, and M. L. Wilson. Larval habitat characteristics determine size of adult *Aedes aegypti* (Diptera: Culicidae) in Iquitos, Peru. J. Med. Entomol 2004. 41:634–642.

Vasquez-Prokopec GM, Galvin WA, Kelly R, Kitron U (2009) A new, cost-effective, battery-powered aspirator for adult mosquito collections. J. Med. Entomol. 46(6):1256-1259.

World Health Organization: Guidelines for efficacy testing of spatial repellents. Ref: ISBN 978 92 4 150502 4 and WHO/HTM/NTD/WHOPES/2013.1 http://www.who.int/whopes/resources/en

World Health Organization: Specifications and evaluations for public health pesticides. World Health Organization, Geneva; 2006. www. who.int/WHOPES/quality/Transfluthrin_eval_only_Nov2006.pdf.

World Health Organization and the Special Programme for Research and Training in Tropical Diseases (TDR): Dengue: guidelines for diagnosis, treatment, prevention and control; 2009. www.who.int/rpc/guidelines/9789241547871/en/

### ANNEXES

**Annex 1. Summary of Dengue Incidence Rates from previous NAMRU-6/UC Davis cohort studies.**

**Annex 2. Entomology Collection Forms**

**Annex 3. Longitudinal Cohort Data Forms (Household census, participant movments)**

**Annex 4. Time in House Forms**

**Annex 5. Dengue Case Investigation Forms**

**Annex 6. Transfluthrin Specifications**

**Annex 7. Spatial Repellent Product Profile**

**Annex 8.  Toxicology Report**

**Annex 9.  Power Analysis for Spatial Repellent Randomized Cluster Trial**
