## Supplementary material for "Efficacy of a Spatial Repellent for Control of *Aedes*-Borne Virus Transmission: A Cluster Randomized Trial in Iquitos, Peru": S3. Final IRB Approved Study Protocol

### Human Subject Research Protocol

#### Part I – COVER PAGE

1. HRPP Number: NAMRU6.2014.0021

2. Title of Protocol: Spatial Repellent Products for Control of Dengue Virus transmission in Iquitos, Peru.

3. Sponsor or other support: *(List industry sponsor, government support, etc.)*

   Work Unit Title And Number:
   Grant Title And Number: Spatial Repellent Products for Control of Vector-borne Diseases (Bill and Melinda Gates Foundation)
   Other:

4. Principal Investigator:
   a. Name, degree, affiliation, address, telephone number, email:
   **Nicole Achee**, Ph.D., Eck Institute for Global Health, College of Biological Sciences, South Bend, Indiana, USA 001-202-459-1896,
   b. Human Subject Protection Education (date completed):
   18 May 2017
   c. Curriculum Vitae attached

5. NAMRU-6 Lead Investigator:
   a. Name, degree, affiliation, address, telephone number, email:
   **Amy Morrison, Ph.D., M.S.P.H.,** NAMRU-6, Clinica-Naval, Av. La Marina con C/Trujillo #951, Punchana, Iquitos, Loreto, Peru, phone: 965-604279,
   b. Human Subject Protection Education, certificate : 24 August 2017
   c. Curriculum Vitae, attached

6. Associate Investigators:
   *(For each please provide (1) name, degree, affiliation, address, telephone number and email, (2) institutional affiliation and (3) date human subject protection education was completed)*:
   a. Name, degree, affiliation, address, telephone number, email:
   **John Grieco,** Ph.D., Department of Biological Sciences, Eck Institute for Global Health, University of Notre Dame, 107D Galvin Life Science, Notre Dame, IN, 46556 USA, phone 574.631.7572,
   **Thomas Scott**, Ph.D., Department of Entomology, University of California, Davis, One shields Ave. Davis, CA95616, USA, phone 530.754.4196,
   **Neil Lobo,** Ph.D., Department of Biological Sciences, University of Notre Dame, Notre Dame, IN 46556 USA, phone 574.631.4155,
   **Julia Sonia Ampuero,** MD, MTMH, Ph.D, NAMRU-6, Lima, Peru, 511-6144110,
   **LT Eugenio Abente**, NAMRU-6, Lima, Peru, 511-6144141 ext 210,

   b. Human Subject Protection Education, certificate
   Grieco: 25 May 2017
   Scott: 09 June 2016
   Lobo: 04 Jan 2017
   Ampuero: 05 January 2017

Abente: 12 December 2018

c. Curriculum Vitae attached

7. Additional Key Study Personnel
(e.g., individuals listed in the protocol, research coordinators, etc.)*(For each please provide (1) name, degree, affiliation, address, telephone number and email, (2) institutional affiliation and (3) date human subject protection education was completed)*:

a. Name, degree, affiliation, address, telephone number, email:
**Helvio Astete**, Department of Virology, NAMRU-6 Iquitos, 377 Callao, Iquitos, Peru, 51-65-600102,
**Clara del Aguila**, B.S., Dirección General de Saneamiento Ambiental (DIGESA), Calle Alzamora 410, Iquitos, Peru
**Robert Reiner,** Ph.D., Statistical modeling support (external collaborator), Institute for Health Metrics and Evaluation | University of Washington,
**Troy Alex Perkins,** Ph.D., Statistical and modeling support (collaborator), Department of Biological Sciences and Eck Institute for Global Health, University of Notre Dame, Notre Dame, IN, USA,
**Juan Wong**, M.D., NAMRU-6, Lima, Peru, 511-6144141x202,
**William Elson**, Consultant, University of California, Davis,
**Manuel Maria Geraldes Barba Beja da Costa,** M.S., Database Manager, UC Davis,

b. Human Subject Protection Education, certificate
Astete: 21 Jan 2019
Del Aguila: 29 March 2016
Reiner: 26 March Nov 2019
Perkins: 23 Sept 2017
Wong: 18 Dec 2017
Elson: 10 May 2016
Beja da Costa: 6 December 2018

c. Curriculum Vitae attached

8. Research Study Site(s):
a. Research Study Site(s): Iquitos, Peru
b. Lead Research Institution (name of site with lead or primary IRB responsibility): NAMRU-6
c. Collaborating Institutions (list all institutions):
University of Notre Dame
University of California, Davis
DIRESA/DIGESA Loreto
University of Washington
d. Host Country Approval Agency (ies) (list all MoH department/offices, local DoD component/command, etc. as applicable):
DIRESA/DIGESA
e. Does a written agreement exist to formalize this collaboration?
No: ☐ Yes: ☒ (*If yes, please provide a copy of the agreement with this application)*:
IRB Relying agreements:
Notre Dame/NAMRU-6
UC Davis/NAMRU-6
University of Washington/NAMRU-6

9. Dates of Research *(Proposed start and end dates):* 15 July 2014 to 30 June 2022

10. Medical Monitor:
Name, degree, affiliation, address, telephone number, email:
Not Applicable (minimal risk study)

11. CRADA number:
9297

12. Program Director:
Name, degree, affiliation, address, telephone number, email:

13. COMMANDING OFFICER:
Name, degree, affiliation, address, telephone number, email:
CAPT Guillermo Pimentel, MSC, USN; US Naval Medical Research Unit No. 6 (NAMRU-6), Peru;; phone: 511 614-4111.

### Part II – Record of Changes

**Amendment #1 (July 2015):**
- Additional Key Study Personnel has been added: Robert Reiner, Ph.D., Indiana University School of Public Health, Bloomington, Indiana, and Troy Alex Perkins, Ph.D., from University of Notre Dame, Notre Dame, IN.
-The names of the department head and commanding officer have been updated.
- Delays is receiving the spatial repellant product resulted in changing the timelines for placing the product in the homes.
- The time interval for replacing products in the homes was changed from 3 weeks to 2-3 weeks to more accurately reflect logistical challenges to field teams. ICDs have been changed accordingly.

**Amendment #2 (March 2016):**
Dr. Christopher Mores will replace Dr. Robert Hontz as the principal investigator.
Dr. Hontz will remain as an associate investigator.
ICDs have been changed accordingly (the name of PI has been removed as POC) and Iquitos land phone number has been updated in the ICDs and information sheets.
Administrative changes: name and contact information for head department have been updated: citi program certification dates have been updated where applicable.

**Amendment #3 (June 2016)**
Dr. Steve Stoddard has been removed from the protocol.
Amendment (1) clarify/increase the number of study participants enrolled in the febrile surveillance cohort, (2) adjust the project timelines slightly to accommodate continued delays in the arrival of the SHIELDS product, (3) add emphasis in the protocol and consent documents that if ZIKV or CHIKV are introduced into the study area we will test for these pathogens (Note this is already included but we add some additional emphasis), and (4) update our clinical data form. Consents and info sheets and protocol have been modified accordingly.

Dates for investigators'Human Subject Protection Education certificate have been updated where applicable.

**Amendment #4 (February 2017)**
Amendment 4 includes the following three modifications: (1) modifies case inclusion criteria and sample collection procedures to improve capture and diagnosis of Zika virus currently circulating in Iquitos; (2) adds a simple questionnaire to access how household participants perceive the efficacy of the SHEILD spatial repellent product, and (3) develop and pilot the use of community volunteers to enhance our symptomatic case surveillance (formally febrile surveillance).

**Amendment #5 (May 2017)**
The associate investigator, Stalin Vilcarromero, NAMRU-6 has been removed from the study.
Madison Bailey and William Elson have been added as key personnel
The dates of citi training for some investigators have been updated.
The name and contact information for the CO has been updated.
The point of contact in the consent forms and information sheets have been changed to remove Dr. Vilcarromero.

**Amendment #6 (May 2018)**
End of research has been extend to 31 December 2019
Key research personnel, Graduate Student Madison Bailey (Notre Dame) has been removed from the study.
Molly Barry has been added as key personnel
CITI training information has been updated where applicable.

**Amendment #7 (July 2018)**
Dr. Morrison will replace Dr. Mores as the PI of the research study.
Dr. Mores will remain in the protocol as associate investigator. His affiliation and contact information have been updated.
Dr. Sarah Jenkins, associate investigator has been removed from the study
Affiliation and contact information for Dr. W. Campbell have been updated
Name and contact information for department head have been updated

**Amendment #8 (April 2019)**
Removed Drs. Wesley Campbell, Joel Montgomery, and Christopher Mores; and graduate student Molly Barry from Protocol.
Added Manuel Beja Da Costa and Dr. Eugenio Abente to protocol.
Updated CITI dates where appropriate.
Updated timeline of some protocol activities and extended febrile surveillance visits and febrile case capture for a period up to 3 years, entomological monitoring through December of 2019, and clarified that the spatial repellent evaluation component of the study ended in March 2019. Other NAMRU-6 studies will use the cohort infrastructure to capture acute febrile illness.

### Abbreviations

| | |
|---|---|
| AI | Active Ingredient |
| CPD | critical path of development |
| CHIKV | Chikungunya virus |
| DCZV | Dengue, Chikungunya, Zika, other arboviruses (Aedes-borne arboviruses) |
| DENV | dengue virus |
| DoD | Department of Defense |
| DIRESA | Dirección Regional de Salud de Loreto (Loreto Regional Health Department) |
| ED50 | Effective Dose for 50% repellency |
| ELISA | Enzyme-Linked Immunosorbent Assay |
| GIS | Geographic Information System |
| HIN | Household Identification Number |
| HITSS | High Throughput Screening System |
| HRP | Horseradish peroxidase |
| IgG | Immunoglobulin G |
| IgM | Immunoglobulin M |
| IRB | Institutional Review Board |
| IRS | Indoor Residual Spraying |
| LLIN | Long Lasting Insecticide treated Net |
| LTFU | Loss to Follow up |
| MFIR | Minimum Field Infection Rate |

| | |
|---|---|
| MoH | Ministry of Health |
| OD | Optical Density |
| PCR | Polymerase Chain Reaction |
| PE | Protective Efficacy |
| RCT | Randomized Cluster Trial |
| RDT | Rapid Diagnostic Test |
| RT-PCR | Reverse Transcription Polymerase Chain Reaction |
| SAE / AE | Serious Adverse Events / Adverse Events |
| SFS | Semi Field System |
| SFS | Semi-field Systems |
| SI | Subject Identity |
| SOP | Standard Operating Procedure |
| SR | Spatial Repellent |
| UIC | Unique Identifier Code |
| WHOPES<br>ZIKV | WHO Pesticide Evaluation Scheme<br>Zika virus |

### Part III – Summary of Research in Lay Terms

*(No more than 500 words)*

Dengue viruses are the most medically important arthropod-borne pathogens worldwide, with transmission occurring in most tropical and sub-tropical regions. An estimated 390 million infections occur yearly. Although, there are considerable ongoing efforts to develop a vaccine, vector control remains the only option for reducing DENV transmission and disease burden. The recent emergence of *Aedes*-borne Zika (ZIKV) and Chikungunya viruses (CHIKV) highlight need for novel vector control tools. The goal of this project is to determine the efficacy of a spatial repellent (SR) product (active ingredient transfluthrin) for reducing contact between household residents and vector mosquitos and as a result reduce DENV, ZIKV, and/or other *Aedes*-borne virus transmission. Henceforth we designate the combined risk of Aedes-borne virus transmission by DCZV. Spatial repellency is used here as a general term to refer to a range of insect behaviors caused by airborne chemicals that reduce contact between people and disease vectors. This can include movement away from a chemical stimulus, and interference with host detection (attraction-inhibition) and/or feeding response.

Protection provided by this product will be measured using entomological and virological approaches, comparing entomological indices will be measured through standard household monitoring of *Aedes aegypti* population densities, while DCZV transmission will be measured through door-to-door surveillance for active dengue disease and through serological monitoring for DCZV exposure in a randomized cluster trial. We will establish a cohort of 3,400 persons, primarily children 2-12 years of age and adults who have not been previously infected with DCZV who will provide annual blood samples when they are healthy, whereas in the same clusters we expect to monitor up to 27,500 residents for active dengue disease. The cohort will be monitored for a period of 2 years. The use of spatial repellents has never been tested on a large scale to reduce disease and could change vector control practices worldwide, reducing the amount of chemical insecticides applied and also prevent the development of insecticide resistance. We plan to implement a short questionnaire to determine levels of acceptability and perceived efficacy amongst participating households.

In 2013 and 2015, Chikungunya (CHIKV) and Zika (ZIKV) virus have emerged in the western hemisphere. CHIKV, an alphavirus and ZIKV a flavivirus very closely related to DENV in clinical presentation, are both vectored by *Aedes aegypti*. Both are circulating in Iquitos, Peru, along with many other arboviral diseases that mimic less severe presentations of DENV. All of the research questions we are posing for DENV are equally relevant for CHIKV and ZIKV. In 2016, the first cases of ZIKA were detected in Iquitos and to date we have identified 12 cases through the NAMRU-6 Clinic-based passive surveillance system. Personal communications with the Loreto Regional Health Department indicate that they have identified about 50 ZIKA infections, but testing contacts have revealed a high rate of inapparent infections consistent with other parts of the world. A complicating factor has been the observation that diagnosis in serum or plasma is very time sensitive with only days to effectively isolate the virus, but the virus persists in urine and other body fluids for longer periods of time out to weeks and as long as 6 months in semen. Also, often these patients may be afebrile, but with dengue like rashes. Appropriate evaluation of PE of SR products against Aedes-borne diseases will require effective capture and diagnosis of ZIKA and CHIKV infections in addition to DENV infections, which will be greatly enhanced through the collection and testing of urine, semen and possibly saliva, nasal or conjunctival discharge, or breast milk in addition to blood to aid in ruling out presence of other arboviral diseases circulating when the predominating symptom may be new onset rash without fever. The presence of these other two arboviral diseases and others (e.g. Mayaro, Oropouche viurses) confound efforts to identify DENV cases and often testing for these other agents is needed to move beyond clinically suspected cases of DENV and a presentation caused by another pathogen. Furthermore, sampling recommendations for these viruses are evolving as epidemiologic transmission studies inform our approach to removing these other agents from the differential of rash with or without fever. Identifying the presence of arboviruses beyond DENV will more definitively test the efficacy of SR products against mosquito-borne disease.

This study protocol was developed principally as Randomized Cluster Trial (RCT) to evaluate a Spatial repellent product for the prevention of Aedes-borne Virus disease and transmission. The intervention portion of the trial ended in March 2019, but additional activities are required by the study sponsor including provision of final study results and community meetings to disseminate these results, post-intervention transmission (unfunded by Gates Foundation) and entomological (funded by Gates Foundation) monitoring. These activities will be finalized by the end of December 2019. The febrile surveillance and longitudinal cohort components of this SR project may be sustained through DOD and other institution sponsored studies provided there is funding. Principally the DTRA funded Multi-Echelon Diagnostics study (NAMRU6.2018.0006) will use part of the SR cohort along with another cohort study, Quantifying Heterogeneities for Dengue Transmission (NAMRU6.2014.0028) to capture acute febrile illness cases for this study. Additionally, we anticipate and plan for MIDRP funding for cohort continuation for FY20-22. Funding for this protocol from the Bill and Melinda Gates Foundation by way of UC Davis will end in December 2019, but the febrile surveillance component of this study protocol will be supported by DTRA and the longitudinal cohort component may potentially be supported by MIDRP.

Prospective cohort studies are critical for addressing gaps identified by the DOD for troop readiness. These include effective vaccines against DENV for improved medical readiness (Gap #203617) and for the mitigation of infectious and vector-borne disease threats during unified land operations (Gap #203221) by the prevention of dengue disease in deployed troops. Vaccine development has been hampered by gaps in our understanding of correlates of protection, especially the role of NT abs, as well as the kinetics and role of cross-reactive antibodies during the immunological response which only a cohort study design can provide. Alternatively, novel anti-viral drugs for prophylaxis or treatment can prevent and treat dengue illness directly addressing the previous two needs assessment gaps. Our overall project objective directly support DENV and other ABV vaccine development by perfecting the infrastructure and procedures necessary to carry out regulated clinical trials for DENV vaccine and potential antivirals, and test novel rapid diagnostics and surveillance systems (Gap#203221 and Gap#203223). NAMRU-6 is currently competing for funding or has been approached by collaborators for the following: (1) seeking funding from the Coalition for Epidemic Preparedness Innovations (CEPI) for a clinical trial for a CHIKV vaccine developed at Walter Reed Army Institute of Research assessing safety of the vaccine in recipients with prior alphavirus exposure (VEEV, MAYV), (2) seeking funding from Peer Reviewed Medical Research Program (PRMRP) of the same CHIKV in alphavirus naïve individuals to assess protection against other alphaviruses, (3) established Non-disclosure agreements with Janssen Pharmaceuticals, part of Johnson & Johnson about some promising antiviral compounds with the potential for human trials, and (4) are establishing a CRADA with Merck Sharp & Dohme Corp., a subsidiary of Merck & Co., Inc, to carry out qualitative interviews to refine and assess a patient-reported outcome (PRO) and an observer-reported outcome (ObsRo) instrument for measurement of symptom severity in dengue illness and eventually test the instrument on a wider-scale, using mobile-device technology, for FDA approval. Each of these initiatives can utilize the existing infrastructure with minimal start up time to execute multiple independent studies. Tools for monitoring study compliance, person-time under surveillance, and self-reporting of signs and symptoms associated with disease are critical for product evaluation and have been developed and are currently bein tested under this protocol. Our cohorts have been leveraged by DTRA partners to carry out “di-risking evaluations of novel lateral flow assays for DENV as well as other high-risk pathogens” and test novel cloud-based reporting of results. These studies have the long-term goal of providing diagnostic capabilities directly in the hands of troops at the point-of-need with the ability to transfer results directly to command centers for rapid feedback to the warfighter. New vaccines, prophylaxis, treatments, or diagnostics would prevent or mitigate the impact of DENV or other ABV on medical fitness.

### Part V – Objectives or Hypotheses

The **primary objective** of the study is to quantify the protective efficacy (PE) of a spatial repellent product in reducing the incidence of dengue infection in a large-scale randomized cluster trial (RCT) in the city of Iquitos, Peru. Project objectives will be met through pilot studies carried out between August 2015-October 2015 and a RCT conducted between July 2016 until a minimum of July 2018 and December 2018.

##### 1. Primary objectives:

- **a.** Compare incidence of DCZV infections in SR treated and control household clusters, receiving standard entomological surveillance and control procedures by the local ministry of health. **The null hypothesis is that there is no difference in measured endpoints (dengue seroconversion and clinical attack rates) between intervention and control arms.**
- **b.** Confirming and measuring the entomological correlates of reduced infection. (e.g., a reduction in adult densities, mosquito infection and parity rates).
- **c.** Determine if the indoor: outdoor ratio of adult *Aedes aegypti* collected is lower in intervention compared to control clusters.

##### 2. Secondary objectives:

- **a.** Evaluate the adoption, maintenance, and sustainability of the SR intervention in the trial clusters.
- **b.** Determine the impact of the SR intervention use on population densities of non-target mosquito species.
- **c.** Provide cohort infrastructure to capture acute febrile cases and the identification of participants with DENV and other ABVs for further immunological, patient reported outcome (PRO), diagnostics evaluations, and development of clinical trial tools and instruments.

**Longitudinal cohort (n=3,400):** Follow-up serum samples will be requested from eligible participants after screening at the same time the SR products are installed (July-December 2016, time 0), and ~6 , ~12 and ~24 months later, primarily during the months of May-July. At each annual bleed, we will recruit participants who have stayed in the study areas under febrile surveillance to participate in the longitudinal portion, replacing individuals lost to follow up, to ensure that the study power is maintained. Identification of seroconversions in participants experiencing 3$^{rd}$ or 4$^{th}$ DENV infections can often be difficult requiring that data be excluded. By restricting our population to seronegative or monotypic we maximize the probability of generating interpretable data. Seroconversion rates will be compared between intervention and control clusters. In the case of novel virus invasions (i.e., Zika virus), previous dengue serostatus becomes less relevant, so we will recruit individuals 2-18 years of age and if more participants are necessary open enrollment to adults.

**Clinical disease surveillance (27,500 residents):** All households on the blocks where longitudinal participants are recruited will be invited to participate in a household census and allow health workers to visit their homes 3x/week to ask if any residents have a fever. When an individual is identified with a fever or in the case of suspected ZIKA in the absence of fever, presenting with rash, arthralgia, arthritis or non-purulent conjunctivitis (Individual level in the table below), in a separate consent process they will be invited to provide acute and convalescent (14-21 days later) blood samples and/or urine, conjunctival, nasal, or saliva samples and receive daily medical exams (physical exam, temperature, vital signs, and tourniquet test) for the course of their illness. While we will be focusing on dengue virus transmission for this study, we also plan to take advantage of the labor-intensive door-to-door surveillance study design to address the objectives of a concurrent study of respiratory pathogen transmission in the region (NMRCD.2010.0010). Human infection by a range of pathogens (such as dengue virus or influenza virus) can lead to non-specific disease presentation, such as fever, headache, and malaise. Thus, arbovirus (dengue, zika, chikungunya) and respiratory pathogen (influenza) surveillance can be integrated seamlessly, maximizing personnel and infrastructure and minimizing inconvenience for the participant. To achieve this alternative objective, during the household febrile surveillance consent process we will obtain permission from the head of household to invite residents with febrile illness compatible with respiratory virus infection, identified during 3x/week home visits, to participate in the separate study. Respiratory samples will be collected from participants under a separate consent process. Confirmed dengue cases will also be invited to participate in separate protocols that include feeding mosquitoes on infected people (NAMRU-6.2011.0002, NAMRU6.2014.0028), or evaluation of novel rapid diagnostic tests (NAMRU-6.2014.003). Incidence of DCZV disease and the symptomatic to asymptomatic case ratio will be calculated between the intervention and control clusters. Residents, living on the study blocks may be recruited as points of contact to report illness to enhance disease capture.

| Household Level (Consent without written documentation, Information Sheet – Explanation of Study) | |
|---|---|
| Inclusion Criteria | Exclusion Criteria |
| • Adult head of households agrees to health visits and census<br>• Individuals spend a minimum of 4 hrs per week during the daytime hours or sleep in the house. Temporary residents can be included | • Households where study personnel identify a security risk (i.e., site where drugs are sold, residents are always drunk or hostile)<br>• Sites where no residents spend time during the day (i.e. work 7d a week outside the home) |
| Individual Level – Clinical cases (Written consent) | |
| • Individual who spends a minimum of 4 hours per week within the household. | • <2 years of ageIndividuals who spend have spent less than 4 hours in the household during the week prior to illness. |

| |
|---|
| • ≥ 2 years of ageFever at the time of presentation or report of feverishness within the previous 24 hours or presenting with a rash, arthralgia, arthritis or non-purulent conjunctivitis (suspicion of ZIKA determined by project physician) |

**<u>Entomological Monitoring (Pilot ~100 households; Efficacy trial ~3,380 households)</u>:** Entomological monitoring will be conducted for both the pilot and the RCT. For the *Pilot study* full pupal/demographic surveys will be carried out in households at the beginning and end of the nine-week monitoring period. In addition, adult mosquito collections will be carried out up to 2x per day. For the *RCT* monitoring, surveys will be conducted at regular intervals in each cluster. For post-intervention entomological monitoring will be carried out at 1-3 month intervals until December 2019.

| Household Level (Consent without written documentation, Information Sheets-Explanation of Study, Entomological Surveys) | |
|---|---|
| Inclusion Criteria | Exclusion Criteria |
| • Adult head of households agrees to surveys | • Properties where study personnel identify a security risk (i.e., site where drugs are sold, residents are always drunk or hostile) |

**<u>SR Intervention (Pilot ~100 households per replicate; Efficacy trial ~3,380 households)</u>:** All households will receive one sheet for every 8$m^2$ area (i.e. a household 100 $m^2$ would require 6-8 sheets) that will be hung from the ceiling. Sheets treated with transfluthrin will be placed in the intervention clusters whereas the control clusters will receive untreated placebo cards. Field personnel will be blinded. Sheets will be collected and replaced at 2-3 week intervals. This portion of the study was completed in March 2019.

| Household Level (Consent without written documentation, Information Sheets-Explanation of Study, Transflutrin Pamphlet) | |
|---|---|
| Inclusion Criteria | Exclusion Criteria |
| • Adult head of households agrees to SR deployment and to provide access to team member at 2-3 week intervals to change cards. | • Properties where study personnel identify a security risk (i.e., site where drugs are sold, residents are always drunk or hostile) |

3. Number of subjects to be enrolled/or charts/records to be reviewed/number of biological specimens to be used in this study: Pilot studies: ~100 households
RCT: ~3,380 households (ask for permission for up to 5,000 households); Longitudinal participant screening (up to 9,000) and active surveillance (up to 27,500); Longitudinal participants eligible for follow-up samples, ~3,400 (we ask for permission up to 5,000 individuals since if we identify more seronegative individuals than expected it will only improve the power of the study).

If only biological specimens will be used, please complete the following information:

a) Biological specimens obtained under a NAMRU-6 IRB approved study; if so, please provide the name and number of the protocol. N/A
Specimens to be used have a future use authorization: Yes

b) Biological specimens obtained from clinical care/collaborative efforts; if so, please provide the source. N/A

4. Duration of individual subject participation *(State, not Applicable for chart/record reviews)*:
Longitudinal and febrile surveillance: 2 years, but with an extension to 5 years (the study is currently slated for 2 years, but we put a clause in saying if we need additional data we will ask if we can continue). An amendment would be submitted before an extension took place. For example, we may try new product formats in subsequent years or need to collect data for additional years if study endpoints are marginally significant or

close to significance. As of March 2019, the study has lasted a total 3 months short of 4 years. We will provide information sheets an explanation to participants during the June-December 2019 period asking for an additional 3-years of participation in the febrile surveillance and longitudinal components of the study. We will emphasize continuation is completely optional and only include the health visits starting in 2020.

5. Age range of subjects *(adults and/or children)*: ≥ 2 years of age.

6. Types of populations involved, *please describe (pregnant women, lactating women, neonates, military, dependent employees, individuals with economical, social, physical or cognitive disadvantages, individuals sedated or traumatized, healthy volunteers)*:

This study aims to enroll all households located in the study area. There will be no exclusion criteria related to gender, ethnic or racial category. Based on previous studies by UC Davis/NAMRU-6, we expect that 52% of the study population will be female. We have also found that adult females are more willing to participate than males. In similar previous studies all participants were Peruvian citizens, Spanish speakers, and of mestizo ethnicity (mixed Amerindian and white). The health status of this population will be mostly healthy, however, we expect that pregnant woman, some individuals with physical and mental disabilities, and individuals with other acute and chronic illnesses and active duty military will be living within the study area; all will be residents of Iquitos, Peru. Since all of these groups are at risk of contracting dengue it would be inappropriate to exclude them if they wish to participate.

Studies requiring blood samples have been limited to individuals ≥ 2 years of age. We excluded younger children because of the inherent difficulties obtaining blood from small children. Pregnant women will be present on the study blocks and allowed to participate in all aspects of the study, because they represent a group at risk for dengue. Little information is available on the effects of DENV infection on fetuses and the procedures for our study do not represent additional risk to the mothers. ZIKV infection does represent a risk for fetuses, but our study procedures do not represent additional risk to mothers and might contribute to their protection. Pregnant women will be provided information on Zika and counseled to take as many personal projection precautions as possible. The fact that they could be in a negative cluster will be emphasized and addition protection suggested. Children are a significant risk group and represent the population with the highest number of susceptible individuals, thus their inclusion is essential to understand the overall transmission dynamics of dengue. Although active duty military members might be living on the study block, their superior officers would have no knowledge of their participation. They are also at risk for dengue and exclusion in this context would be inappropriate and discriminatory.

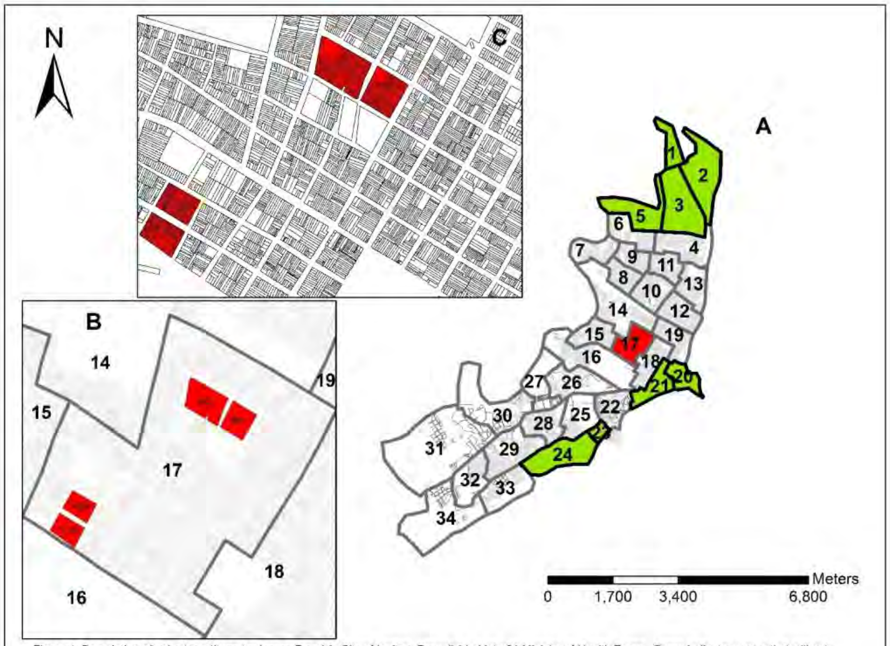

Figure 1. Description of cohort rcrutitment scheme. Panel A: City of Iquitos, Peru divided into 34 Ministry of Health Zones. Green indicates zones that will not not be included, red indicates location of panel B a close up of zone 17, with a single block where recruitment would be initiated shown in panel C.

**Community member points of Contact:** Local residents identified as spending most of their time on the study blocks and with interest in the study may be recruited as study volunteers. They will serve as points of contact for residents to report illness. These individuals will have means to communicate potential cases to study staff via telephone or WhatsApp.

**Time in House:** As part of the initial household census, 1x per month each resident will be asked to estimate the time spent at the residence during daytime hours for weekdays and weekends separately (see Annex 4). This will allow us to control for difference in household exposure to infected mosquito bites among participants.

**Perceived efficacy and acceptability survey:** The Spatial Repellent Questionnaire consists of eight multiple-choice questions to determine participants' perceptions of the spatial repellents' effectiveness. Due to the homogeneity within each cluster, to avoid inadvertently unblinding study staff, the survey will be selfadministered using an Android tablet. A team member will first identify the primary resident in the home, explain the questionnaire and how to use the tablet, and ask if he or she would like to participate. If they receive consent, the study staff will register the participant. Then they will give the tablet to the participant to fill out the survey on their own, sitting nearby in case of any difficulties. If the participant cannot use the tablet or read the questions, someone else in the house, generally of a younger generation, will be invited to read questions to the participant and fill in the participant's answers. If there is no one else home, the study staff will read the questions to them and assist in filling in their answers. Because this occurs so rarely – in a pilot of 93 only 4 times – this will not risk unblinding study staff. More commonly observed in the pilot were the young and old coming together with a shared pride in the younger generation's adeptness with technology (see attached survey). We will carry out additional surveys 2-4 months and about 9 months after SHIELD removal through December 2019.

**Monitoring of DENV Transmission**.

The true impact of the SR vector control intervention will be a reduction in DENV transmission. This will be measured in 2 ways: through serotype-specific seroconversion rates in longitudinal cohort participants providing blood samples at baseline, 12 and 24 months and by identification of active dengue cases through febrile surveillance.

Febrile and DCZV illness capture. This will be implemented at the initiation of the study and will continue for the duration of the study. Public health workers employed by the study will visit each house three times per week. They will record households where no one is home and record any subjects in the house who have febrile illnesses. They will also record changes in census data, noting individuals who move away and individuals who move into the study area. When a participant reports fever or in the case of suspected ZIKA in the absence of fever, presenting with rash, arthralgia, arthritis or non-purulent conjunctivitis, study personnel will take their temperature with a thermometer and administer the informed consent process for DCZV diagnosis. If the subject meets the inclusion criteria described above and provides consent (parental consent and assent for minors) the project physician will be informed, a brief medical exam (vital signs, tourniquet test) and medical history (see Annex 5 -Dengue Case Investigation Form that includes vaccination and dengue history and lists signs and symptoms) will be performed, and an acute phase blood specimen obtained. In the case of suspected Zika, documented by a physician (see "Criteria for a strong clinical and epidemiological suspicion of DENV, ZIKA, CHIKV infection") provide acute and convalescent (14-21 d after acute) blood and/or urine, conjunctival, nasal or saliva samples. If necessary the project physician will evaluate the participant and daily medical exams will be carried out for the course of the illness (usually 3-7 days). If medical exams yield suspicion of DHF/DSS patients will be transported to a local public hospital where their treatment for dengue is free. Ill subjects will be referred to LRHD clinics or hospitals for treatment. Virus isolation will be attempted from acute samples, and both acute and convalescent samples will be tested for anti-dengue and/or other relevant arbovirus antibodies by IgM. In addition, if the participant has a febrile illness with respiratory symptoms such as cough, sore throat, or runny nose, they will be invited to participate in a study of respiratory pathogens circulating in the region (NMRCD.2010.0010). A separate informed consent process will be conducted under that protocol if the participant is willing to provide respiratory samples (nasopharyngeal swabs). Full details of sampling and laboratory procedures are available in the NMRCD.2010.0010 study protocol.

Blood Sample Collection. Blood samples for both the longitudinal and febrile surveillance components of the study will be collected by venipuncture or fingerstick procedures using standard aseptic techniques. An experienced phlebotomist (or study physician) will take the blood sample from an antecubital vein. For the longitudinal measuring of DENV-neutralizing antibodies, up to 5 ml (1 teaspoon) of whole venous blood will be obtained from each volunteer. For febrile participants, up to 10 ml (2.5 teaspoons) of whole venous blood will be obtained. Blood will be collected in one Vacutainer® collection tube (red top) without anticoagulant. If venipuncture is unacceptable to a study subject, the finger prick method will be used for collection of a person's first sample. Providing each participant with a choice of methods will increase compliance and reduce the drop-out rate during subsequent collections. For finger pricks, we will use a BD genie safety lancet and microtainer tube system. After cleaning a finger with 70% alcohol, the sterile lancet will be used to the puncture skin. The finger will be squeezed by the phlebotomist forming a large drop of blood which is held over the microtainer tube. Capillary action draws the blood into the 1 ml tubes. Sera will be separated by centrifugation at 2500 rpm for 10 minutes at $4^0$C, transferred to cryo-vials and stored at -$20^0$C.

**Urine Samples:** Patients who are either febrile or exhibiting rash during enrollment will be asked to provide urine at time of intial blood draw to allow for testing in accordance with current Zika isolation studies. Urine will be collected from both male and female gendered patients following instruction by study staff. CDC guidelines recommend urine testing by rRT-PCR with matched serum. A volume of between 0.5-1mL is required in a sealed sterile screw capped vial. Specimens can be transported on ice (2-8C) or frozen to ≤-20C, and ≤-70C for long term storage (https://www.cdc.gov/zika/laboratories/test-specimens-bodyfluids.html). Collection of urine will follow routine practice of obtaining urine that has ideally been in the bladder for 2-3 hours. Instruction for cleaning of the meatus of the penis or labia of the vagina with a wipe will be provided at time of the collection. Samples will be collected in a sterile cup and immediately placed on ice for transport back to the processing lab.

**Conjunctival /Saliva/Nasal sample collection:** Sampling, if agreed to, will utilize universal transport medium with the soft applicator use to sample conjunctiva secretions, and a separate swab and medium used for swab the buccal mucosa and/or nasal passage. Applicator swabs will be placed in the transport medium and broken off, leaving the swab portion in the medium.

#### SR Description and Deployment (completed in March 2019)

The SR product will be evaluated pending confirmation of minimum performance specifications (**Annex 6,7,8**) against target vector species according to WHO guidelines for efficacy testing of spatial repellents [WHO 2012] to include:

   Perceived efficacy and acceptability survey
   - We plan to carry out a brief, 8 question, survey amongst participating households to determine the perceived effect of SR products on entomological parameters (mosquito density and bite rate) and product acceptability.

- The survey will be carried out 2-3 times during the course of the project to access opinions over time.
- The survey will be carried out 2 times during the post-intervention period (March-December 2019).

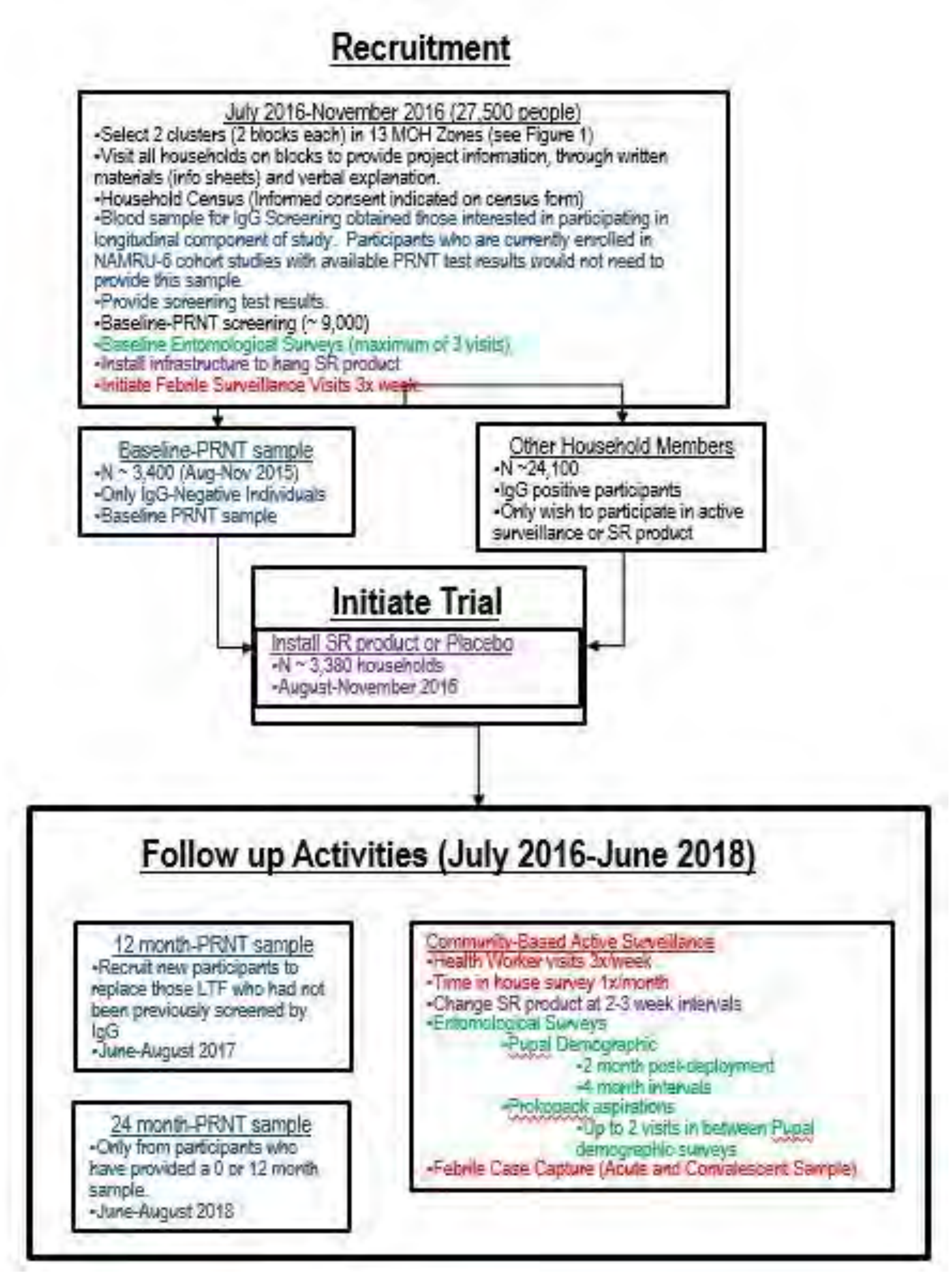

**Figure 2. Flow chart showing RCT activities.**

2. *RCT.* The trial will be carried out from July 2016 to December 2018, but pre-intervention activities could begin as early as July 2015. Data analysis is expected to take until December 2019.
   Pre-intervention assessments/activities
   - House visits, community meetings if necessary, and distribution of study information.
   - Pupal/Demographic Survey and adult mosquito monitoring (2 month intervals).
   - Carry out Household Census
   - Informed consent procedures (documentation for surveillance, entomological monitoring, and SR deployment; written consent for longitudinal blood samples)
   - Time in house surveys
   - Assign unique household codes from GIS maps of the study area
   - Randomize clusters (i.e., block(s) of houses within a single zone) to receive either active or placebo treatment using random number generator.
   - IgG Screening Blood sample for those interested in participating in the longitudinal cohort.
   - Determine SR placement

   Baseline assessments/procedures
   - Informed consent procedures (documentation for surveillance, entomological monitoring, and SR deployment; written consent for longitudinal blood samples)
   - Baseline PRNT blood sample.
   - Pupal/demographic entomological survey.
   - Deployment of SR intervention cards with baseline entomology survey.

   Ongoing assessments/procedures
   - 3 visits from project health worker per week to ask adult family members if anyone in the household has a fever or had a fever within the previous 5 days.
   - SR cards will be removed and replaced at 2-3 week intervals by study personnel.
   - Time in house surveys 1x per month
   - Follow up blood samples ~9 and 24 months post-baseline sample for longitudinal participants.
   - Pupal demographic surveys carried out at 2 month post deployment then at 4 month intervals for the remainder of the project (24 months).
   - Adult mosquito collections using Prokopack aspirator in a sample of study households at 1 week intervals (as part of pupal demographic surveys when performed).
   - Febrile illness capture and monitoring.

   Post-intervention assessments/procedures (all 26 SR clusters)
   - 3 visits from project health workers per week to ask adult family members if anyone in the household has a fever or had a fever within the previous 5 days (after December 2019 study blocks with low participation and high crime will be thanked for their participation).
   - Annual longitudinal blood draws if MIDRP funding permits (This will be done in areas participating after 2020)
   - Pupal demographic surveys carried out at 2-3x between March-December 2019.
   - Adult mosquito collections using Prokopack aspirator in a sample of study households at 1-3 month intervals between March-December 2019.
   - Febrile illness capture and monitoring will continue on all 26 clusters until December 2019, then reduced to areas with high participation and low crime starting in 2020. All people living in study blocks that will not continue will be thanked for their participation after receiving SR study results.

Urine, saliva, nasal, and conjunctival sample processing: RNA will be extracted using Qiagen kits and process as described for blood samples. All fluids will be stored at -80°C.

**Serotype-specific Microneutralization enzyme immunoassay (MNT).** Since 2015, to determine history of DENV infection and identify seroconversions, we will test serum by MNT using a validated NAMRU-6 protocol adapted from Vorndam and Beltran [75]. 96 well plates (TC-treated) will be coated with Vero cells at 2 x 105 cells/ml and then incubated at 37ºC, with 5% $CO_2$ for two days or until the monolayer stabilizes. Diluted virus (dilution factor determined by NAMRU-6 validation assays) with inactivated sera will be incubated at 4ºC overnight. Serum samples will be diluted 1:20 and inactivated at 56°C for 30 min and then added to triplicate wells and serially diluted (2-fold up to 1:1,280).

The serum-virus mixture will be added to Vero cells and incubated at 37ºC with $CO_2$ (5%) for 5 days. After 5 days, the cell culture supernatant will be discarded and the cells will be fixed with ethanol/methanol, washed with PBS, blocked with skim milk, added with HMAF Anti-DENV and incubated for 2 h at 37ºC; washed with PBS again and then incubated at room temperature for an hour with the ABTS substrate. Plates are read using an ELISA reader at 405nm. A cut off value is established for each plate as the numeric value of the 50% of the mean OD of virus controls (WD) and the endpoint titer is the highest serum dilution with mean OD, below the cut off value and is reported as the dilution in that point. Endpoint titers are reported as <1/40, 1/40, 1/80, … ≥1/2560. One of the primary challenges for vaccine development if distinguishing homotypic protective dengue antibody from heterotypic nonprotective but disease-modifying antibody [76]. Furthermore, validation of neutralizing antibody as a biomarker of protection following dengue vaccination will be critical for future efficacy trials.

$$\textbf{Dengue incidence}_i = \beta_0 + \beta_1(\textbf{treatment})_h + \beta 2(\textbf{extent of movement})_i + [\ldots] + \mu_i + \mu_h$$

**(where i=individual level and h=household level)**

#### Methods for recording, and coding of data and/or specimens

#### b. Verbal consent:

We are proposing consent without written documentation for household participation (census information, entomological monitoring, placement of SR Intervention and perceived efficacy and acceptability survey.). Our field technicians will continue to provide a rigorous consent process where information sheets, which contain all key elements of a consent document, will be reviewed with participants and verbal consent documented with the letters "CI" placed next to the head of household providing consent on census forms.

Perceived efficacy and acceptability surveys will be carried out on password protected android mobile devices. The data collected will not contain any personally identifiable information and will include a participant code not interpretable outside the secure database.

Dr. Amy Morrison **(NAMRU-6 Lead Investigator),** will be responsible for the implementation of the study, coordination of all laboratory samples, including PRNT and for all field activities.

Prof. Thomas Scott over 15 years of experience in researching dengue vectors and epidemiology in Iquitos will act in advisory roles.

***Helvio Astete*** **(Supervisor-Project Dengue Field Team)**: Will supervise *Aedes aegypti* field team, supervise SR card replacement, and coordinate with DIGESA collaborators.

***Clara de Aguila*** will serve as collaborator and contact person in the Department of Environmental Health, Loreto Regional Health Department (responsible for Vector Control). Although she will not be responsible for the procedures outlined in the protocol, we will coordinate our activities with her to ensure that we have all Ministry of Health control efforts registered and she is receiving regular reports on *Aedes aegypti* indices in our study areas. As stated in the protocol, our study will not interfere with normal Health Department interventions, but we will coordinate closely with this organization to ensure that participants continue to cooperate with Health Department efforts and they will do their best to support our project.

***Dr. Robert Reiner (External Statistician* and Modeler)** will participate in data analyses, in particular modeling the role of human movement on interpretation of RCT results throughout the study. Will not have access to Personal Identifiable Data.

***Dr. T. Alex Perkins*** (**Statistician and Modeler)** will participate in data analyses, in particular modeling the role of human movement on interpretation of RCT results throughout the study.

***LCDR Wesley Campbell,*** MD, MPH, associate investigator. He will support data analysis and will contribute to manuscript preparation.

***Joel Montgomery***, MS, Ph.D., associate investigator. He will participate in field work, will support data analysis and will contribute to manuscript preparation.

***Julia S. Ampuero,*** MD., MTMH, Ph.D., associate investigator. She will participate in field work, will support data analysis and will contribute to manuscript preparation.

***Juan Wong,*** MD., key personnel. He will aid the project, by assisting in project logistics from Lima, primary the importation permit application with DIGESA.

***William Elson****,* MD, key personnel. He will assist Dr. Morrison with project coordination, in particular management of database and comm care tablet application.

***Manuel Beja Da Costa,*** M.S., key personnel, Will serve as the local Iquitos Database manager, will ensure that relational database is functioning, implement quality controls in system, use system to generate operational lists and tools, and aid investigators with data extraction and cleaning.

***Eugenio Abente,*** Ph.D., associate investigator, He will supervise local Iquitos field staff, support project logistics, facilitate laboratory testing of project blood samples, data analysis, and contribute to manuscript preparation.
