## Supplementary material for "Efficacy of a Spatial Repellent for Control of *Aedes*-Borne Virus Transmission: A Cluster Randomized Trial in Iquitos, Peru": S4. Statistical Analyses Plan

### Spatial Repellent Products for Control of Vector Borne Diseases

**Peru**

Draft Statistical Analysis Plan

**Version 3.0**

**October 26, 2020**

**Prepared by Bobby Reiner**

****

#### Summary of changes to the Peru SAP

| Version | Date | Summary of Changes |
| --- | --- | --- |
| Ver 1.0 | Apr 1, 2019 | Submitted to VCAG for May 2019 Meeting |
| Ver 2.0 | 2020 | Updates based on internal feedback |
| Ver 3.0 | Oct 26, 2020 | **Altering enrollment from exclusively children to any individual who is monotypic or seronegative**<br>Due to enrollment issues, there was a general concern that sample sizes would not be met with only monotypic or seronegative children. The primary requirement of a qualifying participant was that they would have a past exposure background that would allow for the detection of an arbovirus infection. It is extremely difficult to identify tertiary or quaternary dengue infections with the protocol of this study, so this restricted the primary participants to those who were monotypic or seronegative. This primary requirement was not inherently based on the age of the individual, but the age of the individuals was originally specified under the assumption that younger individuals would be less likely to have had multiple dengue infections. Given every individual in the study area was given a MNT test, including adults that were monotypic or seronegative did not include considerable wasted effort.<br><br>**Altering the primary analysis method of the primary endpoint**<br>The original enrollment protocol supposed that individuals would be enrolled at the beginning of the trial, or around the 1-year mark of the trial. In actuality, individuals were continuously enrolled (and several dropped out of the trial within a trial year). As such, the original planned analysis (which assumed that durations of participations were relatively fixed) was inadequate. While the original analysis was still conducted and had a similar result, a more appropriate analysis was conducted. Specifically, we ran a survival analysis with a frailty component to capture variation between clusters. The primary benefit of this approach was that the duration of every individual's participation could be explicitly incorporated in the analysis. **The alteration was discussed with the DSMB external statistician after the dataset was locked and prior to releasing outputs of the original planned analysis to the DSMB external statistician**<br><br>**Altering the secondary analysis of the primary endpoint**<br>The secondary analysis of the primary endpoint is an "intent to treat" analysis. For the same reasons as described above, the method was updated to a survival analysis with a frailty component to account for variation by cluster. The exact definition of "intent to treat" was not defined in the original SAP. Herein we update this oversite to declare an individual received the treatment if there was a SR product in their home for at least 80% of the days they were enrolled in the trial.<br><br>**Updating the primary analysis method of entomological secondary endpoints**<br>We have added more detail to the primary analysis method used for the entomological secondary endpoints. Specifically, we clarify that the mixed effect regressions will be difference in difference mixed effect regressions. |

### 1 Objectives

##### Primary Objective

To evaluate the protective efficacy (PE) of a spatial repellent (SR) against seroconversion to Aedes-borne virus (ABV) infection as measured by micro-neutralization test (MNT).

##### Secondary Objectives

1. To evaluate the protective efficacy (PE) of SR against arbovirus disease as detected by PCR or IgM.
2. To evaluate the effect of SR on female *Aedes aegypti* abundance.
3. To evaluate the effect of SR on female *Ae. aegypti* population age structure, using parity rate as an indicator.
4. To evaluate the effect of SR on *Ae. aegypti* human biting behavior as measured by rates of blood engorged females and female indoor/outdoor abundance ratios.

##### Tertiary Objectives

1. To evaluate the safety of the SR product in human subjects.
2. Evaluate potential diversionary effects of the SR intervention to surrounding homes.
3. To evaluate the resident perceptions on the efficacy and appearance of the SR product using a self-administered mobile application questionnaire.

### 2 Hypotheses

##### Primary Hypothesis

$H_0$: SR does not reduce the probability of individuals seroconverting to ABV compared to placebo.
$H_1$: SR reduces the probability of individuals seroconverting to ABV compared to placebo [seroconversion odds ratio (OR) between SR and placebo is <1; expected odds ratio is 70% or PE is 30%].

##### Secondary Hypothesis

Estimation:

1. The rate ratio of SR versus placebo on ABV disease will be estimated.
2. The change from baseline to post deployment in average household *Ae. aegypti* abundance, parity rate, blood engorged rate, and indoor/outdoor abundance ratio in SR compared to placebo will be quantified.

### 3 Endpoints

- The primary endpoint is the fraction of monotypic or seronegative individuals who seroconvert to an arbovirus during the follow-up period post randomization with intervention. Here, the intervention follow-up period is 2 years after initial deployment of SR or placebo.
- The second endpoints include:
    - Arbovirus disease rate during intervention follow-up period.
    - Entomological endpoints including female *Ae. aegypti* abundance, parity rate, blood engorged rate, and the ratio of the number of female *Ae. aegypti* mosquitoes collected inside of houses versus outside of houses.
- The tertiary endpoints include:

- Safety measures (the frequency of adverse events/AEs and serious adverse events/SAEs) during baseline and intervention follow-up periods.
- Comparing ABV infection and disease metrics as well as entomological endpoints between participating individuals / households in SR clusters and individuals / households from the same clusters who did not agree to the SR component of the trial.
- Resident perceptions include: perceived mosquito density and biting, problems associated with SR product (includes AE above), and willingness to use or buy SR product.

### 4 Study Design

The study design is a cluster-randomized, double-blind, placebo-controlled clinical trial with 13 clusters per intervention arm (SR and placebo). Sixty individuals who are monotypic or seronegative to arboviruses are recruited within each cluster. Individuals are tested for arbovirus seroconversion using MNTs at the end of each transmission season across both years of study of the follow-up period.

### 5 Population for analysis

The intention to treat (ITT) analysis is the primary analysis approach for both the primary and secondary objectives. The ITT population includes the monotypic or seronegative individuals within each recruited household that received at least one SR product or placebo product per the cluster randomization schedule.

The per-protocol (PP) analysis is included as a supplementary analysis for the primary and secondary objectives. The PP population includes the subjects from the ITT population that are treated following the specifications of the study protocol without major protocol deviations. A second PP-like supplementary analysis for the primary and secondary objectives will attempt to estimate fractional impacts of SR for individuals who only received SR products for a fraction of the follow-up period.

#### 5.1 *Subjects who moved to a new house during the intervention follow-up period.*

- For a subject who moved to a different house within the same cluster, that subject will be included in both the ITT and PP analyses.
- For a subject who moved to a different house in a different cluster, the subject will be included in the ITT analysis with the original treatment assignment though the new cluster although the subject moved to might have a different intervention from the original assignment. The subject will also be included in the PP analysis if the new cluster had the same intervention as the original assignment.

#### 5.2 *Subjects who were hospitalized for serious complicated illness (e.g. chronic illness), died, dropped out, or missed scheduled visits due to reasons not related to the ABV disease outcome or intervention during the follow-up period.*

For subjects that fall under this category, the available data from the subjects (up to the time point when the subjects were hospitalized, died, or dropped out; data from the scheduled visits that the subjects did not miss) will be included in both the ITT and PP analyses because the missing or absent data can be ignored (see Section 6.4 of the SAP for more details).

#### 5.3 *Subjects who did not receive (complete) intervention due to travelling outside, mis-application or partial application of the product.*

For the ITT analyses, these subjects will be included as is. For the PP analysis, these individuals will be dropped because they were not treated following the specifications of the study protocol. For the second, PP-like analysis, "travel outside" (Y or N; an individual-level covariate) and the product application rate in each household (expected to be close to 100%) will be included as covariates if the data are not overly imbalanced between the Y and N categories for "travel outside", and there is practically/clinically meaningful variation in the product application rate across households and clusters. An attempt to integrate the seasonality of arbovirus transmission with the period of time that these individuals did or did not receive the product application will be made as possible.

##### 5.4 Replacement subjects

Replacement subjects are defined as subjects who were recruited into the study at a time point after the intervention began to replace initially recruited loss to follow up (LTFU) subjects to maintain minimum cohort numbers. Per this definition, subjects who were absent for an extensive period of time (> 3 scheduled visits), and then returned to study to the same household as before, or to a different household in the same or a different cluster are not replacement subjects (see Section 5.1). As detailed tracking of individual's movements will be conducted, these individuals will be included in secondary analyses.

If the replacement occurs in the baseline period or before the first scheduled visit of the subjects who they replace in either year of the follow-up period, then the data from the replacement subjects will be included in the primary analysis for that year / years. Data from replacement subjects will not be included in the primary analysis for PE if the replacement of the original subject (from the same cluster) occurred after the first scheduled visit of the original subject for that year. However, a supplementary analysis will be performed that includes the replacement subjects.

#### 6 Statistical Methods

##### 6.1 Primary endpoint (ITT Population)

The primary hypothesis on PE against ABV seroconversion will be tested using a survivial analysis with a proportional hazard model with an exponential distribution assumption for the baseline hazard. In particular, if $h(t_{ij}|\boldsymbol{x}_{ij})$ is the hazard rate of the $j^{th}$ individual in the $i^{th}$ cluster with covariate values of $\boldsymbol{x}_{ij}$, then this individuals hazard rate of an arbovirus infection can be writted as:

$$h(t_{ij}|x_{ij}) = h_0(t_{ij}) \cdot \exp(\boldsymbol{\beta}^T \boldsymbol{x}_{ij} + W_i)$$

where $W_i \sim N(0, \sigma_c^2)$ is the random effect of the $i^{th}$ cluster. Covariates included are age, sex, and treatment status (SR or placebo).

If the data are extremely unbalanced in a categorical covariate (e.g., 99% households had the same type of walls) or if a non-ignorable portion of the subjects have missing values on a covariate (due to MAR or MCAR), that covariate may be excluded in the model.

The primary efficacy (PE) will be estimated as $PE = (1 - \exp\hat{\beta}) \times 100\%$, where $\hat{\beta}$ is the estimated regression coefficient for the intervention group and $\exp\hat{\beta}$ is the estimated hazard ratio between SR and placebo. The null hypothesis of $PE = 0\%$ is equivalent to $\beta = 0$, which is tested by Wald's test, $z = \hat{\beta}/s$ where $s$ is the estimated standard error of $\hat{\beta}$, at the 1-sided significance level of 5%.

#### 6.2 Secondary endpoints (ITT Population)

PE of SR protection again incidence of arbovirus disease

The second endpoint on PE of SR protection against the incidence of arbovirus disease will be estimated by conducting a second survival analysis on individuals who received a SR product for at least 80% of the duration of their enrollment in the study.

Entomological effects of SR (on female *Ae. aegypti*)

Entomological effects will be tested using the appropriate corresponding mixed effect regression with random effects by cluster and house. For each indicator, we will use a difference in difference model, comparing the changes in each value from baseline to those measured during the trial between the treatment and control areas.

We expect substantial heterogeneity in all entomological endpoints, and as such expect to find extremely wide uncertainty intervals for estimated effects. To account for this heterogeneity in space and time, we will conduct a secondary analysis using a spatio-temporal model [4]. The model's base structure is still either a negative binomial regression or a logistic regression, but it uses spatial and temporal splines to capture natural underlying variation in mosquito population dynamics.

#### 6.3 Supplementary analysis

The primary and secondary analyses laid out in Sections 6.1 and 6.2 will also be carried out in the PP population, with some modification on the covariate list in the corresponding models for the seroconversion, incidence of disease episodes, and entomological endpoints, as stated in Sec 5 of the SAP. For the second PP-like analysis, "travel outside" (Y or N; an individual-level covariate) and the product application rate in each household (expected to be close to 100%) will be included as covariates if the data are balanced between the Y and N categories for "travel outside" and there is practically/clinically meaningful variation in the product application rate across households and clusters.

As possible, individuals within SR clusters who either do not consent to the SR component of the trial or who enter or leave the trial during the follow-up period may provide an opportunity to assess possible diversionary effects of the SR intervention. Individuals within SR clusters who do not receive the SR product may still consent to the entomological collections, the active febrile surveillance, or, as applicable, the yearly blood draws for ABV seroconversion. A *priori*, there is no guarantee that a large fraction of individuals in SR clusters will agree to participate in the secondary data collection but not the actual SR intervention, and thus it is unclear if there will be power to detect any evidence of diversionary effects (or the lack thereof). That being said, comparisons similar to those described above on both ABV endpoints and entomological endpoints between those who agree to the SR intervention and their neighbors who do not agree will be conducted.

AEs and SAEs will be tabulated and documented.

##### 6.4 Handling of missing data

Per protocol, the subjects are checked for their ABV serostatus (the assay outcome) yearly.

- If a subject missed one or more scheduled visits, the subject will have missing values on the outcome that can be regarded as ignorable missingness.
- If a subject drops out study due to reasons unrelated to the SR product and/or ABV infection, then the missing observations from the subject can be regarded as ignorable missingness.
- In both cases, all available data from the subject will be included in the primary and secondary analysis, without employing any specific technique to deal with the data.

If a non-ignorable portion of the subjects have missing values on a covariate (due to missing at random or missing completely at random), that covariate maybe may be excluded in the model.

##### 6.5 Interim analysis

No formal interim analysis will be performed in this study.

#### 7 Software

Software used will be R version 3.5.3 or higher (R Foundation for Statistical Computing, Vienna, Austria).

#### 8 Sample Size Determination

The sample size determination on the required number of households per cluster is based on the risk of seroconversion comparison in the logistic regression model. Assuming the probability of seroconversion for seronegative or monotypic individuals was 10% with a coefficient of variation of 0.25, and an alpha of 5%, we estimated we would need 26 clusters (13 per arm) with approximately 60 qualifying individuals to achieve 80% to detect a reduction in the odds of 30%. Here, qualifying means a child within a participating house who is seronegative or monotypic.

#### Appendix

I. Mock Tables and Figures

Figure 1: flow diagram of progress of clusters and individuals (From Campbell (2010): *Consort 2010 statement: extension to cluster randomized trials*)

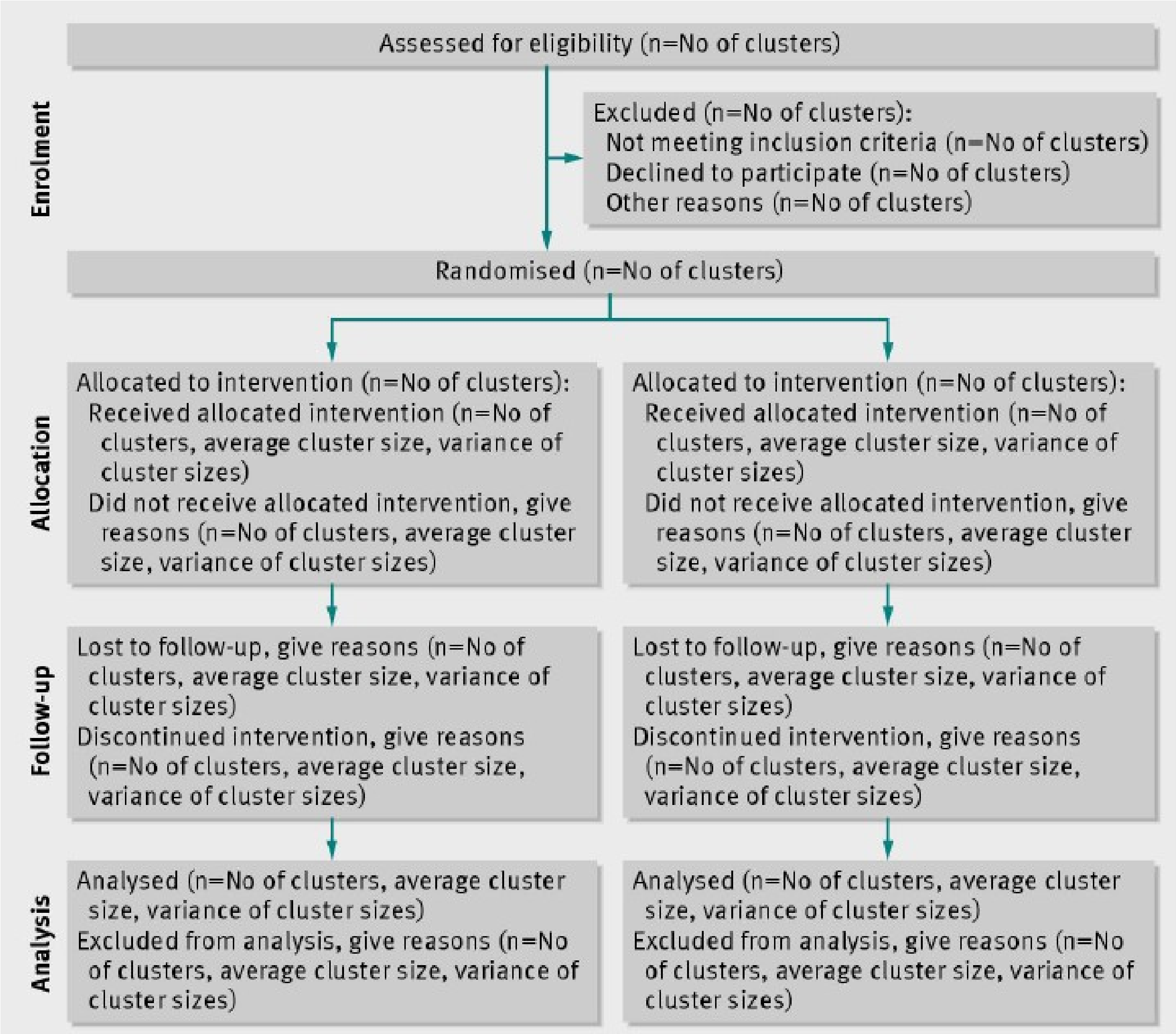

Table 1: Summary on baseline covariates

| | SR | Placebo |
|---|---|---|
| Individual level | | |
| age (mean ± SD, n)<br>gender (% of boys, n) | | |
| household level | | |
| house wall type (%, n)<br>house roof type (%, n)<br>open eaves (Yes%, n)<br># of doors (mean± SD, n)<br>Baseline average *Ae. aegypti* abundance (mean± SD) | | |
| Cluster level | | |
| Cluster size (mean± SD, n)<br>Baseline seropositive rate (mean± SD, n) | | |

Table 2: Protective Efficacy (PE) of SR against seroconversion

| Treatment | Baseline seroconversion rate | # of individuals | # of seroconversions | OR (95% CI) | PE (95% CI) |
|---|---|---|---|---|---|
| SR | | | | | |
| placebo | | | | | |
| Baseline coefficient of variation (CV) of incidence ate: xxx%<br>A similar table will be provided for disease episodes | | | | | |

Table 4: Effects of SR compared to blank on abundance, parity rate, blood feeding rate, indoor/outdoor ratio

| | Mean (95% CI) | | Ratio (95% CI) |
|---|---|---|---|
| Endpoint | SR | Placebo | SR vs. Placebo |
| Abundance | | | |
| Parity rate | | | |
| Blood-feeding rate | | | |

II. Some sample R and R procedures used in the analysis

Note the final codes for estimation of PE could differ slightly from the sample codes below, which are meant to demonstrate the main procedures/commands in R to run the those two types of analyses rather than to be followed strictly.

**a)** For estimating the PE of SR against arbovirus seroconversion using logistic mixed regression

```
require(lme4)
mod_glm <- glmer(outcome ~ age + SR+ as.factor(sex) +
(1|cluster), data= data, family = binomial)
```

**b)** For estimating the PE of SR against arbovirus seroconversion using GEE

```
require(geepack)
mod_gee <- geeglm(outcome ~ age + SR+
as.factor(sex),id=cluster, data= data, family = binomial)
```
