## Supplementary material for "Efficacy of a Spatial Repellent for Control of *Aedes*-Borne Virus Transmission: A Cluster Randomized Trial in Iquitos, Peru": S5. CONSORT Checklist

### Supplemental Information 3. CONSORT 2010 Checklist_Morrisonetal_18Feb2021

| Section/Topic | Item No | Checklist item | Reported on page No |
|---|---|---|---|
| **Title and abstract** | | | |
| | 1a | Identification as a randomised trial in the title | Title Pg1; Abstract Pg 3 |
| | 1b | Structured summary of trial design, methods, results, and conclusions (for specific guidance see CONSORT for abstracts) | Abstract Pg 3-4 |
| **Introduction** | | | |
| Background and objectives | 2a | Scientific background and explanation of rationale | Intro Pg 5-6 |
| | 2b | Specific objectives or hypotheses | Intro Pg 6; SI; Sections 1.2, 1.5; Study protocol (SP); Statistical Analyses Plan (SAP) |
| **Methods** | | | |
| Trial design | 3a | Description of trial design (such as parallel, factorial) including allocation ratio | Methods Pg 7; SI; Sections 1.2, 1.5; SP; SAP |
| | 3b | Important changes to methods after trial commencement (such as eligibility criteria), with reasons | Methods Pg 12; SI; sections 1.2.2; 1.5.1; SP; SAP |
| Participants | 4a | Eligibility criteria for participants | Methods Pg 7,9; SI section 1.3.1; SP, SAPI |
| | 4b | Settings and locations where the data were collected | Methods Pg 7; SI sections 1.1, Fig 1, S1; SP, SAPI |
| Interventions | 5 | The interventions for each group with sufficient details to allow replication, including how and when they were actually administered | Methods Pg 8; SI sections 1.3.2, Fig. 2, S1 |
| Outcomes | 6a | Completely defined pre-specified primary and secondary outcome measures, including how and when they were assessed | Methods Pg 7-8; SI section 1.2.2, 1.5; SP, SAP |
| | 6b | Any changes to trial outcomes after the trial commenced, with reasons | Methods Pg 12; SI section 1.5.1; SAP |
| Sample size | 7a | How sample size was determined | Methods Pg 7; SAP |
| | 7b | When applicable, explanation of any interim analyses and stopping guidelines | N/A |
| Randomisation: | | | |
| Sequence generation | 8a | Method used to generate the random allocation sequence | Methods Pg 8; SI section 1.2.1 |
| | 8b | Type of randomisation; details of any restriction (such as blocking and block size) | Fig 1,2; SI section 1.2.1 |
| Allocation concealment mechanism | 9 | Mechanism used to implement the random allocation sequence (such as sequentially numbered containers), describing any steps taken to conceal the sequence until interventions were assigned | Methods Pg 8; SI section 1.2.1, 1.3.2 |
| Implementation | 10 | Who generated the random allocation sequence, who enrolled participants, and who assigned participants to interventions | Methods Pg 8-9; SI section 1.2.1, 1,2,2, 1.3.1 |

| | | | |
|---|---|---|---|
| Blinding | 11a | If done, who was blinded after assignment to interventions (for example, participants, care providers, those assessing outcomes) and how | Methods Pg 8; SI section 1.2.1, 1.3.2, |
| | 11b | If relevant, description of the similarity of interventions | Methods Pg 8; SI section 1.3.2 |
| Statistical methods | 12a | Statistical methods used to compare groups for primary and secondary outcomes | Methods Pg 10-12; SI section 1.5; SP, SAP |
| | 12b | Methods for additional analyses, such as subgroup analyses and adjusted analyses | Methods Pg 10-12; SI section 1.5, SP, SAP |
| **Results** | | | |
| Participant flow (a diagram is strongly recommended) | 13a | For each group, the numbers of participants who were randomly assigned, received intended treatment, and were analysed for the primary outcome | Results Pg 12-13; Results Fig. 3. (Subject flowchart); SI section 2.1 |
| | 13b | For each group, losses and exclusions after randomisation, together with reasons | Results Pg 12-13; Results Fig. 3. and Fig. S2 (Subject flowcharts); SI section 2.1 |
| Recruitment | 14a | Dates defining the periods of recruitment and follow-up | Results Pg 7, 12-13; Results Fig 2,,Fig. 3. and Fig. S2 (Subject flowcharts); SI section 1.2, 1.3 |
| | 14b | Why the trial ended or was stopped | N/A |
| Baseline data | 15 | A table showing baseline demographic and clinical characteristics for each group | Table 1; Table S5, S6, S7 |
| Numbers analysed | 16 | For each group, number of participants (denominator) included in each analysis and whether the analysis was by original assigned groups | Results Pg 12-13; Results Fig. 3. and Fig. S2 (Subject flowcharts); SI sections 2.5, Table S5,S6, S7 |
| Outcomes and estimation | 17a | For each primary and secondary outcome, results for each group, and the estimated effect size and its precision (such as 95% confidence interval) | Results Pg 13-14; Table 2, SI sections 2.5, 2.6, Table S8-S11, Fig S4, S6,S7 |
| | 17b | For binary outcomes, presentation of both absolute and relative effect sizes is recommended | Results Pg 13-14; Table 2; SI section 2.5, Table S4, S5, Fig S4, S6, S7 |
| Ancillary analyses | 18 | Results of any other analyses performed, including subgroup analyses and adjusted analyses, distinguishing pre-specified from exploratory | Results Pg 13-14; SI sections 2.6; Table S6-S11; |
| Harms | 19 | All important harms or unintended effects in each group (for specific guidance see CONSORT for harms) | Results Pg 14-15; SI sections 2.8 |
| **Discussion** | | | |
| Limitations | 20 | Trial limitations, addressing sources of potential bias, imprecision, and, if relevant, multiplicity of analyses | Discussion Pg 15-16; SI section 1.5.1, 2.5, 2.6 |
| Generalisability | 21 | Generalisability (external validity, applicability) of the trial findings | Discussion Pg 16; SI section 1.5.1, 2.5, 2.6 |
| Interpretation | 22 | Interpretation consistent with results, balancing benefits and harms, and considering other relevant evidence | Discussion Pg 16; |
| **Other information** | | | |
| Registration | 23 | Registration number and name of trial registry | Abstract Pg 4; Methods Pg 6 |
| Protocol | 24 | Where the full trial protocol can be accessed, if available | Submitted as SI |
| Funding | 25 | Sources of funding and other support (such as supply of drugs), role of funders | Funding Disclosure Pg 20-21 |
